# A clinically relevant morpho-molecular classification of lung neuroendocrine tumours

**DOI:** 10.1101/2025.07.18.25331556

**Authors:** Alexandra Sexton-Oates, Émilie Mathian, Noah Candeli, Yuliya Lim, Catherine Voegele, Alex Di Genova, Laurane Mangé, Zhaozhi Li, Tijmen van Weert, Lisa M. Hillen, Ricardo Blázquez-Encinas, Abel Gonzalez-Perez, Maike L. Morrison, Eleonora Lauricella, Lise Mangiante, Lisa Bonheme, Laura Moonen, Gudrun Absenger, Janine Altmuller, Cyril Degletagne, Odd Terje Brustugun, Vincent Cahais, Giovanni Centonze, Amélie Chabrier, Cyrille Cuenin, Francesca Damiola, Vincent Thomas de Montpréville, Jean-François Deleuze, Anne-Marie C. Dingemans, Élie Fadel, Nicolas Gadot, Akram Ghantous, Paolo Graziano, Paul Hofman, Véronique Hofman, Alejandro Ibáñez-Costa, Stéphanie Lacomme, Nuria Lopez-Bigas, Marius Lund-Iversen, Massimo Milione, Lucia Anna Muscarella, Sergio Pedraza-Arevalo, Corinne Perrin, Gaetane Planchard, Helmut Popper, Luca Roz, Angelo Sparaneo, Wieneke Buikhuisen, José van den Berg, Margot Tesselaar, Jaehee Kim, Ernst Jan M Speel, Séverine Tabone-Eglinger, Thomas Walter, Gavin M. Wright, Justo P. Castaño, Lara Chalabreysse, Liming Chen, Christophe Caux, Marco Volante, Nicolas Girard, Jean-Michel Vignaud, Esther Conde, Audrey Mansuet-Lupo, Luka Brcic, Giuseppe Pelosi, Mauro Giulio Papotti, Sylvie Lantuejoul, Jules Derks, Talya Dayton, Nicolas Alcala, Matthieu Foll, Lynnette Fernandez-Cuesta

## Abstract

Lung neuroendocrine tumours (NETs, also known as carcinoids) are rapidly rising in incidence worldwide but have unknown aetiology and limited therapeutic options beyond surgery. We conducted multi-omic analyses on over 300 lung NETs including whole-genome sequencing (WGS), transcriptome profiling, methylation arrays, spatial RNA sequencing, and spatial proteomics. The integration of multi-omic data provides definitive proof of the existence of four strikingly different molecular groups that vary in patient characteristics, genomic and transcriptomic profiles, microenvironment, and morphology, as much as distinct diseases. Among these, we identify a new molecular group, enriched for highly aggressive supra-carcinoids, that displays an immune-rich microenvironment linked to tumour—macrophage crosstalk, and we uncover an undifferentiated cell population within supra-carcinoids, explaining their molecular and behavioural link to high-grade lung neuroendocrine carcinomas. Deep learning models accurately identified the Ca A1, Ca A2, and Ca B groups based on morphology alone, outperforming current histological criteria. The characteristic tumour microenvironment of supra-carcinoids and the validation of a panel of immunohistochemistry markers for the other three molecular groups demonstrates that these groups can be accurately identified based solely on morphological features, facilitating their implementation in the clinical setting. Our proposed morpho-molecular classification highlights group-specific therapeutic opportunities, including DLL3, FGFR, TERT, and BRAF inhibitors. Overall, our findings unify previously proposed molecular classifications and refine the lung cancer map by revealing novel tumour types and potential treatments, with significant implications for prognosis and treatment decision-making.

## Introduction

Neuroendocrine neoplasms (NENs), originating from the diffuse neuroendocrine system, can arise in almost any tissue. They represent a diverse group of neoplasms with varying clinical features, pathology, and prognosis^1^. Lung neuroendocrine tumours (NETs) and carcinomas (NECs, including small-cell lung cancer [SCLC] and large-cell neuroendocrine carcinoma [LCNEC]) are considered distinct diseases due to their differing aetiologies, with NECs strongly linked to smoking, unlike NETs^2,3^. Lung NETs, also classically known as typical and atypical carcinoids, account for 2% of primary lung cancers, with rising incidence, and largely unknown aetiology and pathogenesis^4,5^. According to the World Health Organisation (WHO) Classification of Thoracic Tumours, mitotic count and necrosis differentiate grade-1 typical from grade-2 atypical carcinoids, with five-year survival rates exceeding 90% and 60%, respectively^3,6^. The current morphological classification has limited predictive value for treatment response, and its prognostic value is hindered by significant inter-observer variability in distinguishing typical from atypical carcinoids^7^. Our recent work on emerging markers and deep learning to address this inter-observer variability suggests that the potential of current morphological criteria for classifying lung NETs into typical and atypical types may have been exhausted^8^. In a previous study, we identified three distinct molecular groups with different molecular features (Ca A1, Ca A2, and Ca B^9,10^), validated in independent studies, each comprising a mixture of typical and atypical tumours, and which are starting to guide clinical research^11–14^. However, the differing information provided by the morphological and molecular classifications poses a dilemma for their implementation in clinical practice, and clarification is required regarding the relationship between and utility of the two lung NET classification systems^1,15^. Our previous results also pose a challenge to lung NEN classification in general, given that we identified a new entity, the “supra-carcinoid,” with low-grade morphology but molecular and clinical features resembling those of high-grade NENs (SCLC and LCNEC)^9^. The discovery of supra-carcinoids further complicates current classification and clinical management by questioning the assumption that lung NETs and NECs are unrelated diseases^16^. Additionally, the limited number of samples has hindered the identification of diagnostic or targetable features of this aggressive group.

In the current study, we leverage a comprehensive dataset of multi-omic data on 319 histopathologically annotated tumours and associated whole-slide images, primarily from the lungNENomics network^8^ (**Extended Data Fig. 1a**), performing state-of-the-art spatial omics and deep learning analyses to establish a unified morpho-molecular classification of lung NETs aimed at improving prognosis and treatment. We identify the fourth distinct molecular group of lung NETs as being supra-carcinoid enriched (sc-enriched), and that each of the molecular groups (Ca A1, Ca A2, Ca B, and sc-enriched) present with unique aetiology, clinical and epidemiological features, molecular profiles, microenvironmental niches, and evolutionary histories, suggestive of different diseases. These molecular groups are independent of but complementary to the current morphological classification, with molecular data revealing tumour biology and guiding future clinical interventions, while grading, as in the current WHO classification, remains key for prognostication. We also provide compelling morphological and molecular data characteristic of supra-carcinoids, explaining their biological link with lung NECs. These data consolidate supra-carcinoids as a new biological entity and provide a window into the so far unrecognised lung NEN plasticity.

### Supra-carcinoids: the fourth biologically distinct group of lung neuroendocrine tumours

We conducted Multi-Omics Factor Analysis (MOFA^17^) incorporating gene expression, DNA methylation, small and structural variants, and copy number variant data on a cohort of 319 lung NETs, enriched for the aggressive atypical type, primarily sourced from the lungNENomics network (**Extended Data Fig. 1a, Supplementary Figs. S1 and S2, Supplementary Tables S1-S5**). Archetype analyses (ParetoTI^18,19^) identified four distinct molecular profiles, aligning with the three previously known molecular groups (Ca A1, Ca A2, and Ca B^9^) and a fourth, enriched for supra-carcinoids (sc-enriched or sc-e, 14/16 tumours were supra-carcinoid, **Fig. 1a**, **Extended Data Fig. 1b, Supplementary Fig. S3, Supplementary Tables S1 and S6**). Both grade-1 (typical) and grade-2 (atypical) NETs were found in each group, as well as differing proportions and distinct variations in sex, age, and tumour location (**Fig. 1b, Supplementary Tables S1 and S7**, *P* value < 0.05 for all features cited as enriched thereafter, Binomial tests). Ca A1 was enriched for females (80%), patients aged over 40 (90%), typical tumours (60%), and distal locations (72%). Similarly, Ca A2 was enriched for females (64%) and typical tumours (65%), but featured younger patients (42% under 40, comprising nearly all under 40 cases), and mainly proximal tumours (90%). Ca B showed no predominance for a particular tumour type or location but was enriched for males (77%) and patients over 40 (97%). The sc-enriched molecular group displayed no statistically significant enrichment but had a higher proportion of atypical tumours compared with other groups (62%, *P* value = 1.4 × 10^−4^ to 0.035, Fisher’s exact tests). Regarding potential causative factors, no smoking history differences were found, but we noted a significant difference in asbestos exposure between groups (*q* value = 0.03, Fisher’s exact test), albeit in low numbers of patients (*n* = 1 in Ca A1 and Ca A2, *n* = 2 in sc-enriched, and *n* = 5 in Ca B, **Supplementary Tables S1 and S7**). Notably, 19 of 26 patients with a prior cancer diagnosis belonged to Ca A1, a higher number than in Ca A2 and Ca B (*n* = 3 and 1, *P* values = 2.14 × 10^−3^ and 7.22 × 10^−4^, respectively, Fisher‘s exact tests). The remaining three patients with a previous cancer diagnosis were sc-enriched, also significantly more than in Ca A2 and Ca B (*P* values = 1.38 × 10^−2^ and 5.62 × 10^−3^, respectively, Fisher’s exact tests, **Supplementary Tables S1 and S7**). Survival analysis revealed significant differences among groups (**Fig. 1c**). Ten-year overall survival (OS) rates were 86% for Ca A1, 83% for Ca A2, 70% for Ca B, and 60% for sc-enriched (**Supplementary Table S8**). Atypical carcinoids showed worse survival than typical carcinoids when corrected for molecular group (overall and event-free survival *P* values < 1 × 10^−4^, Wald tests, **Supplementary Table S8**), suggesting that both molecular groups and morphological types have orthogonal prognostic values.

**Figure 1.**
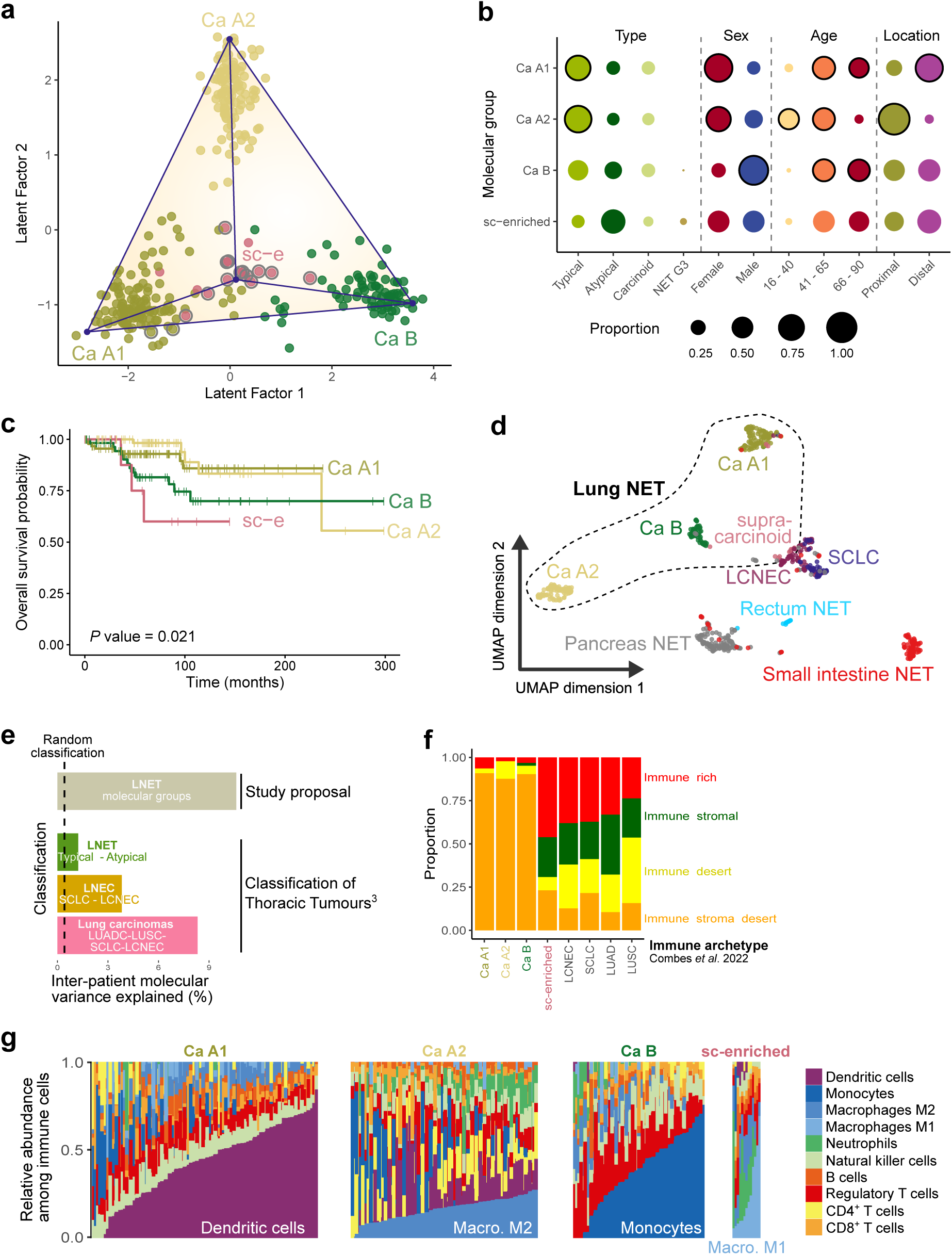
Molecular groups of lung NETs and their clinical and microenvironmental features. **a**, 319 tumours from the lung neuroendocrine tumour (NET) cohort plotted over MOFA Latent Factors 1 and 2, contained within a tetrahedron (blue lines) formed by four phenotypic archetypes (blue dots) identified through ParetoTI. Archetype positions are labelled by molecular group, and tumours are coloured by molecular group membership (*n* = 121 Ca A1, *n* = 107 Ca A2, *n* = 75 Ca B, *n* = 16 sc-enriched). Supra-carcinoids are circled in grey (*n* = 16). sc-e, supra-carcinoid enriched. **b**, Proportion per molecular group of each category within four feature groups: type (where “carcinoid” indicates tumours that could not be definitively classified as typical or atypical), sex (inferred from omics data), age category, and tumour location. Molecular group proportions per category within a feature group sum to 1. Circled features indicate significant enrichment within group (*P* value < 0.05, Binomial test). **c**, Kaplan-Meier plot of overall survival probability over time in months per molecular group (*P* value = 0.021, Log-rank test). **d**, Uniform Manifold Approximation and Projection (UMAP) of transcriptomes of neuroendocrine neoplasms from multiple organs (*n* = 641). NET, neuroendocrine tumour. **e**, Proportion of variance in gene expression explained by the proposed and existing thoracic tumour classifications, where 0% indicates the classification explains no difference in gene expression profiles between patients and 100% indicates that all differences between patients are explained by the classification. The dashed line corresponds to the value expected under a random classification, obtained using permutations of the molecular group labels. LNEC, lung neuroendocrine carcinoma; SCLC, small cell lung cancer; LCNEC, large cell neuroendocrine carcinoma; LUADC, lung adenocarcinoma; LUSC, lung squamous cell carcinoma. **f**, Proportion of samples per molecular group assigned to four immune archetypes. **g**, Relative abundance of ten immune cell types, in each molecular group, estimated from RNA-seq data using deconvolution (software quanTIseq). Individual samples are shown on the x-axis, relative abundances on the y-axis. **f-g** display lung NET cohort samples for which RNA-seq data were available (*n* = 273).

As lung NETs are one of many types of neuroendocrine neoplasms, we wanted to investigate the position of lung NETs within the spectrum of lung NENs and NETs from other body sites. Dimension-reduction analysis of RNA-seq data from 393 lung NENs and 241 gastroenteropancreatic NETs^20–22^ revealed that only the lung NETs formed distinct within-organ molecular groups, while other NETs clustered homogenously by their organ of origin (**Fig. 1d**). This indicates that strikingly different within-organ molecular groups are a unique feature of lung NETs. As a measure of molecular group distinctiveness, as exemplified by **Fig. 1d**, and how well our four-group classification captures differences between lung NET patients in comparison to the existing morphological classification, we calculated the percentage of molecular variation explained by the current classifications. We found that the four groups explain more inter-patient molecular variance than the WHO Classification of Thoracic Tumours^3^, be that of lung NETs, lung NECs, or even that of all lung carcinomas (**Fig. 1e**). This demonstrates the potential of lung NET molecular groups to be the basis of a new, more biologically grounded classification.

Lung NET molecular groups are also associated with distinct tumour microenvironments (TMEs). Our previous study identified dendritic cell enrichment in Ca A1 and monocyte enrichment in Ca B^9^, which was confirmed in a single-cell analysis study that concluded tumour-intrinsic features drive immune cell differences^23^. Using transcriptomic data and cell deconvolution methods^24,25^, we found the sc-enriched group had significantly higher immune infiltration and cancer-associated fibroblast (CAF) levels (**Extended Data Fig. 1c-d, Supplementary Tables S9 and S10**), aligning it with recently proposed immune- and stromal-rich tumour immune archetypes^26,27^ (**Fig. 1f, Supplementary Fig. S4, Supplementary Table S11**). Each molecular group showed a dominant immune cell type: dendritic cells in Ca A1, alternatively activated (“M2”) macrophages in Ca A2, monocytes in Ca B, in line with the male bias in this group^28^, and classically activated (“M1”) macrophages^24,29^ in sc-enriched, though cell proportions varied within groups (all *P* values < 0.05, Pearson correlation tests, **Fig. 1g, Supplementary Tables S9 and S12**). Further to this, the molecular groups Ca A1, Ca A2 and Ca B could also be distinguished through the expression of immune-related proteins within tumoural and microenvironmental regions (**Supplementary Figs. S5, S6 and S7**, **Supplementary Tables S13 and S14**).

These findings confirm the distinctiveness and robustness of the molecular groups and establishes supra-carcinoids as a clear molecular group rather than isolated cases. The molecular groups are associated with unique clinical, epidemiological, and microenvironmental characteristics, potentially suggestive of different carcinogenic processes.

### Lung NET molecular groups harbour specific genomic drivers

As compared to 25 cancer types in the Pan Cancer Analysis of Whole Genomes (PCAWG) consortium^30^, lung NETs have relatively quiet genomes, with below median levels of structural variants (SVs, 11 per tumour in lung NETs versus 64 in PCAWG), copy number variants (CNVs, 4% of genome amplified versus 8% in PCAWG, and 2% deleted versus 3% in PCAWG), and small variants (SNVs, 2175 per tumour versus 6243 in PCAWG, **Supplementary Fig. S8a-b, Supplementary Tables S15-S20**). However, the four molecular groups show distinct genomic profiles as compared to one another. The sc-enriched, Ca B, and Ca A1 groups have the highest small variant burden (**Supplementary Fig. S8c, Supplementary Table S20**), mostly due to a higher contribution of endogenous mutational processes (age-related COSMIC signatures SBS1, and SBS8, possibly due to late replication errors, **Supplementary Fig. S9a-b**, **Supplementary Table S21**). The sc-enriched and Ca A1 groups also have the largest SV burden, primarily due to more numerous translocations (R2 SV signature) and deletions (**Supplementary Fig. S8d-f, Supplementary Table S21**). In accordance with the greater small variant, SV, and deletion burden of sc-enriched samples, signature variability analysis^31^ showed that sc-enriched tumours display significantly more diverse mutational repertoires, suggesting that they undergo more mutational processes than tumours from other groups (**Supplementary Fig. S9c, Supplementary Table S21**). In contrast to other groups, Ca A2 have the least of all variants except for amplifications (**Supplementary Fig. S8e, Supplementary Table S20**). Given the association between genomic instability and increased malignancy in cancer, and the observation that molecular groups contain both grade-1 and 2 tumours (**Fig. 1b**), we additionally investigated genomic alterations by histological type. We found that the types also differ in mutational burdens, with grade-2 tumours presenting more small variants, SVs, and deletions than grade-1, in accordance with the poorer prognosis of grade-2^32^ (**Supplementary Fig. S8c-f, Supplementary Fig. 10d, Supplementary Table S20**). Interestingly, we find that the effect of grade is nested within that of molecular types, with grade-2 Ca A1 having generally more small variants, SVs and deletions that their grade-1 counterparts, grade-2 Ca A2 having more small variants, SVs, and amplifications than grade-1 Ca A2, and grade-2 Ca B having more deletions than grade-1 Ca B (**Extended Data Fig. 2a, Supplementary Table S20**). This suggests that grade-2 tumours may have accumulated greater numbers of alterations in a molecular group-specific pattern, i.e. deletions in Ca A1, and amplifications in Ca A2.

Next, we used computational methods to identify driver alterations involved in carcinogenesis. Eight significantly recurrent CNV events were identified as drivers at the cohort level, and an additional three at the level of individual molecular groups, of which most were chromosome-arm or whole-chromosome events. These were amplifications of chromosomes *(chr) 4* (Ca B-driver), *chr 5* (encompassing *TERT*), *chr 7, chr 8* (Ca A2-driver), and *chr 14p* (overall and Ca A1-driver), and deletions of *chr 1p13.3*, *chr 3* (encompassing *BAP1,* overall and Ca A1-driver), *chr 11q13.1* (including the *MEN1* locus), and *chr 11* (Ca B-driver) (**Fig. 2a**, **Extended Data Fig. 2b-c, Supplementary Table S17**). Large-scale chromosomal aberrations were identified with whole genome doubling (WGD) being the most frequent (11%, *n* = 11) and shattered chromosomes indicative of chromothripsis or chromoplexy found in 11% (*n* = 11) (**Fig. 2b**, **Supplementary Fig. S10, Supplementary Table S17**). WGD was observed across all molecular groups (*P* value = 0.75, Fisher’s exact test) while the presence of shattered regions differed by group (*P* value = 0.045, Fisher’s exact test), whereby they were more frequent in sc-enriched compared with Ca B samples (*P* value = 0.025, Fisher’s exact test). No homologous recombination deficiency or microsatellite instability were detected (**Supplementary Tables S17 and S22**). The IntOGen pipeline^33^ discovered ten driver genes based on small variants (**Fig. 2b, Supplementary Table S23**). As previously reported^9^, *MEN1* was the most frequently altered driver gene (loss of function), with somatic small variants found in Ca B (*n* = 9) and sc-enriched (*n* = 2) samples. In Ca B, 45% of sequenced samples had *MEN1* inactivation, either via mutation (8%), deletion (8%, *chr 11q13.1* focal), or both (29%). *ARID1A* was the second most altered driver (loss of function, *n* = 8), followed by *EIF1AX* (activating, *n* = 6), and *ATM* (loss of function, *n* = 5). Two sc-enriched had *BRAF V600E* mutations. *MEN1* alterations were significantly enriched in Ca B (*P* value = 5.0 × 10^−3^), while *BRAF* mutations were exclusive to sc-enriched (*P* value = 0.01), as were *EIFIAX* to Ca A1, though not significantly associated. We have previously reported that chromatin-remodelling genes were frequently mutated in lung NETs^34^. Here we found significant enrichment for epigenetic regulatory genes, and specifically histone modifiers, within genes harbouring damaging small variants (*P* value = 3.1 × 10^−11^ and 6.9 × 10^−7^, Fisher’s exact test, respectively, **Supplementary Table S24**). In line with their higher mutational and CNV burdens, grade-2 tumours also had significantly more driver events than grade-1 overall *(P* values = 0.016 and 0.036, small variants and CNVs, respectively, logistic regression). This grade-effect was additionally apparent within the Ca A1 molecular group (small variant drivers, *P* value = 0.023).

**Figure 2.**
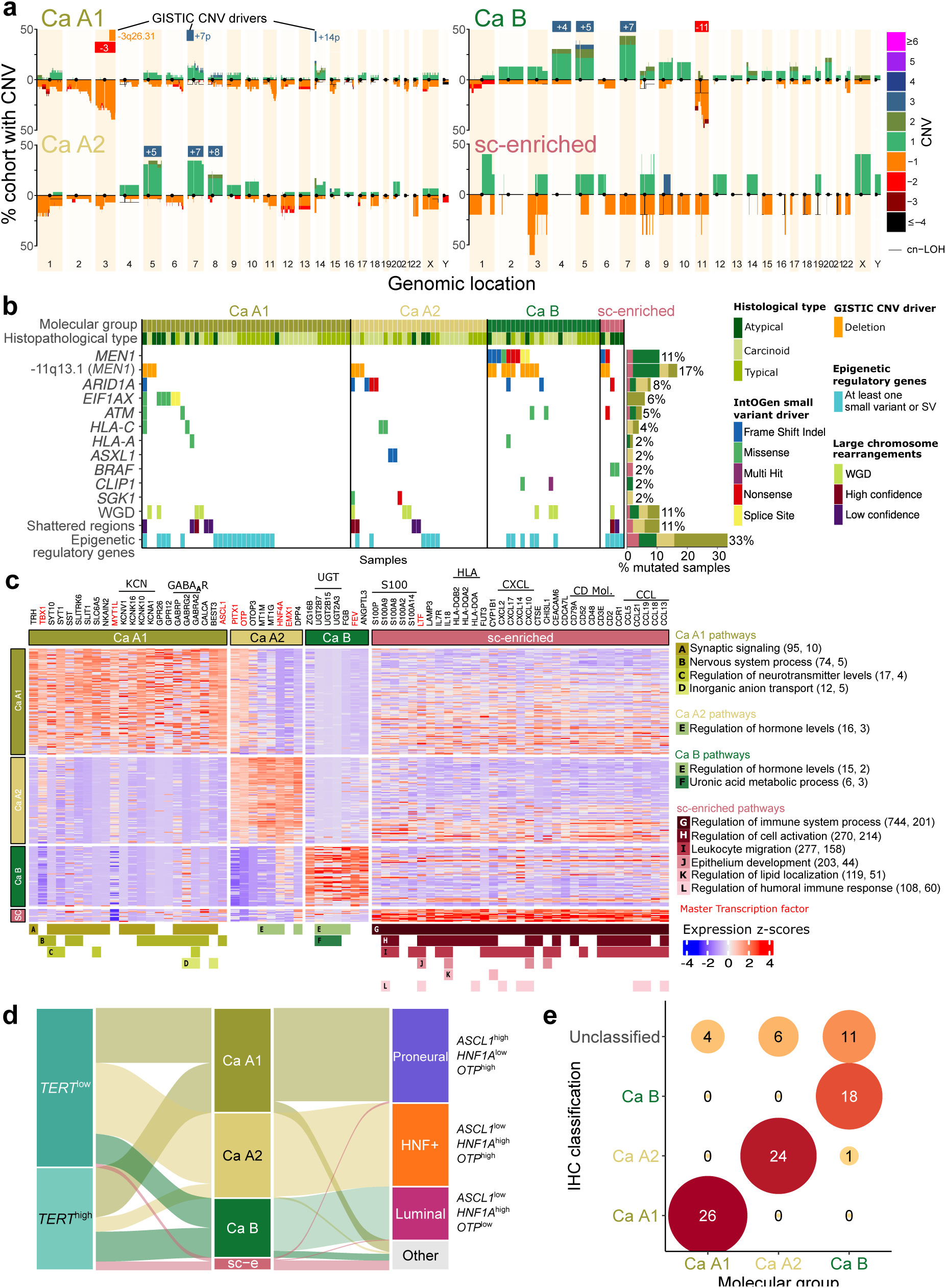
Genomic and transcriptomic profiles of lung NET molecular groups. **a**, Copy number profile of each molecular group; the y-axis represents the percentage of samples in each group with a given alteration (amplifications above the 0 line, deletions below), and colours represent the copy number. The black line represents copy-neutral loss of heterozygosity (cn-LOH). Driver copy number variants (CNVs) detected in each group by the GISTIC2 method are represented by a square above each profile (red rectangles for deletions, blue rectangles for amplifications). **b**, Oncoplot of small and structural variants within driver genes identified by IntOGen, and the presence of at least one small variant or structural variant (SV) in at least one epigenetic regulatory gene (excluding IntOGen drivers already represented above). Copy number deletion of the region containing *MEN1* is also rerepresented. **a-b** display samples from the lung NET cohort with WGS data (*n* = 102). **c**, Expression heatmap of selected genes (columns) for each sample of the lung NET cohort (rows, *n* = 273). Gene expression values are displayed as z-scores of log2(TPM+1) values. Genes are grouped by the molecular group for which they are either (i) a core upregulated gene and in a super-pathway (black), or (ii) a putative master transcription factor (red). Coloured bars on the bottom indicate gene membership in super-pathways, described in the legend on the right. Values between brackets next to the pathways correspond to the number of core upregulated genes (first value), and core pathways (second value), included in the super-pathway. **d**, Alluvial plots comparing the proposed four-group molecular classification with other published classifications, determined by marker gene expression; box sizes correspond to sample sizes in the study cohort, *n* = 273. **e**, Confusion matrix of the performance of the immunohistochemistry (IHC) protein classification for molecular groups, *n* = 90.

Each group also exhibited differences in acquired hallmarks of cancer based on their small variants^35,36^, with sc-enriched tumours showing significantly more hallmarks of cancer impacted by small variants than the other three groups (*P* value = 1.3 × 10^−2^ compared to Ca A1, 7.4 × 10^−3^ compared to Ca A2, and 2.0 × 10^−2^ compared to Ca B, two-sided Mann-Whitney U test, **Extended Data Fig. 2d, Supplementary Table S25**). Three hallmarks were significantly enriched in sc-enriched tumours: cell replicative immortality, escaping immune response to cancer, and proliferative signalling (logistic regression model *P* value = 1.5 × 10^−2^, logistic regression model *P* value = 4.3 × 10^−2^, and Fisher’s exact test *P* value = 3.2 × 10^−2^, respectively, compared to Ca A1, S**upplementary Fig. S11a,b, Supplementary Table S25**). Hallmark acquisition also increased with histological grade, overall and within the Ca A2 molecular group (*P* values = 1.2 × 10^−2^ and 0.047, respectively, **Supplementary Fig. S11c**, **Supplementary Table S25**).

Finally, given the association of Ca A1 with history of cancer, and young age at diagnosis with Ca A2, we investigated pathogenic and likely pathogenic^37^ germline variants within the cohort. One patient with a Ca A1 tumour had a germline *MEN1* alteration and a concurrent report of a neuroendocrine genetic disorder, and two other patients, within Ca A1 and Ca B, were reported to have neuroendocrine genetic disorders, however, no WGS data were available for the latter. Investigation of their RNA-seq profiles was suggestive of germline *MEN1* alterations given the high allelic fractions for these two patients. No additional recurrent germline variants were identified that clearly suggested underlying genetic susceptibility within the cohort, or within a specific molecular group (**Supplementary Table S26**).

In summary, we show that despite their low tumour mutational burden, lung NETs are a disease of the genome, with at least one genomic driver identified in most tumours (**Extended Data Fig. 2e**). We find that these drivers differ by molecular group, and increase in number with grade, further demonstration of the strong differences among molecular groups.

### A unifying molecular classification offering a single framework for patient stratification

To understand the characteristic biological pathways underpinning each molecular group, core upregulated and downregulated genes were identified for each group based on significant positive or negative associations with archetype proportions (**Supplementary Tables S27 and S28**). Expression profiles were also examined to assess the relationship between molecular groups and previously proposed lung NEN subtypes and biomarkers.

Ca A1 displayed the most neuroendocrine-like profile, having the highest expression of lung neuroendocrine gene markers (**Extended Data Fig. 3a, Supplementary Table S29**), and in line with this was the only molecular group to harbour enrichment within core upregulated genes for those highly expressed in lung neuroendocrine cells^38^ (**Supplementary Table S30**). Pathway analysis of core upregulated genes highlighted nervous system functions, including synaptic signaling (*SYT1,* and *SYT10*, and potassium channel (KCN) and GABA_­_receptor families), nervous system processes (GABA_­_receptor family), and regulation of neurotransmitter levels (*SYT1*, *SYT10*, *GABRA2*)^39,40^ (**Fig. 2c**). The enrichment for nervous system function pathways within Ca A1 may suggest a role for the nervous system in carcinogenesis, for instance GABAergic neurons within the TME have been shown to promote tumour cell proliferation and immune escape through interactions with GABA_­_receptors expressed by tumour cells^41^. Alternatively, it could reflect the neuroendocrine origin of Ca A1 and suggests they may be relatively more similar to neuroendocrine cells than other molecular groups. Potential master transcription factors^42^ (TFs) for the Ca A1 molecular group included proneural TFs *ASCL1*^40^ and *MYTL1*^43^ (**Fig. 2c**), and in line with this, almost all Ca A1 tumours were of the proneural regulatory subtype, identified through analysis of cis-regulatory elements in lung NETs by Davis *et al.*^44^ (**Fig. 2d, Supplementary Table S31**). Delta-like protein 3 (*DLL3*), a direct downstream target of ASCL1, was identified as a Ca A1 core upregulated gene in our previous analysis of a smaller cohort^9^ and was significantly positively associated with Ca A1 archetype proportion in the current study, however, it fell just below the fold-change requirement to be classified as a core gene (0.95) (**Extended Data Fig. 3b, Supplementary Table S32**). The elevated *DLL3* gene expression levels observed in Ca A1 tumours have been shown to be correlated with higher protein expression^12^, making these tumours suitable candidates for treatment with DLL3 inhibitors. DLL3 inhibitors have shown promise for the treatment of lung NECs, and proof of concept was recently shown in a case report for lung NET^45^. Antibody drug conjugates (ADCs) targeting DLL3, previously trialed in SCLC^46^, may also be of benefit to Ca A1.

Ca A2 and Ca B shared high expression of HNF genes, including *HNF1A* and *HNF4A*^47^, characteristic of the HNF+ and Luminal regulatory subtypes described by Davis *et al.*^44^ (**Fig. 2d**, **Extended Data Fig. 3c, Supplementary Table S31**). Consistent with their findings^44^, we confirm that Ca A2 and Ca B were associated with higher expression of *FGFR3* and *FGFR4*, which may be amenable to treatment with currently available FGFR inhibitors^48^ (**Extended Data Fig. 3c**). However, Ca A2 and Ca B differed in their expression of the neuronal TF *OTP*^49^, identified as a potential master TF for Ca A2 and a core downregulated gene in Ca B, indicating Ca B likely encompasses the Luminal regulatory subtype (**Fig. 2d, Supplementary Tables S27 and S31**). Low *OTP* expression, as exhibited by Ca B and sc-enriched (**Fig. 2c**), has also been previously associated with poor patient prognosis, as reported by Moonen *et al.*^49^. Ca A2 displayed high *SSTR2* and *SSTR5* expression, therapeutic targets currently exploited in the treatment of both lung and extrapulmonary NENs with somatostatin analogues and radioligands^50–52^ (**Supplementary Tables S32 and S33**). Lastly, Ca B was characterised by downregulation of *NKX2-1* (core downregulated gene), and upregulation of UGT genes, located on amplified *chr 4*, crucial for the elimination of toxic xenobiotic compounds^53^, suggesting environmental exposures may be involved in the development of Ca B (**Fig. 2a,c**, **Extended Data Fig. 3d, Supplementary Table S27**).

Sc-enriched samples had the greatest number of core upregulated genes, most of which were associated with the immune system, including a high proportion of chemokine ligand genes, with 56% of the core upregulated genes linked to regulation of immune system process, leukocyte migration, or regulation of humoral immune response (**Fig. 2c, Supplementary Tables S27 and S28**). The prominence of immune system pathway overexpression is in line with the findings that sc-enriched were most often classified as having an immune-rich microenvironment archetype (**Fig. 1f, Supplementary Table S11**) and suggests an opportunity for immunotherapy treatment for this group. We subsequently examined expression of a panel of 18 genes that capture a T-cell inflamed phenotype and is associated with response to PD-1 inhibitor pembrolizumab in nine different cancer types^54^. Mean expression was significantly higher in sc-enriched than other molecular groups (*P* values < 8.22 × 10^−9^, **Supplementary Table S34**), a promising finding given the higher expression observed in pembrolizumab responders^54^. An additional potential therapeutic option for sc-enriched may be an ADC targeted to TROP2 (*TACSTD2*), a core upregulated gene in sc-enriched, that has been investigated for use in SCLC^46^. Unlike other molecular groups, the sc-enriched core upregulated genes were enriched for hallmarks of cancer (seven out of ten), including escaping immune response to cancer and proliferative signalling (*q* values < 0.05, Fisher’s Exact Tests, **Extended Data Fig. 3e**), both also identified through small variant hallmark enrichment (**Supplementary Fig. S11, Supplementary Table S25**). In line with this, sc-enriched tumours had the highest proliferation index^55,56^, even when considering grade-2 tumours only (**Supplementary Tables S1, S33, and S35**).

Given the similarity between supra-carcinoid and lung NEC molecular profiles^9,10^ (**Supplementary Table S4**), we examined the alignment between lung NETs and proposed markers for TF-driven subtypes of SCLC. SCLC-A, SCLC-N, SCLC-P, and SCLC-I are characterised by elevated *ASCL1*, *NEUROD1*, or *POU2F3* expression, or low expression of all three alongside an inflamed gene signature and *YAP1* expression, respectively^57^. Our analysis revealed that Ca A1, Ca A2 and Ca B lung NETs either fell into the SCLC-A subtype (mainly Ca A1, aligning with the proneural lung NET subtype), or were negative for all three markers (**Extended Data Fig. 3f**). In the case of sc-enriched, they displayed the highest inflamed score (SCLC-I), but interestingly approximately half were also *ASCL1*-high (**Extended Data Fig. 3f,g**).

Finally, we examined the relationship between molecular groups and the recently proposed marker of lung NET prognosis, *TERT* expression. High expression of *TERT* was found to be predictive of prognosis independently of tumour grade^58^. Here, we found that while *TERT* was a core downregulated gene in Ca A2 (**Extended Data Fig. 3h, Supplementary Table S27**), a subset of tumours in Ca A1 and Ca B were *TERT* high, and importantly, high expression was associated with worse prognosis within these molecular groups, making it a therapeutic option for a subset of these tumours (**Fig. 2d**, **Supplementary Table S36**).

The current morphological classification, despite its prognostic value, provides little information for therapeutic management. In contrast, our findings demonstrate the ability of molecular groups to provide a clear framework for patient treatment stratification, notably DLL3 and EGFR inhibitors^59^, and DLL3 ADCs for Ca A1, FGFR inhibitors for Ca A2 and Ca B, SSTR analogues and radioligands for Ca A2, BRAF inhibitors, TOPO2 ADCs, and immunotherapies for supra-carcinoids, and TERT inhibitors for the most aggressive forms of Ca A1, Ca B, and sc-enriched. These molecular groups also capture previously proposed classifications by Laddha *et al*.^11^, and Davis *et al*.^44^ and even partially overlap with SCLC subtypes^57^. Given these features, we believe our molecular groups are the ideal classification to combine with existing morphological (or novel biomarker) grading, for optimal clinical management. A key challenge remains in its implementation in clinical practice. To address this, we evaluated the recently proposed immunohistochemistry (IHC) lung NET classification panel^12^ in our cohort. Leunissen *et al*. demonstrated that a combination of ASCL1, HNF1A, and OTP protein expression could accurately classify a small cohort of lung NETs into their known molecular group (Ca A1, Ca A2, and Ca B) based on a subset of our series (*n* = 15). Applying the panel to 90 additional samples in our series (*n* = 30 each of Ca A1, Ca A2, and Ca B) resulted in a predicted molecular group for 77% of samples. Of the classified samples, 99% were correctly predicted, and only one was misclassified (Ca B predicted to be Ca A2) (**Fig. 2e, Supplementary Figure S12, Supplementary Table S37**). Despite a portion of tumours remaining unclassified, these data validate the IHC classification panel proposed by Leunissen *et al*., paving the way for its implementation in the clinical setting.

### Whole-slide image deep learning reveals hidden morphological features linked to molecular groups

Despite the good performance of an IHC panel in distinguishing molecular groups, a number of tumours were unable to be classified, therefore we investigated morphological features to determine whether tumour morphology differed between molecular groups, and thus whether it could have potential use in diagnosis and classification. For this, a self-supervised deep learning model based on Barlow Twins^60^ was trained on patches of whole slide images (WSIs), called tiles, extracted from haematoxylin/eosin or haematoxylin/eosin/saffron (HE/HES) stained slides, that then clustered the tiles into partitions which shared common morphological features (**Fig. 3a Self-supervised branch**). A random forest model trained on tile proportions in relevant morphological partitions performed poorly in predicting histological types, achieving a weighted F1 score of 0.34 (**Fig. 3b, Supplementary Tables S38 and S39, Supplementary Fig. S13**). Interestingly however, applying the same method to predict the three most common molecular groups (Ca A1, Ca A2, and Ca B, for which sufficient samples for analysis were available) from relevant morphological partitions yielded much better performance, with a weighted F1 score of 0.72 (**Fig. 3b, Supplementary Tables S38 and S39, Supplementary Fig. S14**).

**Figure 3.**
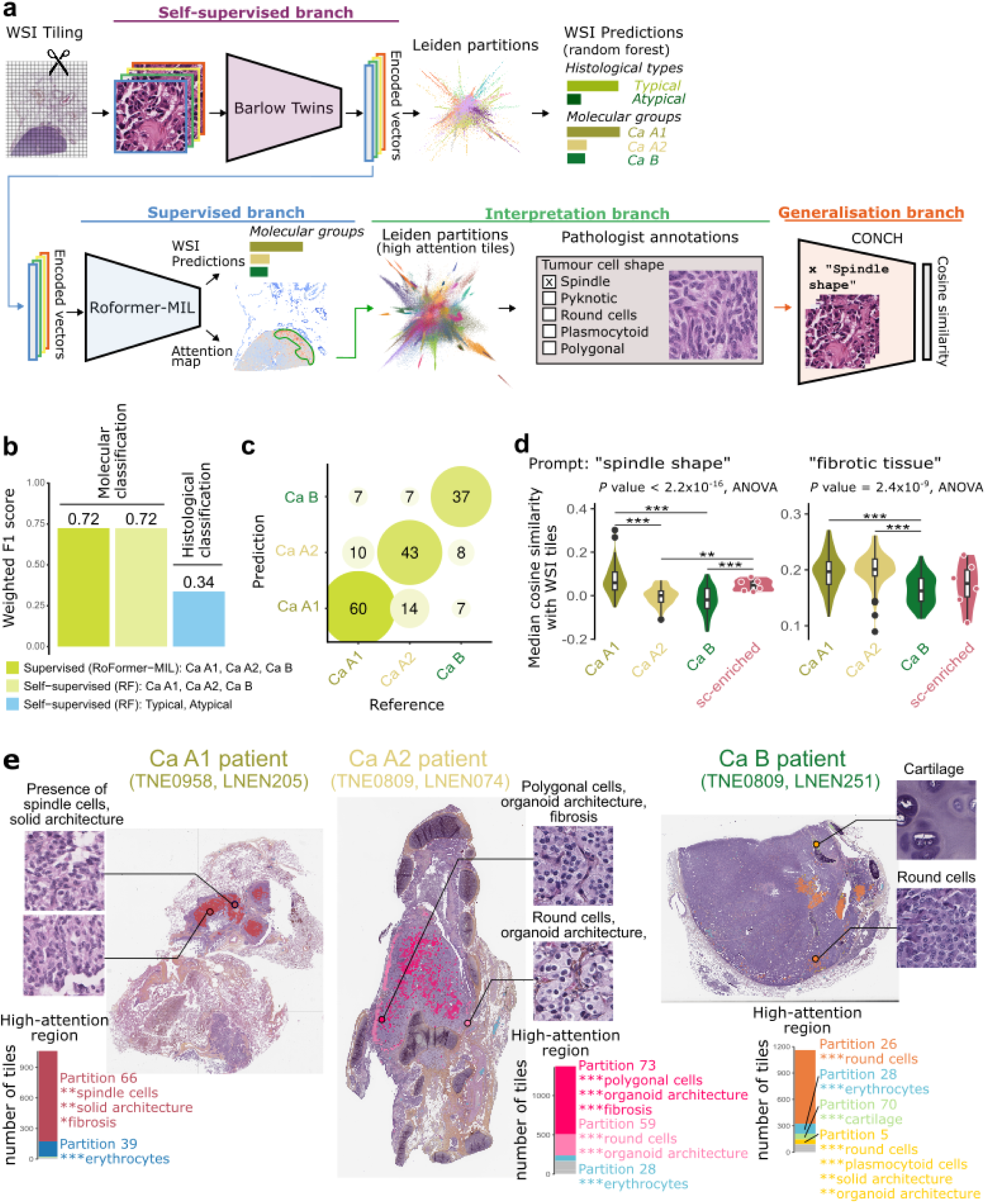
Morphological features of lung NET molecular groups identified by deep learning. **a**, Schematic representation of the deep learning algorithm applied to H&E and H&E/S stained whole-slide images (WSIs). WSIs are divided into smaller regions, referred to as tiles. These tiles are used to train the Barlow Twins algorithm in a self-supervised manner, aiming to generate similar encoded vectors for tiles with common morphological features. The encoded vectors are grouped into morphological partitions using the Leiden clustering algorithm. Tile proportions within each Leiden partition are then used to train random forest models to predict either the tumour histology or the molecular group. A supervised branch, based on RoFormer-MIL, was subsequently employed to identify the most informative tiles for molecular group classification. RoFormer-MIL processes a matrix of encoded vectors from Barlow Twins for each WSI and predicts the molecular group by considering all interactions between the vectors. Attention scores from RoFormer-MIL were used to extract the most relevant tiles, which were clustered using the Leiden algorithm and interpreted by a panel of expert pathologists (Interpretation branch). To generalise the representation of key features across all tiles, cosine similarity scores between tile embedding vectors and text-based prompts were computed using the CONCH visual-language foundation model. **b**, Comparison of deep learning performance for each classification system, measured by weighted F1 score. **c**, Confusion matrix of molecular group predictions by RoFormer-MIL. **d**, Distribution of median cosine similarity scores per WSI for the text prompts indicated above the figure, by molecular groups (x-axis). **e**, Example of how the morphological criteria identified by deep learning could help diagnose patient molecular group. Coloured squares correspond to high-attention tiles identified by RoFormer-MIL to predict molecular groups. Colours represent different partitions, and barplots display the number of tiles from the different partitions in these high-attention regions. Example tiles from partitions along with their consensus pathological annotations from two independent pathologists are also represented, where stars correspond to the significance of the association between partitions and annotations. * 0.01 < *q* value < 0.05; ** 0.001 < *q* value *≤* 0.01; *** *q* value *≤* 0.001.

A supervised deep learning model based on RoFomer-MIL^61^ was then used to identify the most relevant tiles for molecular group classification, via an attention mechanism (**Fig. 3a Supervised branch**), and subsequent morphological examination for a human interpretation of AI results (**Fig. 3a Interpretation branch, Supplementary Fig. S15**). This interpretable deep learning model achieved equally good performance in classifying 193 WSIs into molecular groups, with a weighted F1 score of 0.72 (**Fig. 3b,c, Supplementary Tables S39-S41**). Tiles with the highest attention scores (i.e., those most relevant for distinguishing between the groups) were extracted and grouped into morphological partitions. Random tiles (*n* = 1,095 in total, from 128 different patients) from the most promising partitions, i.e., partitions most likely to be informative about cell morphology and patient molecular groups (**Extended Data Fig. 4a, Supplementary Fig. S16, Supplementary Table S42**), were then meticulously annotated by a panel of six expert pathologists to identify whether key morphological features were characteristic of any particular group (**Fig. 3a Interpretation branch, Supplementary Tables S43-S45, Supplementary Figs. S17 and S18**). This analysis demonstrated that Ca A1 tumour cells more frequently had a spindle shape (*q* value < 1 × 10^−4^, captured by partitions 4, 72, 21, 22, 32, 43 and 66, **Supplementary Fig. S17**), in line with a previous report of spindle cell enrichment within a cluster of Ca A1-like carcinoids^62^ (dominated by typical, peripheral tumours in women, distinctive features of Ca A1, **Fig. 1b**), and also in line with the higher epithelial-mesenchymal transition (EMT) score observed in Ca A1 tumours (**Extended Data Fig. 4b, Supplementary Tables S1, S33 and S35**). Ca A1 tumours also more frequently displayed nested cells (*q* value < 1 × 10^−4^, partitions 22, 30, 32, 34, and 66), solid tissue architecture (*q* value = 9.67 × 10^−4^, partitions 5, 21, and 66), cells with clear cell changes (*q* value < 1 × 10^−4^, partitions 22, 32, 34, and 66), and an enrichment for salt and pepper chromatin (*q* value = 4.71 × 10^−3^, 14 partitions, **Supplementary Fig. S17**). Ca A1 and A2 tumours shared an enrichment for fibrotic tissue (*q* value = 9.75 × 10^−3^, partitions 30, 32, 58, and 66, **Extended Data Fig. 4c, Supplementary Fig. S17**). Ca A2 tumours more frequently had a plasmacytoid cells (*q* value < 1× 10^−4^, partitions 5, 21, 34, 43, and 68), oncocytic changes (*q* value = 4.71 × 10^−3^, partitions 22 and 34), and organoid tissue architecture (*q* value < 1 × 10^−4^, partitions 13, 44, 59, 64, 5, 15, 43, 48, and 68, **Extended Data Fig. 4c, Supplementary Fig. S17**). Ca A2 and B tumours also had more frequently an acinar tissue architecture than Ca A1 tumours (*q* value = 0.0263, **Supplementary Fig. S18**). Tumour cell size was also found to be slightly different between the groups, with Ca A1 having marginally but significantly smaller cell sizes (*q* value = 5.80 × 10^−4^) and a smaller nucleus to cytoplasm ratio (*q* value = 0.0346, **Supplementary Fig. S18**). Additional features were highly specific to certain partitions, such as cartilaginous tissue in partition 70, which was enriched for Ca A2 and Ca B (**Extended Data Fig. 4c, Supplementary Fig. S17**). This finding is likely related to the proximal location of these tumours compared with Ca A1 (**Fig. 1b**) and importantly demonstrates the biological and clinical relevance of the model outputs. We then chose to test the generalisation of two key features to the entire WSI, rather than randomly selected tiles, that may allow us to distinguish between groups. We selected spindle cells (associated with Ca A1) and fibrotic tissue (associated with Ca A1 and Ca A2), as they can be easily assessed by pathologists and were among the most statistically significant features associated with the groups. To perform this generalisation, we automatically estimated the presence of the features using the visual-language model CONCH^63^ (**Fig. 3a Generalisation Branch**). This demonstrated that spindle cell shape was indeed characteristic of Ca A1, and interestingly, this feature was also common in supra-carcinoid samples (**Fig. 3d**, **Extended Data Fig. 4c,d**). The analyses further confirmed that the absence of fibrotic tissue was associated with Ca B (**Fig. 3d**, **Extended Data Fig. 4c,d**).

These findings suggest that differences among molecular groups translate into distinct morphological features detectable by deep learning, such as spindle cells in Ca A1 tumours, the presence of fibrotic tissue in Ca A1 and Ca A2, and its absence in Ca B. As an illustration, we show examples of WSIs correctly classified with high confidence by the RoFormer-MIL deep learning model (probability >0.95) in **Fig. 3e**, along with the high-attention regions used by the algorithm that could prompt future pathological examinations. We can see that the algorithm indeed mostly focused on tiles with spindle cells and solid morphology to predict the Ca A1 tumour (partition 66), on regions with organoid tissue architecture to predict the Ca A2 tumour (partitions 59 and 73), and on regions with round cells and cartilage to predict the Ca B tumour (partitions 26, and 70, respectively). Using a selection of these deep learning-identified features, spindle cells, organoid and solid architecture, and fibrosis, the unclassified/misclassified cases from the previous IHC experiment (*n* = 22, **Fig. 2e**) were reassessed, and nine were able to be correctly grouped, despite their status as more complex cases (**Supplementary Figure S12, Supplementary Table S37**). A more detailed morphological evaluation, focusing on the characteristic features of each molecular group identified through the aforementioned WSI deep learning analyses, combined with the validated IHC classification panel, may assist pathologists in achieving a definitive diagnosis in the clinical setting (**Fig. 2e**, **Fig. 3e**).

### Evolutionary trajectory and plasticity of lung NET molecular groups

The existence of distinct molecular groups of lung NETs suggests varied carcinogenesis processes, potentially influenced by differences in cells of origin, genomic events, and the TME. Understanding the origin and temporal evolution of such groups may aid in designing strategies to monitor and intercept tumour progression.

We first explored whether the groups had different cell states^64^, hinting at different cells of origin or varied cell differentiation trajectories after the onset of carcinogenesis. To do so, we compared lung NET bulk RNA-seq-based expression profiles with a panel of single-cell sequencing reference profiles. To accurately represent the potential neuroendocrine cellular states of lung NETs, we used a reference profile made up of epithelial cells from foetal lung tissue airway organoids^65^, enriched for both lower airway progenitor (LAP) and pulmonary neuroendocrine cells^66^, as well as lung NET TME cells (stromal and immune)^23^ to avoid overfitting. Deconvolution of lung NET molecular groups revealed that most tumours contained a mix of the terminally differentiated neuroendocrine cell types, and club cells, however proportions did vary within and between molecular groups (**Fig. 4a, Supplementary Table S9**). Ca A1 had the highest proportion of differentiated neuroendocrine cells (**Fig. 4b, Supplementary Tables S9 and S46**), consistent with its enrichment for characteristic lung neuroendocrine cell genes^38^ and nervous system function pathways found in upregulated genes, as well as the proneural nature of putative driver TFs *ASCL1* and *MYTL1* (**Fig. 2c**, **Extended Data Fig. 3a,f, Supplementary Tables S28-S31**). Ca B contained the highest proportion of club cells, thought to be important in the elimination of xenobiotic compounds^67^ and in line with the high expression of *UGT* genes in Ca B (**Fig. 2c, Supplementary Table S27**), whereas Ca A2 displayed a mixed profile of both differentiated neuroendocrine cells and club cells (**Fig. 4a,b, Supplementary Tables S9 and S46**). The pulmonary NE precursor, LAPs, were found in the highest proportion in sc-enriched samples and were virtually absent in Ca A1, Ca A2 and Ca B (**Fig. 4b, Supplementary Tables S9 and S46**), supporting their loss of neuroendocrine phenotype and suggesting an undifferentiated, stem-like cellular state (**Extended Data Fig. 3a, Supplementary Table S29**). Additionally, sc-enriched tumours had similar levels of LAP and differentiated neuroendocrine cells to LCNEC, reflecting their LCNEC-like molecular profile (**Fig. 4b, Supplementary Tables S4, S9 and S46**). Spatial transcriptomic analysis of four lung NET samples (supra-carcinoids) showed that these cell types occupy distinct spatial locations, with clear separation between terminally differentiated areas and LAP areas (spatial correlations ranging from −0.29 to 0.01, and from −0.36 to 0.05, **Fig. 4c**, **Extended Data Fig. 5a**, **Supplementary Figs. S19-S21, Supplementary Table S47**). Interestingly, this spatial analysis of supra-carcinoids also demonstrated that these tumours contain regions with a molecular profile similar to that of the Ca A1 or Ca B molecular groups as well as sc-enriched (**Supplementary Fig. S19**). As expected from our bulk analyses, we find that presence of the sc-enriched profile was significantly spatially correlated with that of the LAP cell profile (correlations ranging from 0.02 to 0.32), while presence of the Ca A1 and Ca B profiles were significantly spatially correlated with terminally differentiated cell profiles (correlations ranging from 0.14 to 0.67, **Supplementary Figs. S19 and S20, Supplementary Table S47**).

**Figure 4.**
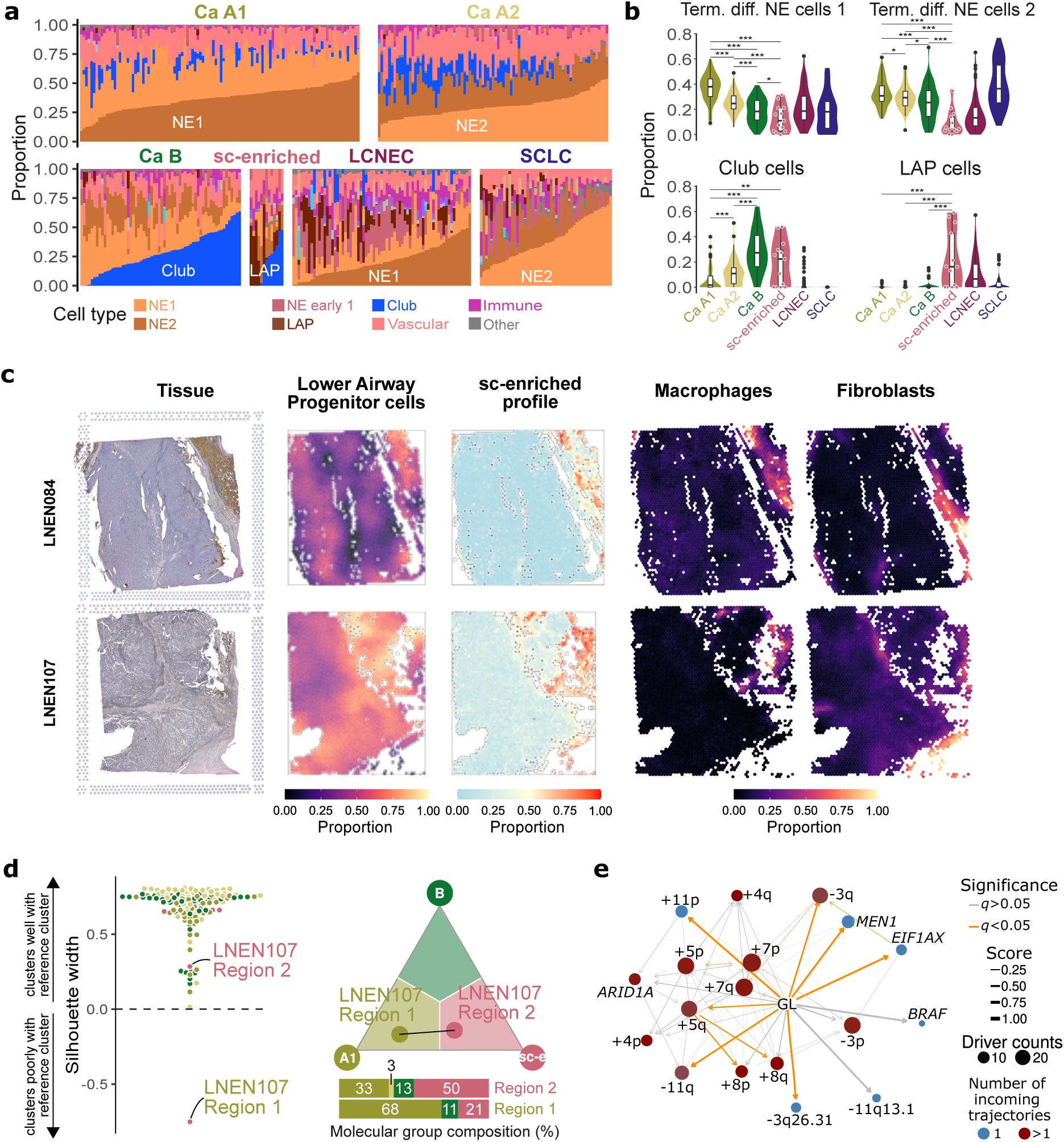
Evolution of lung NET molecular groups. **a**, Estimated proportion (y-axis) of cell types per sample (x-axis), using deconvolution with a custom single-cell reference database including a diverse repertoire of neuroendocrine cells. NE: neuroendocrine; LAP, lower airway progenitor. **b**, Estimated proportion of terminally differentiated NE cells, club cells, and LAPs in the four molecular groups and lung NECs. **a-b** display samples from the lung NET cohort (*n* = 273) and LCNEC (*n* = 69) and SCLC (*n* = 51) for which RNA-seq data is available. Asterisks correspond to significance assessed by t-tests. **c**, Spatial transcriptomics of supra-carcinoid samples. From left to right, columns represent: the H&E image, proportion of lower airway progenitor cell expression profile per spot, proportion of sc-enriched group expression profile per spot, the estimated location and proportions of macrophages and fibroblasts (scaled so 1 is the maximal value observed in a given sample). **d**, Silhouette width of intra-tumoural heterogeneity (ITH) samples, measuring the distance between the sample and its patient reference molecular group (molecular group of the ITH piece included in lung NET cohort MOFA) (left). Archetype analysis of a heterogeneous multi-regional tumour (LNEN107). Barplots correspond to the molecular group proportion composition of each region (right). **e**, Driver to driver trajectories among tumours with identified driver mutations. Arrow colours correspond to the significance of Fisher’s exact test of the association between pairs of drivers (FDR corrected using the Benjamini-Hochberg method), arrow size corresponds to the strength of the inferred association, point size to the number of driver events, and point colour indicates the number of paths leading to a given driver (blue: a single trajectory leads to this driver, brown: multiple trajectories lead to it). GL, germline. * 0.01 < *P* value < 0.05; ** 0.001 < *P* value *≤* 0.01; *** *P* value *≤* 0.001.

The fact that supra-carcinoids harbour mixtures of different molecular components, each in a different microregion, with some resembling Ca A1 or Ca B tumours, suggests the possibility of a dynamic molecular profile where supra-carcinoids arise from the other molecular groups and then stay contained within microregions. To explore this hypothesis, we performed integrative analyses of 91 tumours from 41 patients using multi-regional, multi-omic sequencing. MOFA followed by archetype analysis of each region showed that, in 40 out of 41 patients, the different tumour regions belonged to the exact same molecular group, showing that groups are generally spatially stable (**Fig. 4d**, **Extended Data Fig. 5b, Supplementary Table S48**). Interestingly, the only tumour (LNEN107) that had regions belonging to different groups (Ca A1 and sc-enriched) was a supra-carcinoid, suggesting that transitions between groups, though infrequent, are possible for supra-carcinoids (**Fig. 4d**). Furthermore, within each patient the proportion of sc-enriched archetype per tumour region was the most variable of all archetype proportions (mean difference of 0.080 versus 0.045 for the other archetypes, *P* value = 0.0298, t-test, **Extended Data Fig. 5b, Supplementary Table S48**), suggesting that some Ca A1, A2, and B tumours possess a small and localised supra-carcinoid component. In addition to this intra-tumoural heterogeneity (ITH), we found that the sc-enriched molecular group displayed the most inter-patient heterogeneity of all groups, with mean Euclidean distance between samples of 1.81 versus 0.94-1.31 for the other groups (**Supplementary Table S48**). This variability was also exemplified by two supra-carcinoid samples having molecular profiles closer to that of Ca A1 than to samples from the sc-enriched group (**Table 1, Supplementary Table S49**).

**Table 1.**
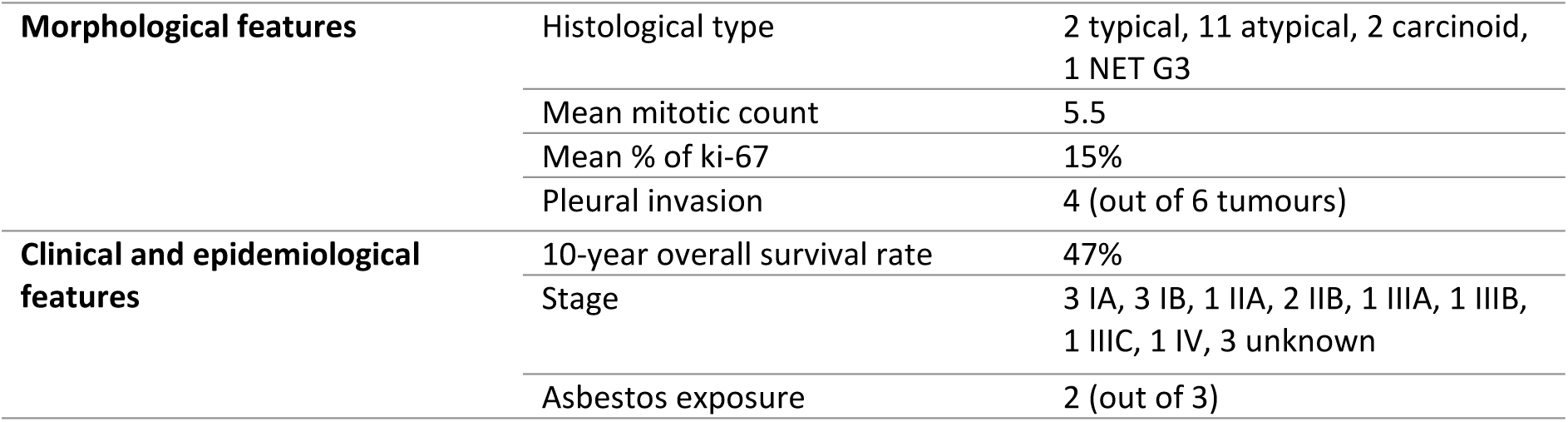
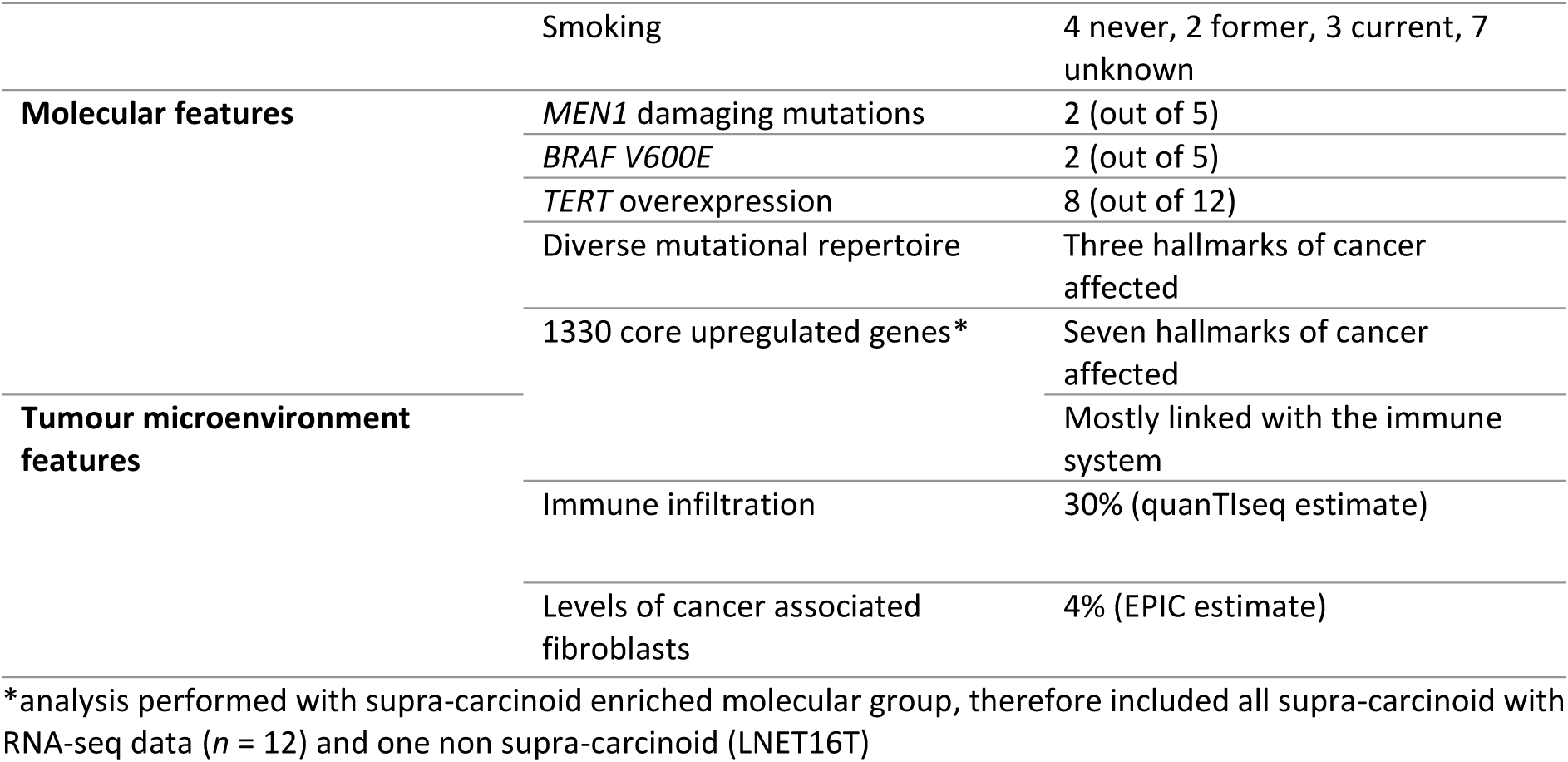
Main features of supra-carcinoids (*n* = 16).

|  |  |  |
| --- | --- | --- |
| **Morphological features** | Histological type | 2 typical, 11 atypical, 2 carcinoid, 1 NET G3 |
|  | Mean mitotic count | 5.5 |
|  | Mean % of ki-67 | 15% |
|  | Pleural invasion | 4 (out of 6 tumours) |
| **Clinical and epidemiological features** | 10-year overall survival rate | 47% |
|  | Stage | 3 IA, 3 IB, 1 IIA, 2 IIB, 1 IIIA, 1 IIIB, 1 IIIC, 1 IV, 3 unknown |
|  | Asbestos exposure | 2 (out of 3) |
|  | Smoking | 4 never, 2 former, 3 current, 7 unknown |
| **Molecular features** | *MEN1* damaging mutations | 2 (out of 5) |
|  | *BRAF V600E* | 2 (out of 5) |
|  | *TERT* overexpression | 8 (out of 12) |
|  | Diverse mutational repertoire | Three hallmarks of cancer affected |
|  | 1330 core upregulated genes* | Seven hallmarks of cancer affected |
| **Tumour microenvironment features** |  | Mostly linked with the immune system |
|  | Immune infiltration | 30% (quanTlseq estimate) |
|  | Levels of cancer associated fibroblasts | 4% (EPIC estimate) |

\*analysis performed with supra-carcinoid enriched molecular group, therefore included all supra-carcinoid with RNA-seq data ( $n = 12$ ) and one non supra-carcinoid (LNET16T)

Most driver mutations were clonal and almost exclusive to a single molecular group, including *EIF1AX* (only in Ca A1), *BRAF* (only in sc-enriched), *MEN1* (9/11 in Ca B), and *ARID1A* (4/6 clonal mutations in Ca A2, and no subclonal mutations in Ca A2), suggesting that molecular groups are determined early by genomic events (**Extended Data Fig. 5c**). This was further corroborated by phylogenetic analyses (REVOLVER method^68^), which identified 26 significant driver-to-driver trajectories (**Fig. 4e, Supplementary Table S48**). These phylogenetic analyses showed that alterations associated with the molecular groups such as *chr 3* deletions, *EIF1AX* mutations, *chr 5* amplifications, and *MEN1* alterations/*chr 11* deletions, were indeed predominantly initiation events, while alterations shared across groups such as *chr 5* and *7* amplifications, and *ARID1A* mutations, were typically subsequent events (**Fig. 4e**, **Extended Data Fig. 5d, Supplementary Table S48).** In contrast, sc-enriched emerged through multiple trajectories which were common to other groups (e.g. *chr 3* deletion, *MEN1* alterations, or *chr 5* amplification, **Extended Data Fig. 5d**), further supporting the hypothesis that supra-carcinoids can emerge from all molecular groups. Interestingly, because *BRAF* V600E mutations were only reported in supra-carcinoids, these mutations might constitute a direct route to this group without the need to pass through others. Grade was associated with the presence of a driver event, with grade-2 tumours more frequently having a driver event than grade-1 (*P* value = 0.011, Fisher‘s exact test), suggesting that genomic driver events may influence tumour proliferative activity. Molecular group was associated with the presence of secondary events, with sc-enriched all having undergone secondary driver events, further suggesting that they may have evolved from other entities (*P* value = 0.049, Fisher‘s exact test, **Extended Data Fig. 4e**, **Supplementary Table S48**).

Although sc-enriched tumours did not have a single defining genomic driver, they shared a common stromal and immune-rich microenvironment characterised by fibroblasts and macrophages (predicted M1) (**Fig. 1g**, **Extended Data Fig. 1c,d, Supplementary Table S9**), hinting that the TME might be the main common driver of supra-carcinoids. Indeed, the spatial transcriptomics data showed that sc-enriched regions within supra-carcinoid samples were relatively small in size, with specific microenvironments, co-localising with macrophages and fibroblasts (**Fig. 4c**, **Extended Data Fig. 5a**, **Supplementary Figs. S19 and S20, Supplementary Table S47**). Nevertheless, it is noteworthy that we have previously successfully established a patient-derived tumour organoid for a supra-carcinoid (LNET10)^59^, suggesting that while the TME seems to play a key role in the development of supra-carcinoids, it may not be essential for maintaining their phenotype and growth.

To further evaluate the role of tumour-TME interactions in lung NET carcinogenesis, we analysed tumour neoantigens. Neoantigens provide both an estimate of the degree to which a tumour is visible to the immune system and can also reveal how much selective pressure the immune system has exerted on the tumour in the past by comparing the non-synonymous to synonymous mutation ratio in antigen-rich versus antigen-poor genomic regions (immune dN/dS ratio)^69^. Supra-carcinoids and Ca A1 tumours exhibited the highest neoantigen counts, though fewer than expected due to purifying selection from immune predation (immune dN/dS ratio < 1, **Extended Data Fig. 5e**). Ca A2 tumours presented very few neoantigens, presumably also due to past immunoediting (ratio < 1), while Ca B tumours presented few neoantigens, but more than expected (dN/dS ratio >1), suggesting the presence of driver mutations among these neoantigens (**Extended Data Fig. 5e**). Interestingly, intrinsic patient characteristics may influence tumour evolution, as patients with Ca B tumours showed significantly higher HLA class I gene diversity (involved in antigen recognition) compared to other groups (**Extended Data Fig. 5e**), suggesting that their immune systems can recognize a broader range of antigens, consistent with findings that HLA diversity can protect against lung cancer^70^. However, this diversity might also drive tumour evolution through selective pressure. These results indicate that, despite their predominantly immune desert nature (**Fig. 1f**), the immune microenvironment of lung NETs actively contributes to tumour cell elimination and influences tumour evolution.

Overall, these findings suggest that the cell state (or potential cell of origin), early genomic events, and the TME all contribute to the development of lung NET molecular groups, and that there may be grade progression within groups, supported by the increased TMB, driver alterations, and hallmarks seen in grade-2. Whilst Ca A1, Ca A2, and Ca B appear to be temporally stable, often determined by early genomic events, with transitions between them being either very rare or occurring very rapidly, the carcinogenic processes leading to the development of supra-carcinoid are less clear. Supra-carcinoids may emerge from other groups, perhaps through the dedifferentiation of neuroendocrine cells towards a LAP-like precursor either driven or accompanied by tumour cell interaction with macrophages and/or fibroblasts, or possibly directly through the acquisition of *BRAF* mutations.

### Supra-carcinoids within the spectrum of lung NEN

In recent years, several studies have reported an emerging morphological entity of lung NETs with carcinoid morphology but higher mitotic counts and Ki-67 levels than expected for such grade-1/2 tumours, similar to the already established grade-3 neuroendocrine tumours described in other organs (NET G3)^71–75^. Like other lung NETs, they display *MEN1* mutations and lack *RB1* and *TP53* alterations, however, these tumours show more aggressive behaviour, with higher rates of postsurgical recurrence than expected for grade-2 (atypical) carcinoids (approximately 20-30%)^15^. Our data show that supra-carcinoids can be, but are not always, NET G3 tumours. Indeed, supra-carcinoids showed more aggressive behaviour than other lung NETs^9^, with more cancer-associated hallmarks, a worse 10-year overall survival rate, and more frequent pleural invasion (67% versus less than 5% in other groups), in line with previous reports indicating that visceral pleura invasion correlates with higher aggressiveness and poorer prognosis, regardless of tumour size^76–78^ (**Fig. 1c**, **Extended Data Fig. 2d**, **Table 1**, **Extended Data Fig. 3e, Supplementary Tables S1, S8 and S49**). However, although these tumours have higher *MKI67* gene expression levels than other molecular groups, the proliferative activity estimated via Ki-67 protein expression shows only a moderate increasing trend, and both Ki-67 index and mitotic count remained within the expected range for grade-1 and grade-2 lung NETs, except for one tumour (**Fig. 5a, Supplementary Table S49**). This sample, a supra-carcinoid with high ki-67 and greater than 10 mitoses/2mm^2^, was classified as NET G3 by consensus during the central pathology review, along with a second sample from the Ca B molecular group (**Fig. 5b**). Furthermore, when considering individual pathologist assessments, four additional samples were annotated by at least one pathologist as having more than 10 mitoses/2mm^2^: one was classified as sc-enriched, one as Ca A2, and the remaining two as Ca B (**Fig. 5b**). This suggests that samples across all molecular groups may have the potential to progress to NET G3 as part of their disease course mirroring the possible progression from grade-1 to grade-2 tumours within each molecular group.

**Figure 5.**
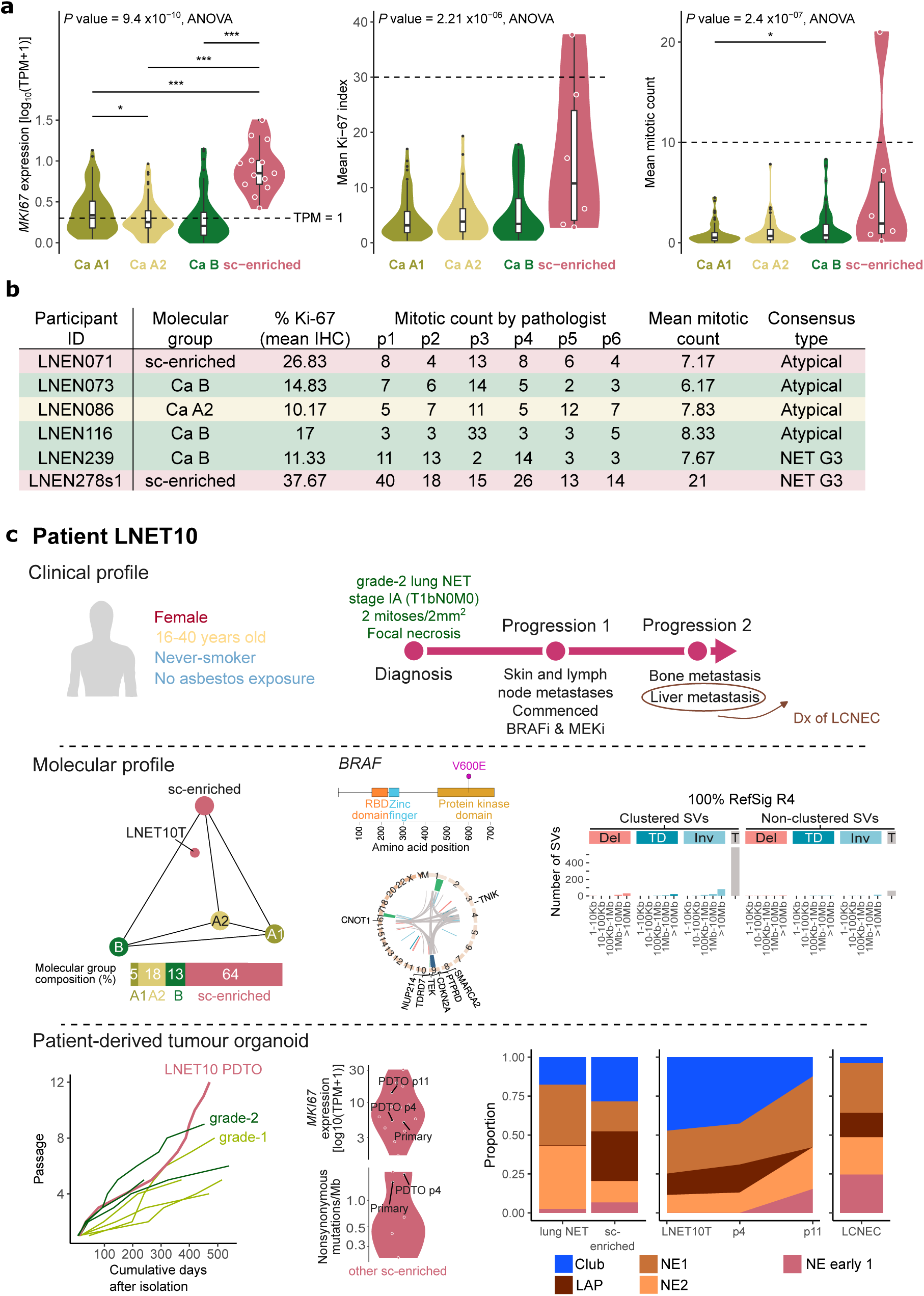
Spotlight on supra-carcinoids. **a**, Distribution of *MKI67* gene expression per molecular group (*n* = 273, left), distribution of mean Ki-67 index (mean of six pathologist’s readings) per molecular group (*n* = 196, middle), distribution of mean mitotic count (mean of six pathologist’s readings) per molecular group (*n* = 197, right). All pairwise comparisons performed with t-tests. * 0.01 < *P* value < 0.05; ** 0.001 < *P* value *≤* 0.01; *** *P* value *≤* 0.001. TPM, transcripts per kilobase million. **b**, Mitotic counts by pathologist and additional features for six samples which were recorded by at least one pathologist to have a mitotic count > 10, i.e. outside the range of grade-2 lung NET, samples are coloured by their molecular group. **c**, Profile of a patient with a supra-carcinoid tumour for which a patient-derived tumour organoid (PDTO) was developed, allowing drug screens. Top panel, clinical profile and disease course. BRAFi, BRAF inhibitor, MEKi, MEK inhibitor, Dx, diagnosis; LCNEC, large cell neuroendocrine carcinoma. Middle panel, molecular profile of primary tumour (LNET10T) showing location in archetype space (left), *BRAF* V600E alteration and shattered region of *chr 9* (middle), and structural variant (SV) signature profile of clustered translocations (T) corresponding to RefSig R4 (right). Del, deletions; TD, tandem duplications; Inv, inversions. Bottom panel, PDTO features of growth trajectory in comparison to other grade-1 and grade-2 lung NETs (left), *MKI67* expression and nonsynonymous mutations per megabase (Mb) for primary tumour, PDTO passage 4 (p4) and passage 11 (p11) in comparison to other sc-enriched tumours (middle), and cell typeproportions (right). Proportions for lung NET (Ca A1, Ca A2 and Ca B), sc-enriched and LCNEC are mean proportion per cell type for *n* = 260, *n* = 13, and *n* = 69 tumours, respectively. LAP, lower airway progenitor; NE, neuroendocrine.

Another controversial question is whether supra-carcinoids may represent a transitional biological entity between lung NETs and NECs, which would then represent the two ends of a spectrum of highly stable tumour groups rather than two completely unrelated diseases^79,80^. Supporting this hypothesis is the recent identification of a subset of SCLC tumours with clinical and genomic similarities to lung NETs^81^. Like carcinoids, these aggressive tumours lacked the hallmark co-inactivation of *RB1* and *TP53* observed in SCLC^82,83^, and arose in light or never smokers, unlike most SCLC cases where >95% occur in heavy smokers^84^. They were additionally characterised by frequent chromothripsis, leading to extrachromosomal amplification. Importantly, several of these atypical SCLC were identified in metastases of patients with carcinoid primary tumours. These primary tumours displayed shattering events on *chr 2*, *3*, *6*, *7*, *9* and *11*, and thus demonstrate the possibility of NEC development through transformation of a lung NET via chromothripsis. In light of these cases, we report here upon a supra-carcinoid patient for which a patient-derived tumour organoid (PDTO) was created^59^. LNET10, a female never-smoker in the age range 16-40, was diagnosed with an atypical carcinoid at stage IA (T1bN0M0), with two mitoses/2mm^2^, focal necrosis, and Ki-67 index of 20% (**Fig. 5c, top panel**). Molecular analysis of both the primary tissue and PDTO passage 4 (p4) identified moderately high *MKI67* expression (5 and 14 TPM, respectively), *BRAF* (V600E) and *PTEN* mutations, expression-based *c*lustering with LCNEC^59^, and a chromothripsis-like clustered SV pattern affecting *chr 9* similar to the aforementioned NETs with SCLC metastases^81^ (**Fig. 5c, middle panel, Supplementary Tables S1, S4, S15 and S16**). One year after diagnosis, metastatic disease in the skin and lymph nodes was noted, and the patient was then treated with BRAF and MEK inhibitors, dabrafenib and trametinib, respectively. After an initial response, bone and liver metastases developed, and biopsy of the liver metastasis followed by WGS revealed a deletion in exons 2–8 of *BRAF*, a known mechanism of resistance to BRAF inhibitor therapy, that was absent in the p4 PDTO. Crucially, based on the liver biopsy, the pathologist revised the diagnosis to LCNEC, suggesting that a full transformation to NEC may have occurred. This case report provides additional evidence that a low-grade NET may progress to a high-grade NEC in the context of chromothripsis, and suggests the previously identified atypical SCLC^81^ may in fact be fully transformed supra-carcinoids.

The PDTO was one of the fastest-growing lung NET PDTOs, ~three months to passage three versus on average six months for the others, and displayed an increase in mutational burden and *MKI67* gene expression over time, from the primary to passage 11 (p11, **Fig. 5c, bottom panel**), thus reflecting the tumour aggressiveness seen in the patient. We subsequently assessed the cell type make-up of the primary and PDTO passages, finding that while both the primary and p4 PDTO contained LAP cells, a unique feature of supra-carcinoids (**Fig. 4a,b**), these were absent in p11 (**Fig. 5c, bottom panel**). There was also an observable shift from a NET-like to NEC-like profile over time, despite the lack of TME in culture (**Fig. 5c, bottom panel**), mirroring the patient’s progression. Although further experiments are warranted, our observations from spatial transcriptomics and organoid culture point to supra-carcinoids, promoted by the presence of LAP cells within a specific TME, being a subset of lung NETs with the capacity to progress to lung NECs, and that their aggressive profile, once acquired, may be independent of the TME.

## Conclusion and perspectives

The 2021 WHO Classification of Thoracic Tumours categorises lung NETs into grade-1 typical carcinoids and grade-2 atypical carcinoids. However, this classification has two major limitations. First, it does not account for newly emerging biological and morphological entities with more aggressive behaviour, such as supra-carcinoids and grade-3 lung NETs. Second, while tumour histology remains the gold standard for diagnosis and clinical decision-making, grading alone is insufficient for therapy selection and relapse prediction. This may be attributed to the fact that, although the WHO Classification of Tumours has progressively integrated molecular data for many cancers, the classification of lung NETs has remained largely unchanged for decades. The data from the lungNENomics project shows a partial overlap between molecular and histological classifications suggesting that each captures different aspects of the disease, highlighting their complementary clinical value (**Fig. 6**). These new data present a unique opportunity to refine the current morphological classification by incorporating new criteria that more accurately reflect tumour biology and clinical behaviour. Finally, combining our findings with recent data from Rekhtman and colleagues^81^ suggests that the aggressiveness of supra-carcinoids may be driven by a distinct undifferentiated cellular state that remains undetected within an apparently low-grade tumour until a specific trigger initiates its progression. This raises the question of whether supra-carcinoids represent a crucial intermediate stage in the multi-step carcinogenesis process leading to the development of a subset of neuroendocrine carcinomas, such as the carcinoid-like SCLC in never-smokers. This possibility warrants further investigation through new single-cell and spatial technologies and the use of longitudinal samples, allowing for deeper characterisation of this population of cells and their interactions with the TME.

**Figure 6.**
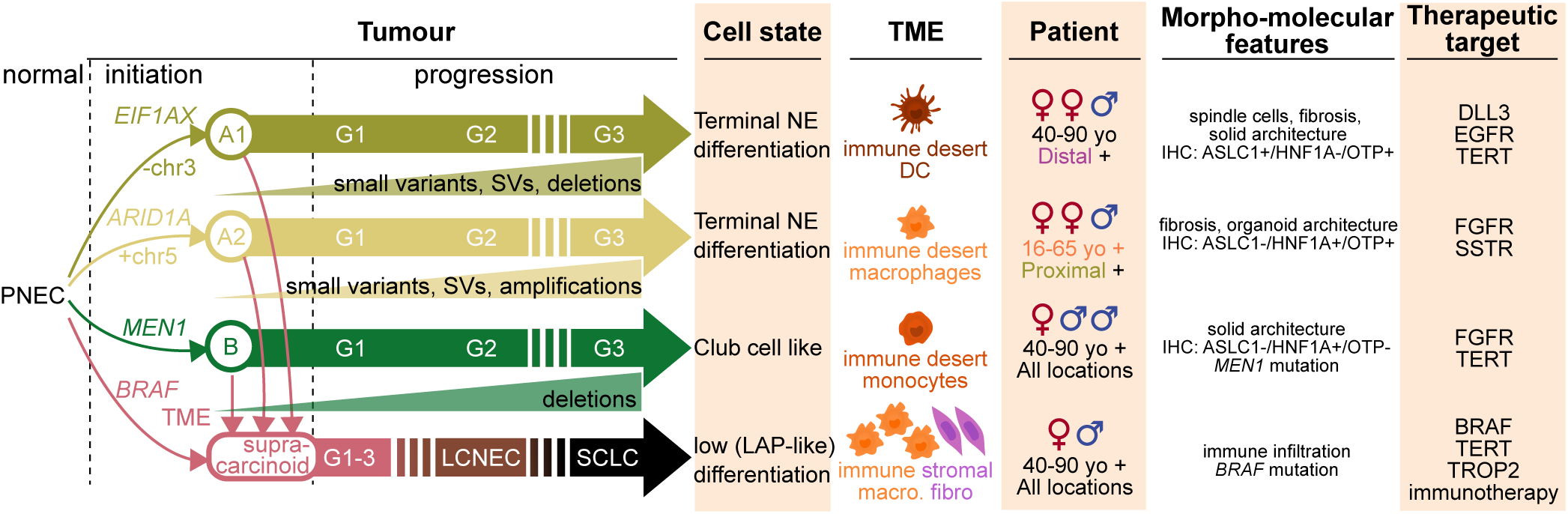
A morpho-molecular classification for lung NETs. Schematic depicting the molecular, clinical, and morphological characteristics of the proposed morpho-molecular classification for lung NETs.

## Additional content

Methods, Main and Extended Data Figures, Supplementary Figures, and Supplementary Tables are available in separate files.

## Supporting information

Methods_11072025_medrxiv.pdf

## Methods

See attached document entitled Methods_11072025_medrxiv.pdf

## Data availability

Sequencing data are available on the European Genome-Phenome archive (study EGAS00001005979). Previously published data from Alcala *et al*.^9^ and Dayton *et al*.^59^ are also available on EGA (studies EGAS00001003699 and EGAS00001005752, respectively).

## Code availability

The code used are available on IARC’s github repository https://github.com/IARCbioinfo/ under open-source licenses. In particular, the code to process the sequencing data (QC, alignment, variant calling) are listed at https://github.com/IARCbioinfo/IARC-nf, the code to process the whole-slide images are available at https://github.com/IARCbioinfo/WSIPreprocessing and ­and the code to analyse the data and reproduce the main results (Figures) is available on the github repository https://github.com/IARCbioinfo/MS_lungNENomics.

## Acknowledgements

The lungNENomics project is part of the Rare Cancers Genomics initiative (www.rarecancersgenomics.com/) led by the Computational Cancer Genomics team at the IARC (https://www.iarc.who.int/teams-ccg/). This work is also part of the European Neuroendocrine Tumor Society (ENETs) lung task force. We thank the Hospices Civils de Lyon (CRB-HCL BB-0033-00046) and Centre Léon Bérard (CRB-CLB BB-0033-00050) biobanks in Lyon, as well as Côte d’Azur University’s biobank (BB-0033-00025) in Nice, France, all authorised by the French Ministry of Research, for sharing human biological samples and associated data. We thank the 12 centres that voluntarily participated in the lungNENomics project by providing one FFPE block per patient: The Tumour Bank of the François Baclesse Centre in Caen (France), the Institut für Diagnostik und Forschung in Pathologie of the Medical University of Graz (Austria), the Department of Biopathology of the Léon Bérard Centre in Lyon (France), the Institute of Pathology of the Hospices civils de Lyon (France), the Department of Surgical Oncology of St Vincent’s Hospital in Melbourne (Australia), the Department of Oncology and Haemato-Oncology of the University of Milan (Italy), the Department of Biopathology at the Nancy Regional Hospital (France), the Laboratory of Clinical and Experimental Pathology at the Pasteur Hospital in Nice (France), the Departments of Pathology and Oncology at Oslo University Hospital (Norway), the Pathology Department at the Cochin Hospital in Paris (France), the Oncology Unit at the IRCCS Cas Sollievo della Sofferenza Foundation in Rotondo (Italy), and the Oncology Department at the University of Turin (Italy). The results published here are in part based upon data generated by the TCGA Research Network: https://www.cancer.gov/tcga. We also would like to acknowledge the contribution of Jean-Philippe Berthet, Frederic Bibeau, Cécile Blanc Fournier, Anne Boland, Christelle Bonnetaud, Charlotte Cohen, Concetta Martina Di Micco, Jean Philippe Lerochais, Nathalie Rousseau, and Giovanna Sabella.

## Author contributions (CRediT)

Conceptualization: N.A., M.F., L.F-C.

Data curation: A.S-O., E.M., N.C., C.V., A.DG., N.A.

Formal analyses: A.S-O., E.M., N.C., Y.L., C.V., A.DG., La.Ma., Z.L., R.B-E., A.G-P., M.L.M., E.L., Li.M., Li.B.,V.C., N.A.

Funding acquisition: N.A., M.F., L.F-C.

Investigation: T.vW., L.M.H., La.Mo., J.A., C.D., A.C., Cy.C., F.D., N.G., A.G., C.P., S.T-E., La.C., M.V., J-M.V.,E.C., A.M-L., Lu.B., G.P., M.G.P., S.L.

Methodology: E.M., N.A.

Project administration: A.S-O., M.F., L.F-C.

Software: A.S-O., E.M., N.C., Y.L., A.DG., La.Ma., Z.L., N.A.

Resources: G.A., O.T.B., G.C., V.T.dM., J-F.D., A-M.C.D., E.F., P.G., P.H., V.H., S.L., M.L-I., M.M., L.A.M., C.P., G.P.,

H.P., L.R., A.S., W.B., J.vdB., M.T., E.J.M.S., S.T-E., T.W., G.M.W., La.C., C.C., M.V., J-M.V., Lu.B., M.G.P., S.L.

Supervision: A.I-C., N.L-B., S.P-A., J.K., E.J.M.S., S.T-E., J.P.C., Li.C., N.G., S.L., J.D., T.D., N.A., M.F., L.F-C.

Validation: A.S-O., E.M., N.C., N.A.

Visualization: A.S-O., E.M., Y.L., La.Ma., N.A.

Writing – original draft: A.S-O., E.M., N.C., Y.L., La.Ma., N.A., M.F., L.F-C.

Writing – review & editing: all authors

## Additional information

### Supplementary Figures

See attached document entitled Suppelementary_Figures.pdf

### Supplementary Tables

See attached document entitled Index_of_Supplementary_Tables.pdf and Excel files

## Extended Data Figures

**Extended Data Figure 1.**
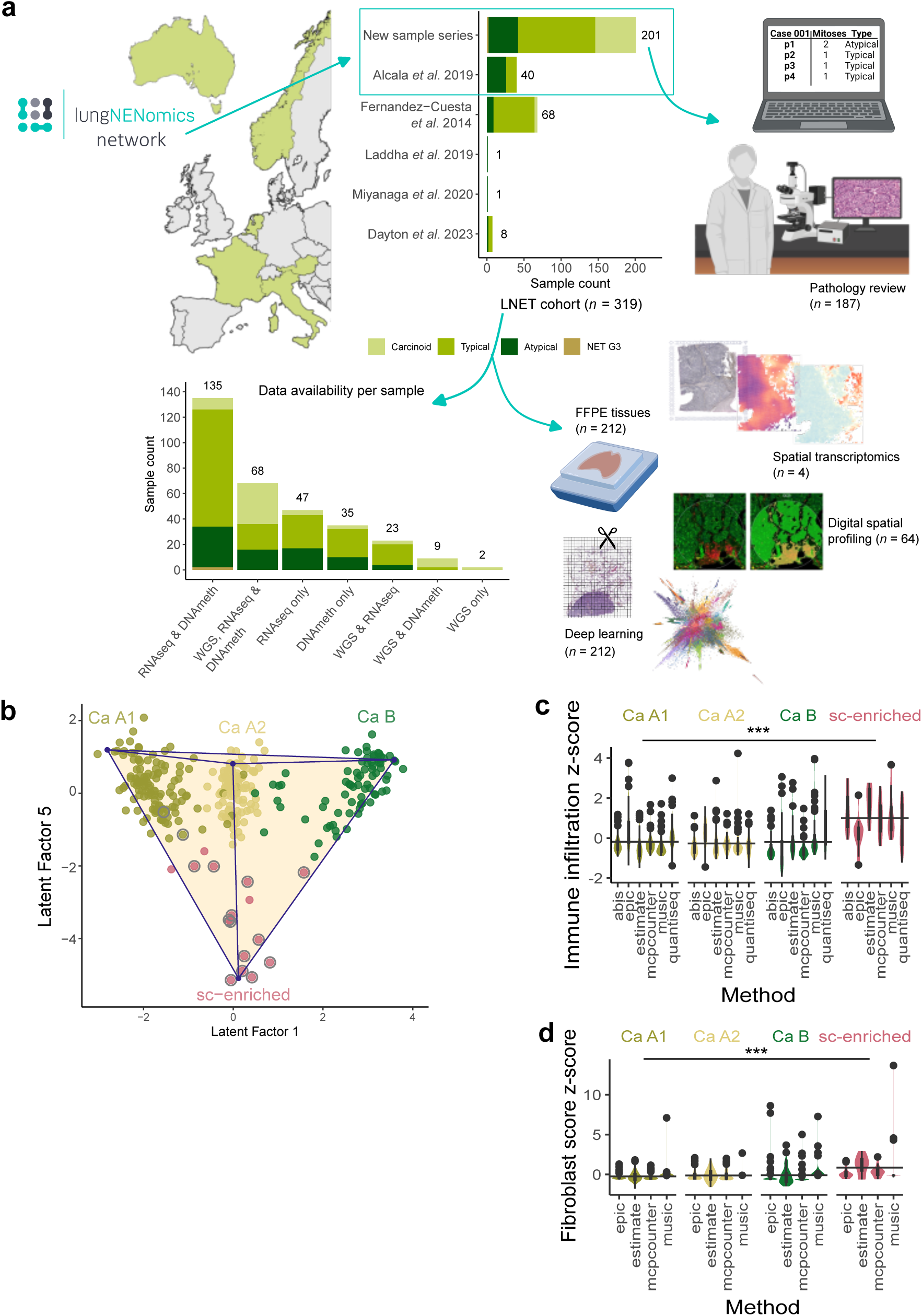
Study design overview and additional features of lung NET molecular groups. **a**, Origin of samples within the lung NET cohort (*n* = 319, lungNENomics network or previous publications), and number of samples per histological type by origin (where “carcinoid” indicates tumours that could not be definitively classified as typical or atypical). Depiction of central pathology review. Distribution of omics data generated, or publicly available, by sample type for the lung NET cohort. Analysis techniques performed with formalin-fixed paraffin-embedded (FFPE) tissues from the lungNENomics network series. Figure created, in part, with BioRender.com. **b**, Data as in Fig. 1a plotted over MOFA Latent Factors 1 and 5. **c**, Infiltrating immune cell scores from six estimation methods per molecular group (*n* = 273). Mean z-scores per molecular group were compared by t-tests (linear mixed model with methods as random effects). **d**, Distribution of fibroblasts score z-scores from six estimation methods for each molecular group (*n* = 273). Mean z-scores per molecular group were compared by t-tests. *** *P* value *≤* 0.001.

**Extended Data Figure 2.**
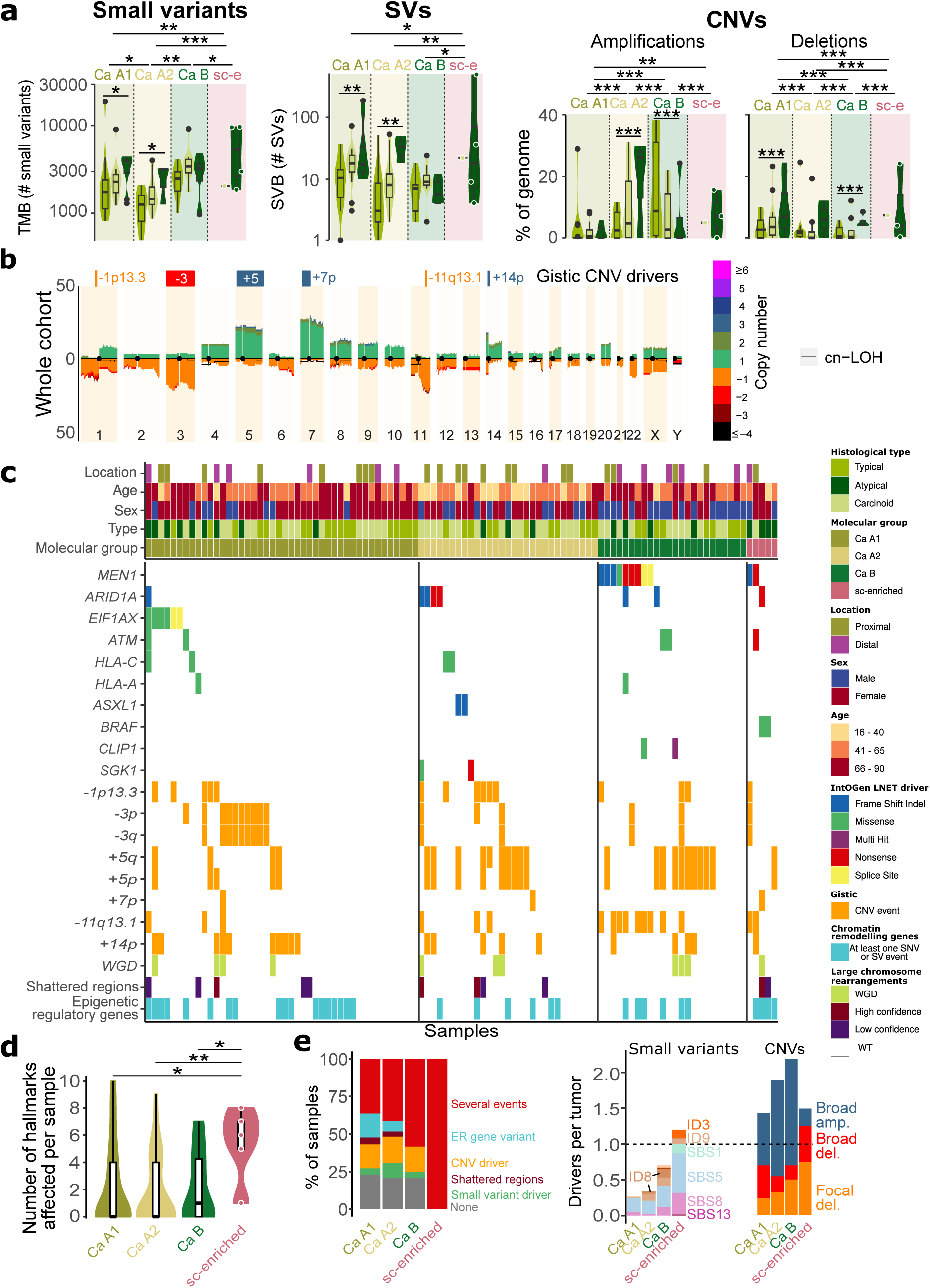
Genomic profile of lung NET molecular groups. **a**, Molecular group and histological type mutational burdens for different types of alterations. Stars represent the significance level of two-sided t-tests (small variants, structural variants, SVs), or permutation tests (copy number variations, CNVs). **b**, Copy number profile of the lung NET cohort; the y-axis represents the percentage of samples in each group with a given alteration (amplifications above the 0 line, deletions below), and colours represent the copy number. The black line represents copy-neutral loss of heterozygosity (cn-LOH). Driver CNVs detected in each group by the GISTIC2 method are represented by a square above each profile (red and orange rectangles for deletions, blue rectangles for amplifications). **c**, Oncoplot of small variants in IntOGen identified driver genes, CNV drivers identified by GISTIC2, and large chromosomal rearrangements. Selected clinical features and molecular group assignment are represented at the top along the x-axis. **d**, Distribution of the number of hallmarks affected by small and structural variants per patient for each group. Stars represent the significance level of Mann-Whitney U Tests. **e**, Percentage of samples with genomic drivers of different types in each molecular group (left), average number of drivers of different types in each molecular group (right). SBS1/5/8/13 and ID3/8/9 refer to single base substitution and indel signatures, respectively. amp, amplification; del, deletion. * 0.01 < *P* value < 0.05; ** 0.001 < *P* value *≤* 0.01; *** *P* value *≤* 0.001.

**Extended Data Figure 3.**
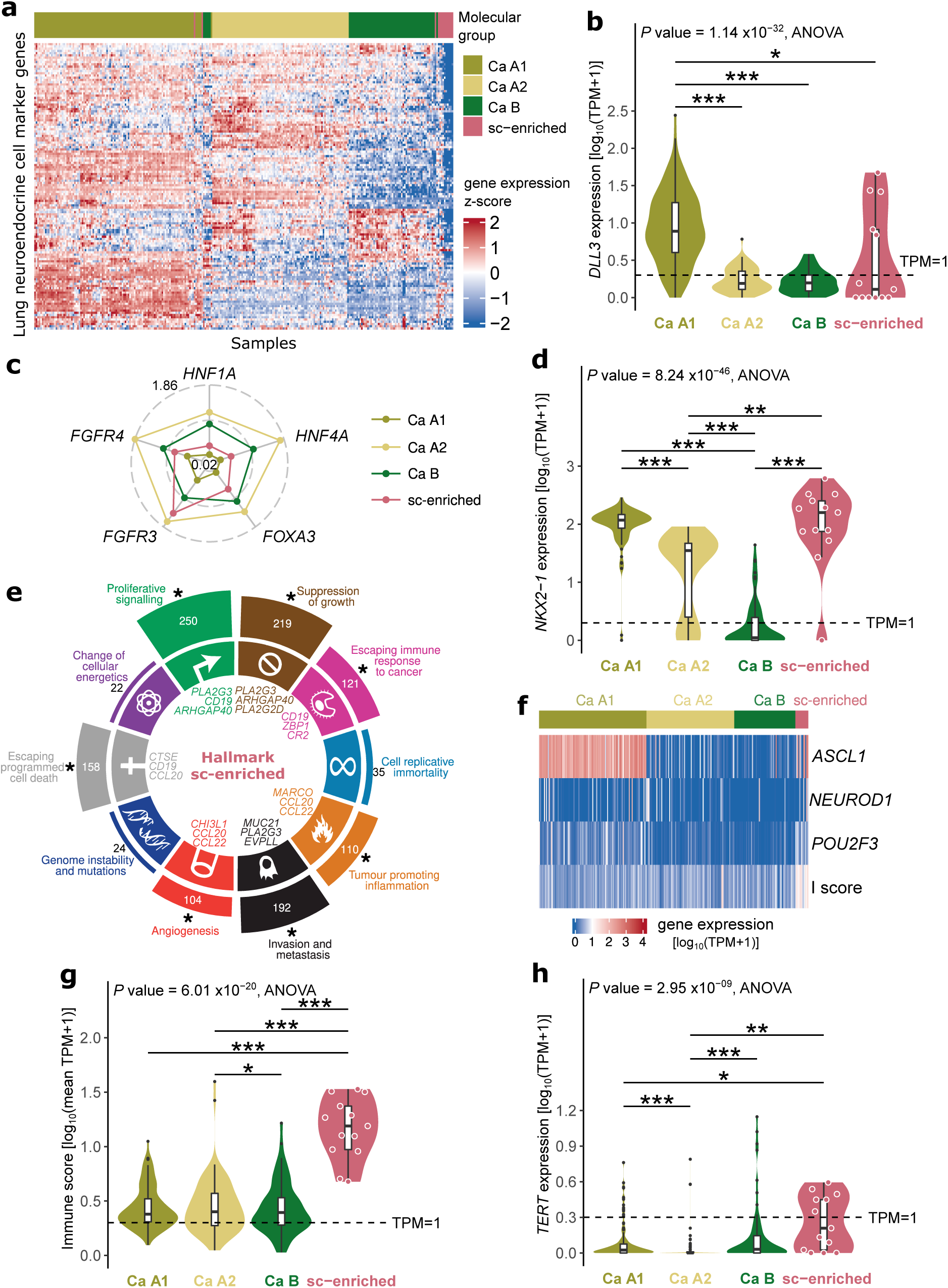
Transcriptomic profiles of lung NET molecular groups. **a**, Gene expression values of lung NET samples (*n* = 273), plotted as z-score of variance stabilised transformed expression values, for 128 genes highly expressed in lung neuroendocrine cells. **b**, Distribution of *DLL3* expression by molecular group. TPM, transcripts per kilobase million. **c**, Radar plot of log10(TPM+1) values of genes associated with the HNF+ subtype, values range from 0.02 to 1.86. **d**, Distribution of *NKX2-1* expression by molecular group. **e**, Polar bar plot of the number of sc-enriched core upregulated genes per feature. Asterisks indicate that the corresponding feature is significantly enriched in sc-enriched core upregulated genes. The three genes per hallmark with the largest fold change in sc-enriched are displayed. **f**, Heatmap of sample gene expression of markers of small cell lung cancer molecular groups, in log10(TPM+1) units, coloured bars along the top represent sample molecular group. Immune score (I score) is the log10(mean(TPM+1)) of selected immune genes. **g**, Distribution of the I score by molecular group. **h**, Distribution of *TERT* expression by molecular group. Pair-wise comparisons performed with t-tests. * 0.01 < *P* value < 0.05; ** 0.001 < *P* value *≤* 0.01; *** *P* value *≤* 0.001.

**Extended Data Figure 4.**
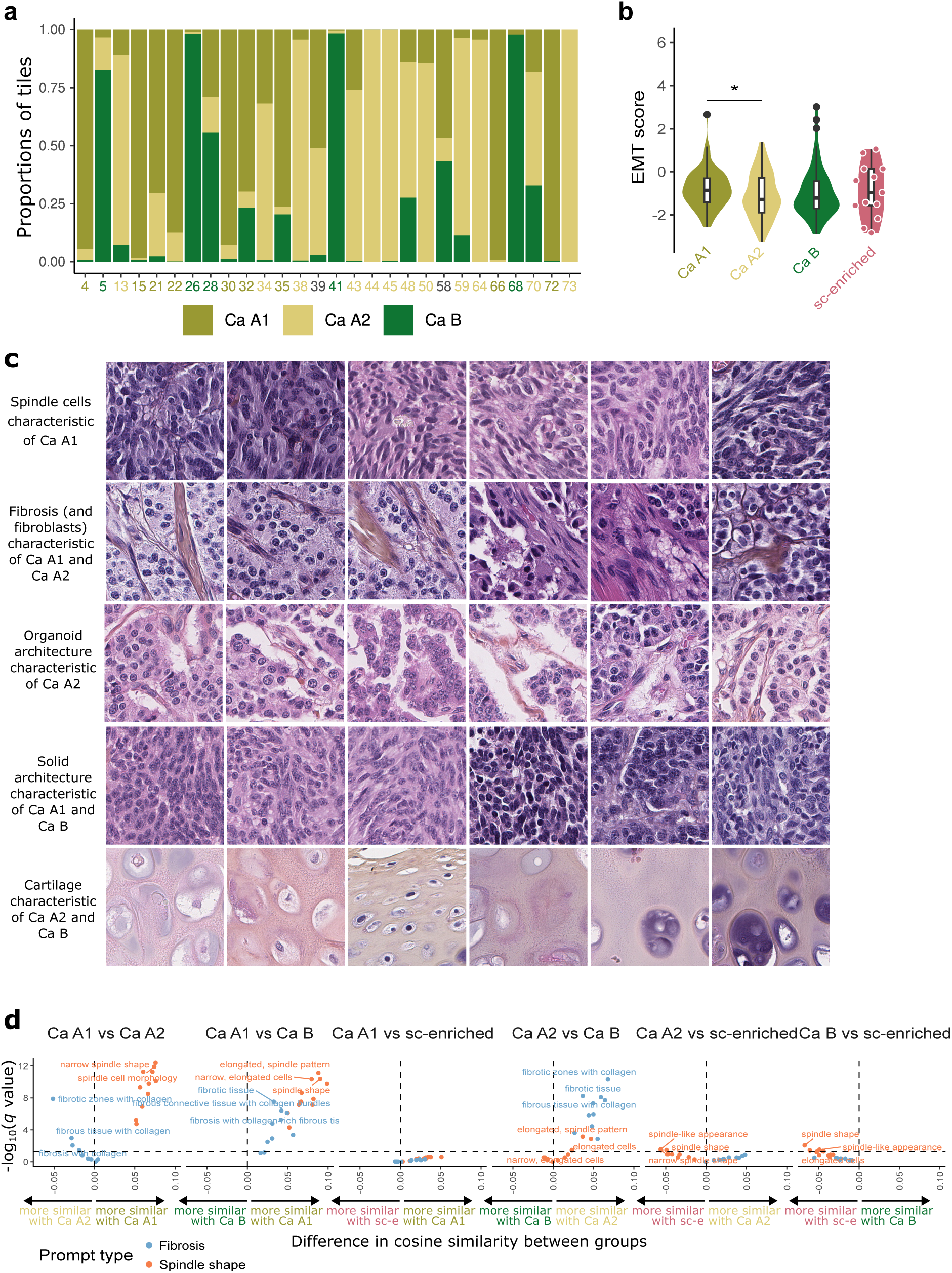
Deep learning analyses of whole-slide images. **a**, Proportion of tiles from each molecular group within the final 28 partitions defined by the supervised branch and interpreted by the panel of pathologists. Partition numbers are coloured by their most frequent molecular group. **b**, Epithelial-Mesenchymal Transition (EMT) score, computed from marker gene expression. Pair-wise comparisons performed with t-tests. * 0.01 < *P* value < 0.05. **c**, Representative tiles of five key features, illustrating the morphological characteristics of the molecular groups. Each tile was independently reviewed by two pathologists, and only tiles where the feature was reported by both pathologists are shown. **d**, Robustness analysis of the results presented in Fig. 3c, using alternative prompts. Volcano plots represent the statistical significance (*q* value, y-axis) versus the effect size (x-axis) of the differences between molecular groups in terms of cosine similarity between CONCH WSI tile embeddings and text prompt embeddings, for different prompts (points).

**Extended Data Figure 5.**
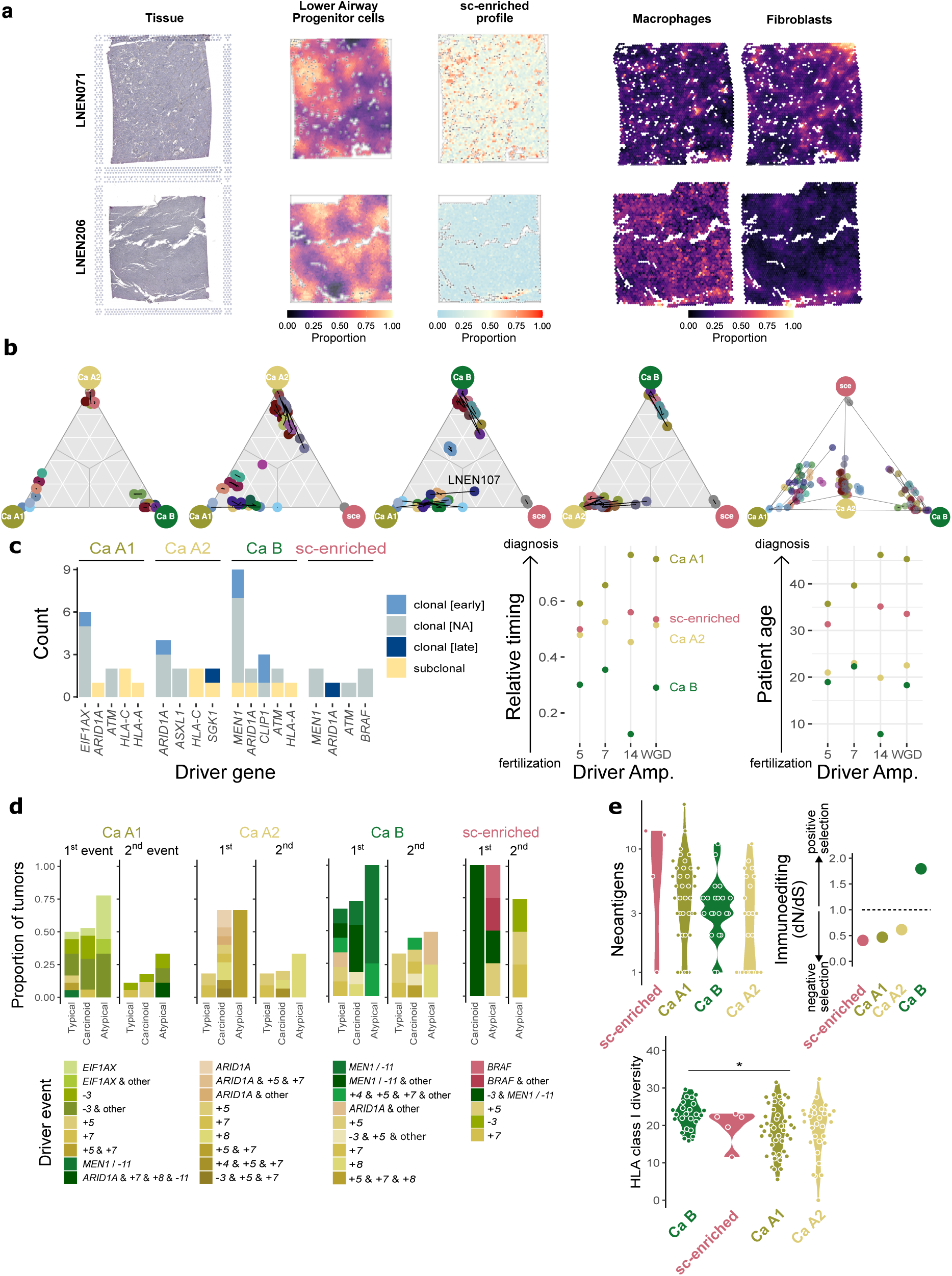
The evolution of lung NET groups. **a**, Figure design as in Fig. 4c, for samples LNEN071 and LNEN206. **b**, Ternary plots depicting the relative position of intra-tumoural heterogeneity (ITH) samples in space between different molecular groups. Black lines join ITH samples from the same patients. ITH samples are coloured by patient. Patient LNEN107 highlighted as the patient whose ITH samples were assigned to two different molecular groups. **c**, Clonality (colour) of driver alterations identified in each molecular group (left), and inferred timing of driver amplifications in timing relative to the number of mutations in the chromosome segment (0: amplification happened before all mutations, 1: amplification happened after all mutations, middle), and chronological age (in years, right) scales. WGD, whole-genome doubling. **d**, Recurrent evolutionary trajectories based on small variants and CNV drivers detected using the REVOLVER method. Colours represent the driver event, “1st event” refers to initiating events (trajectories from germline to a driver in Fig. 4e), and “2nd event” refers to subsequent events (trajectories between two drivers in Fig. 4e). **e**, Tumour-Immune genomic interactions. Number of neoantigens per molecular group (top left), level of immunoediting, estimated as the selection coefficient of small variants in neoantigen-rich regions (ratio of non-synonymous to synonymous mutations, dN/dS) by the SOPRANO method (top right), genetic diversity score (Grantham 1974’s amino acid score using exons 2 and 3) of germline HLA class I alleles in patients who developed lung NETs of different molecular groups (bottom). Stars represent the significance levels of t-tests. * 0.01 < *P* value < 0.05.

