## Supplementary Fig. for "A clinically relevant morpho-molecular classification of lung neuroendocrine tumours"

### Supplementary Figures for article “A clinically relevant morpho-molecular classification of lung neuroendocrine tumours"

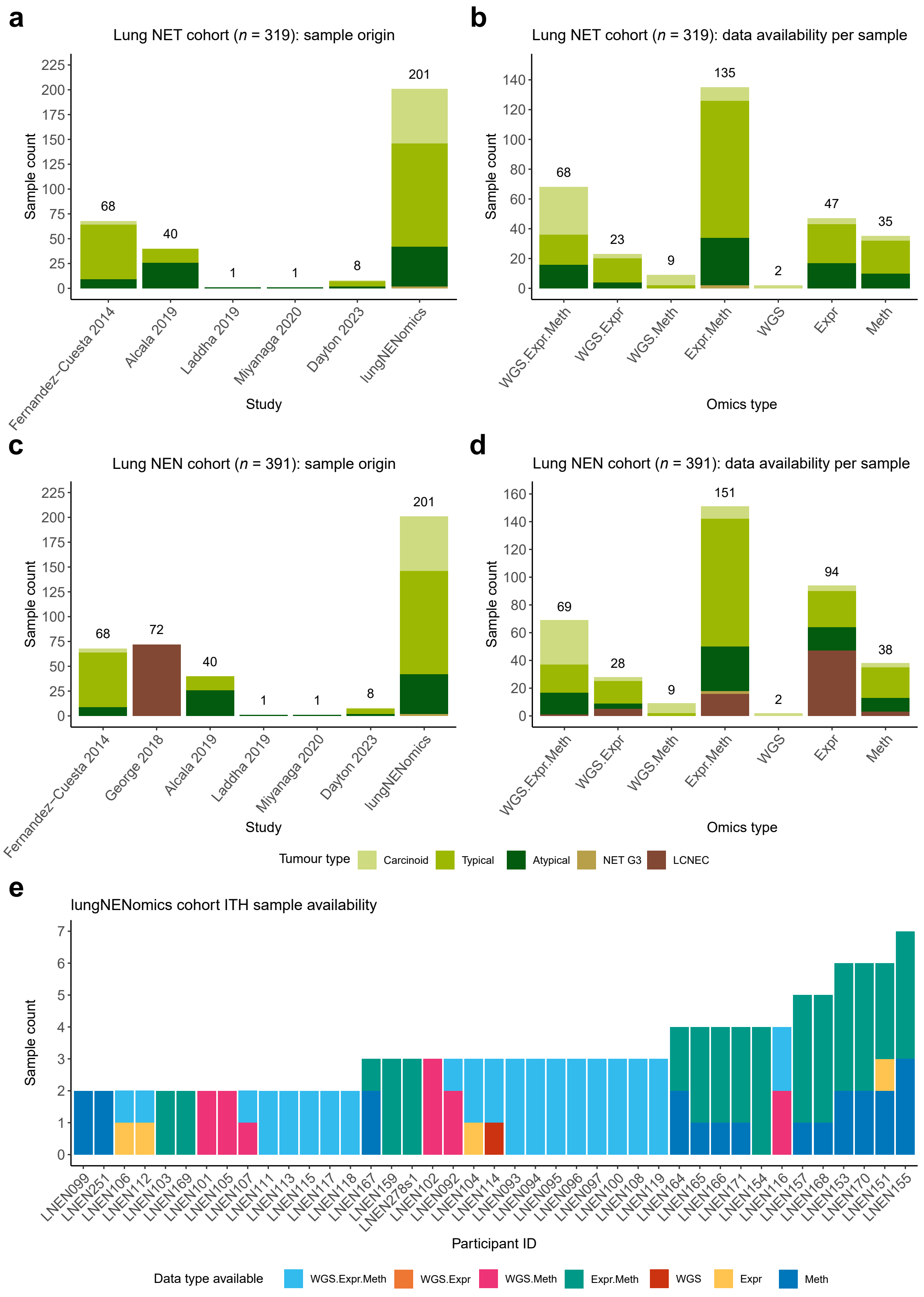

**Supplementary Figure S1. Overview of lung NET and lung NEN cohorts. a**, Distribution of histological types per study or cohort of origin in the lung neuroendocrine tumour (lung NET) cohort (*n* = 319). **b**, Distribution of histological type by molecular data types available in the lung NET cohort. **c**-**d**, as per (**a**-**b**) for the lung neuroendocrine neoplasm (lung NEN) cohort (*n* = 392). **e**, data types available for each sample of the lungNENomics intra-tumoural heterogeneity cohort (ITH), for which multiple regions were sequenced (*n* = 41 patients, *n* = 138 samples).

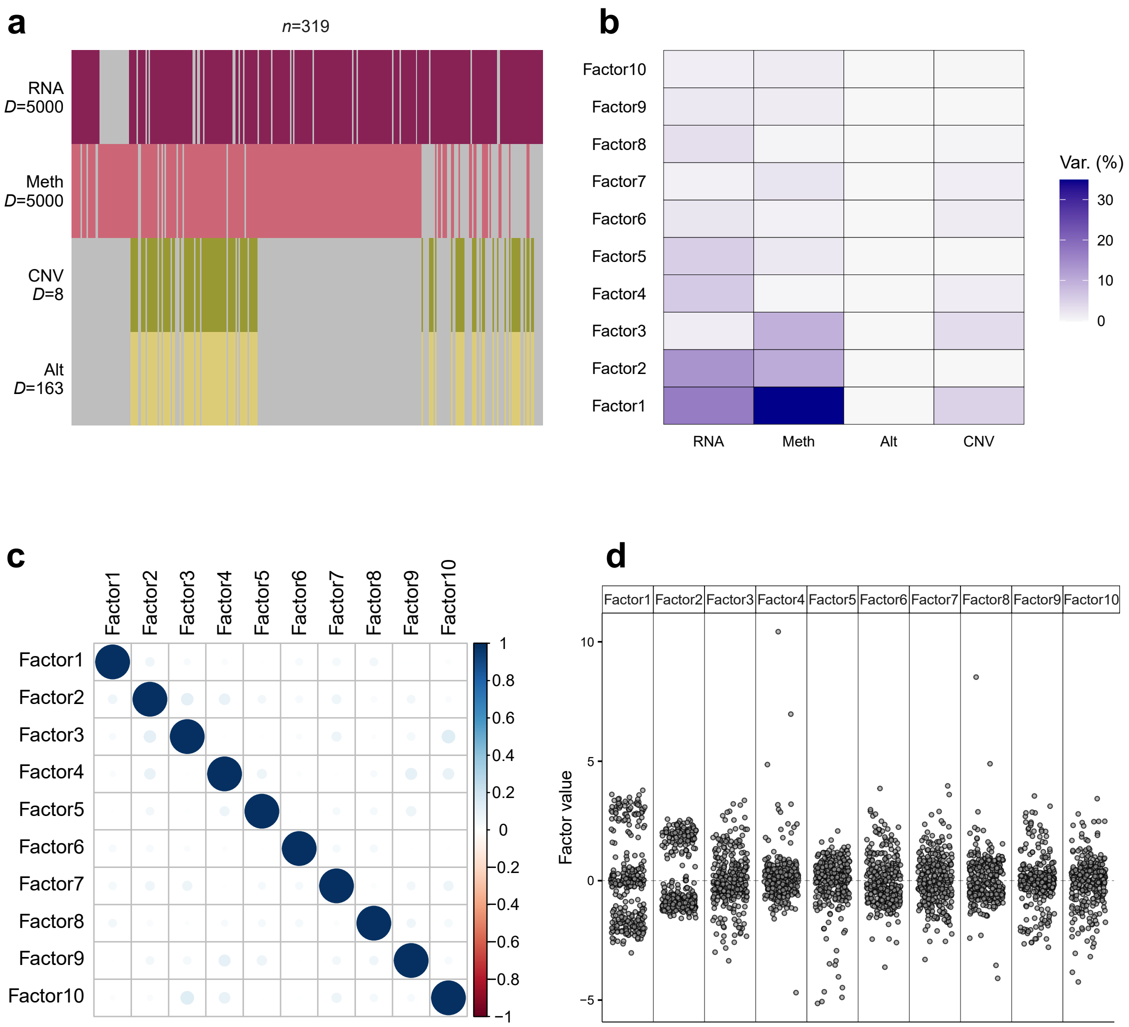

**Supplementary Figure S2. Overview of inputs and outputs of lung NET cohort MOFA. a**, Input data matrices. D is the number of omic features incorporated per matrix [expression levels for 5000 genes (RNA), DNA methylation levels at 5000 CpG sites (Meth), copy number values at eight broad and focal regions (CNV), small and/or structural variant status at 163 genes (Alt)]. Grey indicates missing data. **b**, Variance explained by each latent factor for each data type. **c**, Correlation between sample coordinates along latent factors. **d**, Distribution of samples along each latent factor.

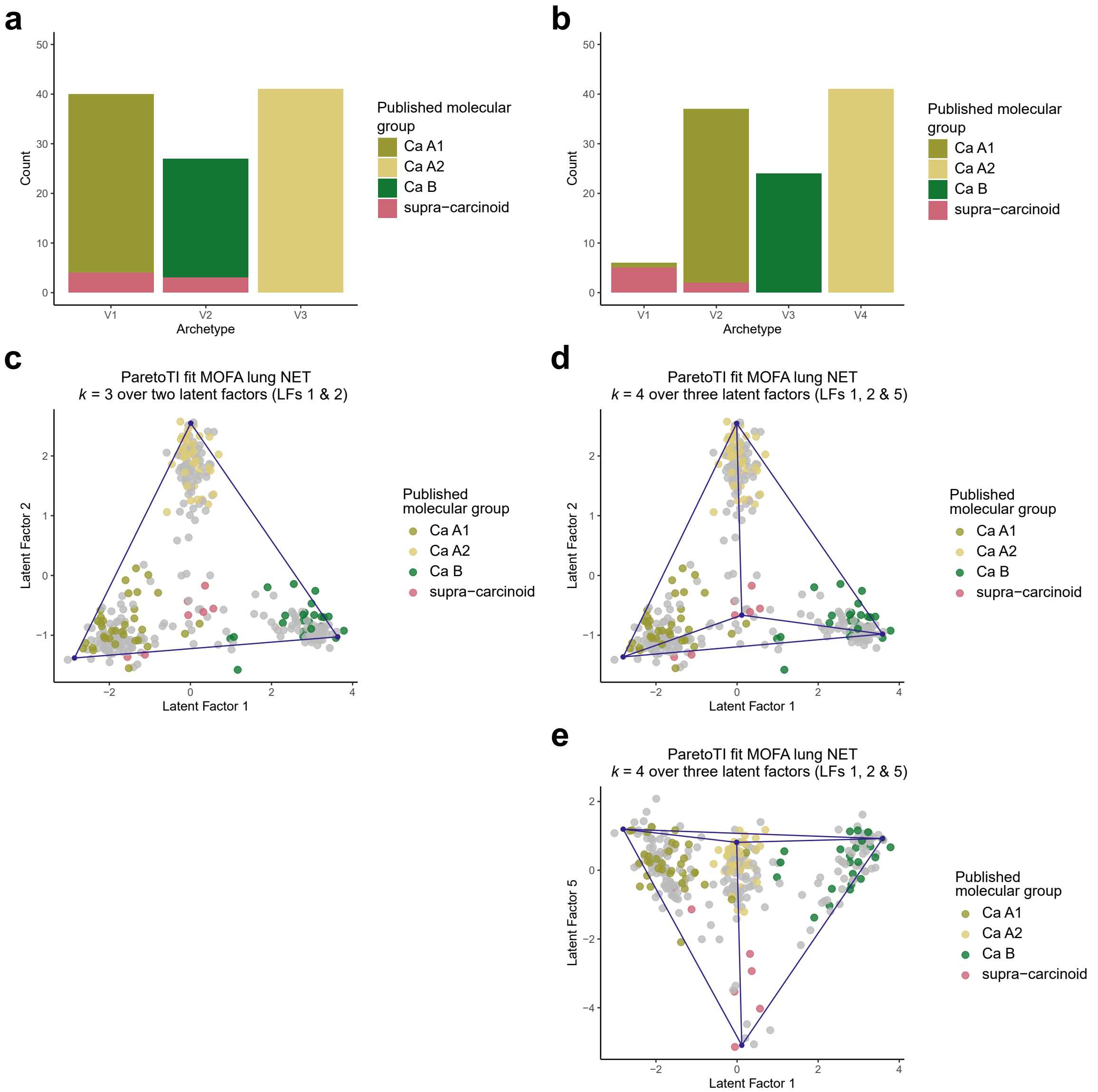

**Supplementary Figure S3. Correspondence between ParetoTI archetypes identified in the lung NET cohort and previously published molecular groups. a–b**, Count of samples from Alcala *et al.* 2019 ($n$ = 107) and Dayton *et al.* 2023 ($n$ = 1) coloured by published molecular group within archetypes found in the current study when considering $k$ = 3 (**a**) and $k$ = 4 (**b**) archetypes. **c–e**, Scatterplots showing the coordinates of samples from Alcala *et al.* 2019 and Dayton *et al.* 2023 (coloured by previously published molecular group), remaining lung NET cohort samples ($n$ = 211, grey points), and archetype proportions (blue points) along latent factors for (**c**) $k$ = 3 archetypes derived from latent factors (LFs) 1 and 2, and (**d–e**) $k$ = 4 archetypes derived from LFs 1, 2 and 5.

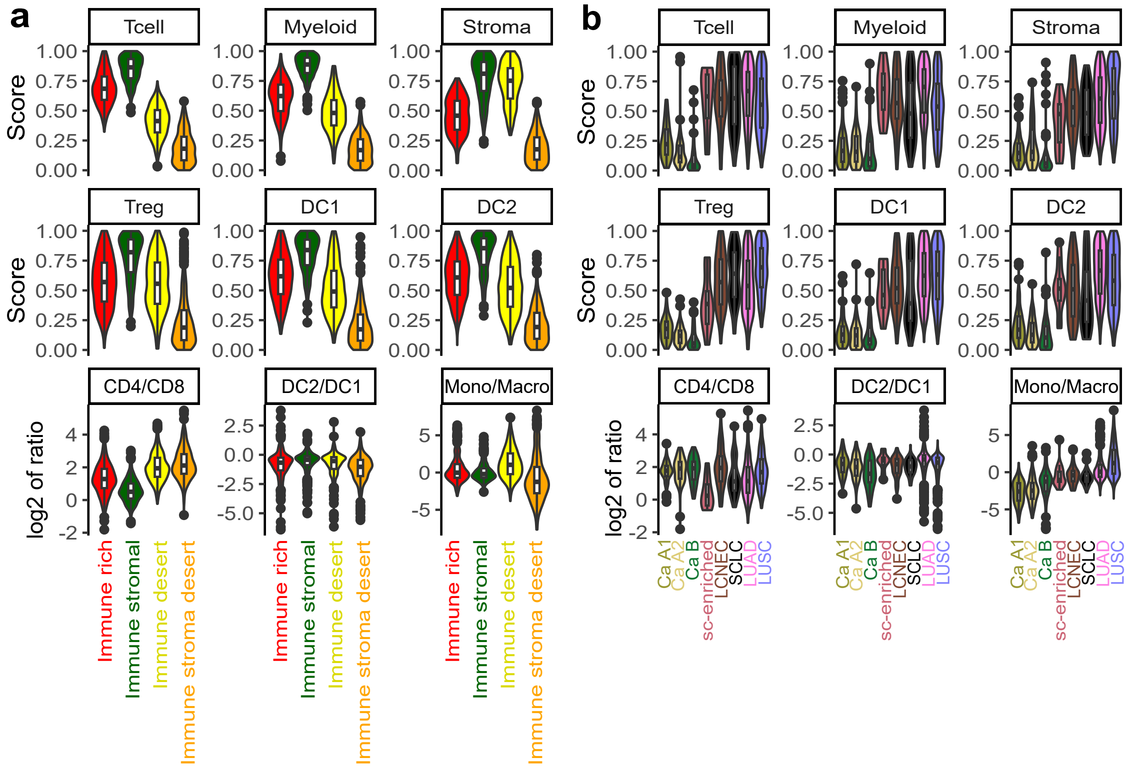

**Supplementary Figure S4. Identification of coarse immune archetypes in lung neoplasms using the three features from Combes *et al.* 2022. a**, Violin plots of each of the three features used for the classification (T cell, Myeloid, and Stroma), in units of percentile rank of expression among samples, across the four immune clusters we identified. **b**, Same as (**a**) but comparing lung NET molecular groups and lung carcinoma types. In (**a**) and (**b**) samples correspond to cohort lung NEN plus SCLC, and LUAD (lung adenocarcinoma) and LUSC (lung squamous cell carcinoma) TCGA datasets ($n$ = 1420).

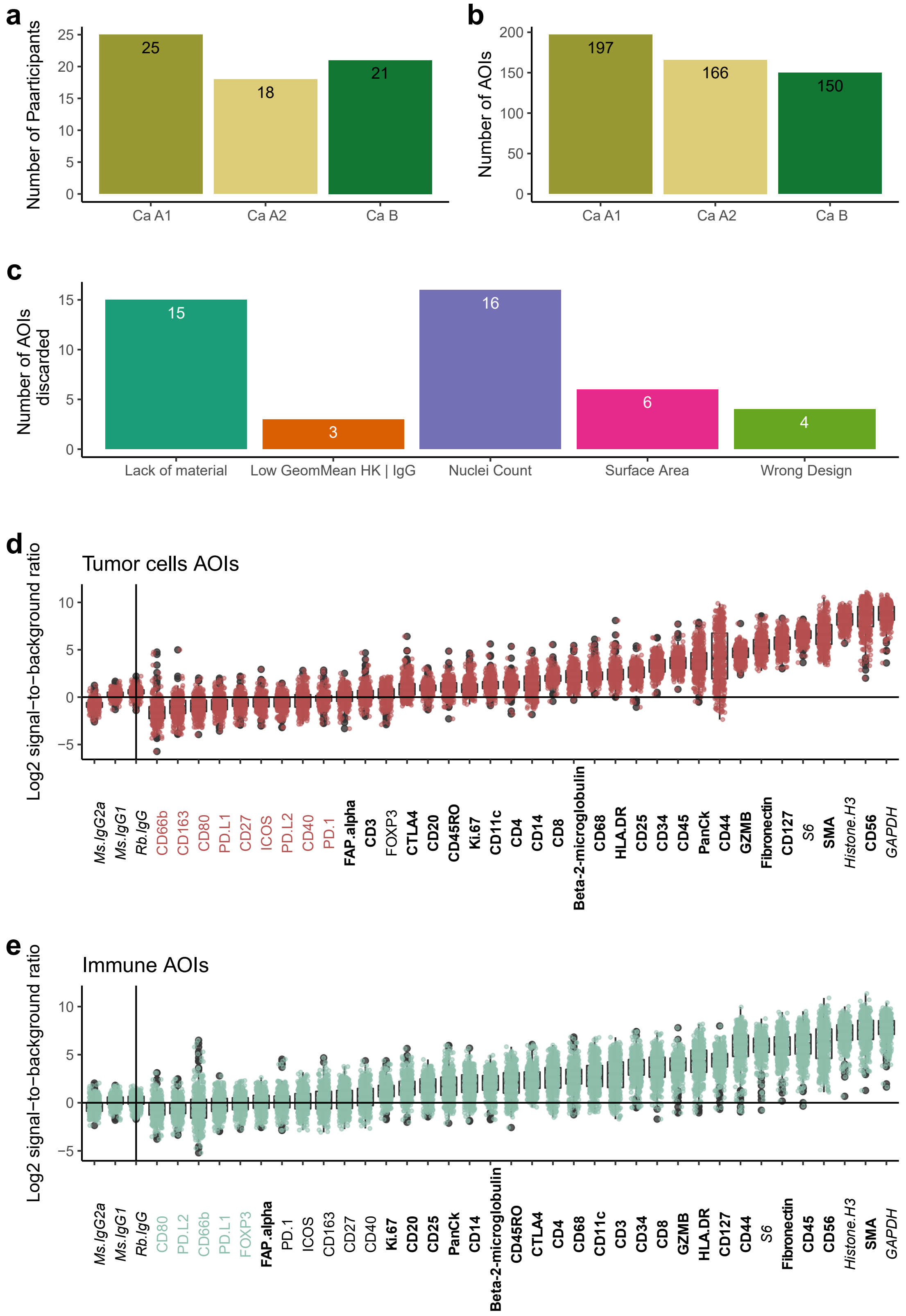

**Supplementary Figure S5. Data description and quality control of the Digital Spatial Profiling (DSP) experiment. a**, Number of patients included in the DSP experiment per molecular group. **b**, Number of areas of interest (AOIs) per molecular group selected for the DSP experiment. **c**, Quality control of the AOIs and reasons for exclusion for 44 AOIs: lack of collected material, insufficient expression (geometric mean, Low GeomMean) of housekeeper (HK) probes S6, Histone H3, and GAPDH or negative control probes (IgG) Rb IgG or Ms IgG2, low number of nuclei (i.e., less than 20), low surface area (i.e., less than 1600 μm2), or wrong (mixed tumour/immune) design. **d**, Log2 signal to background ratio distribution (y-axis) of profiled proteins (x-axis) within the tumour cell AOIs. The background was estimated as the geometric mean of the negative control probes. The signal from the IgG probes is indicated to the left of the vertical bar. The name of the protein in colour corresponds to the protein that was excluded from the analysis according to the following criteria: mean log2 signal to background ratio less than 0. Probes in bold were included in subsequent analysis, probes in italics are control probes. **e**, as per (**d**) for immune cell AOIs.

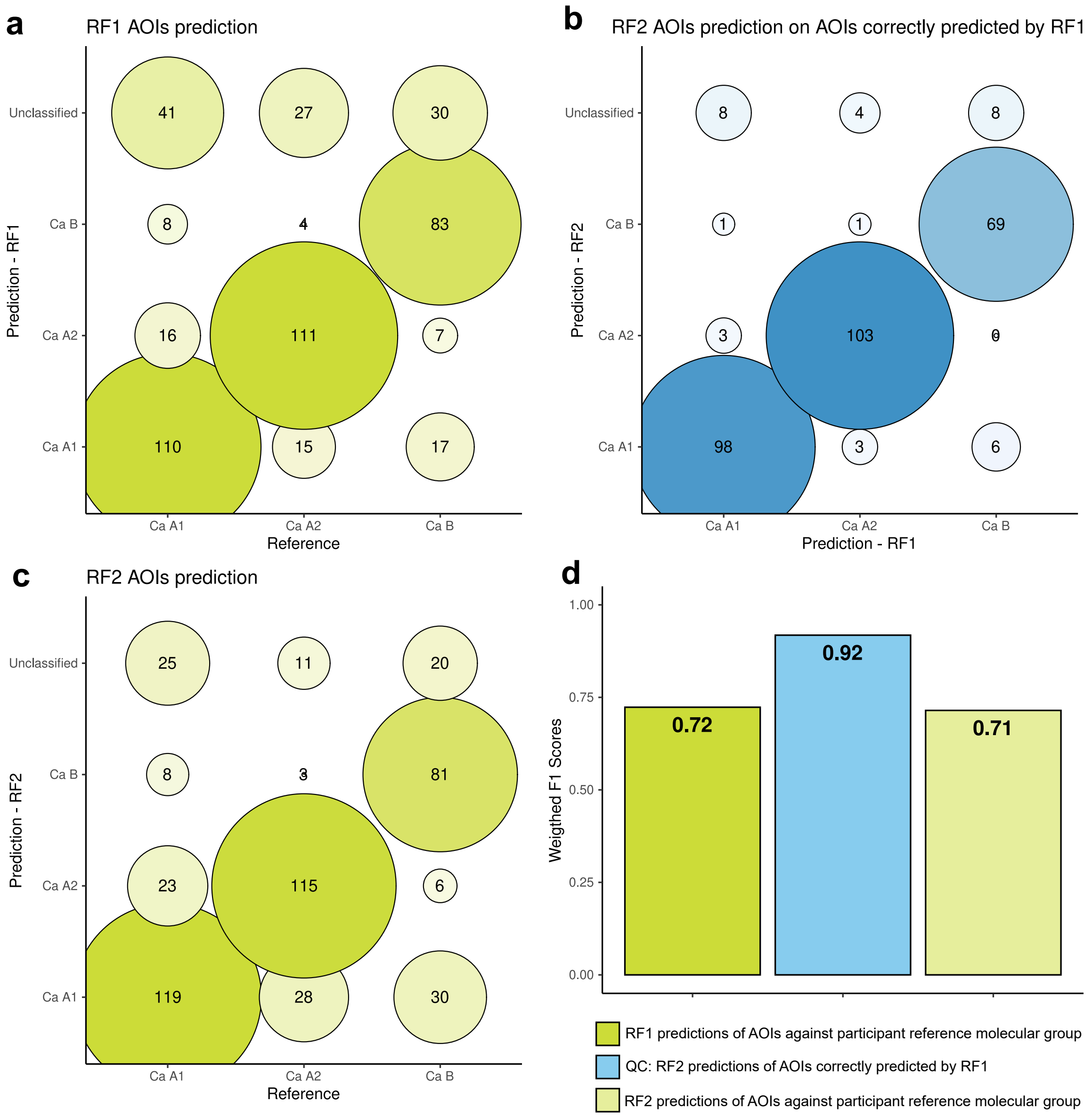

**Supplementary Figure S6. Performance of random forest predictions from DSP data.** **a**, Confusion matrix of molecular group predictions from digital spatial profiling (DSP, y-axis) against ground truth from tumour bulk multi-omic analyses (x-axis) of the AOIs selected for DSP, by a first random forest model, RF1 trained on all AOIs using five-fold cross-validation. Note that discrepancies between reference and predictions could either be classification errors from the model or genuine biological differences between an AOI's group and the patient level group. **b**, Confusion matrix of molecular group predictions of AOIs for which the predictions of RF1 matched the molecular group defined at the participant level (*n* = 304 AOIs on the diagonal in panel **a**). These predictions result from a second random forest model, called RF2, trained only on AOIs correctly predicted by RF1 in order to reduce training data noise due to intra-tumour heterogeneity. **c**, Confusion matrix of molecular group predictions (y-axis) of all AOIs according to the RF2 models as a function of the molecular group of the participants (x-axis). Note that in **c**, contrary to model RF1, because RF2 was trained on a dataset with limited intra-tumoural heterogeneity, discrepancies between reference and predictions are more likely to be due to intra-tumour heterogeneity than in **a**. **d**, Weighted F1 scores of the three sets of predictions presented in the confusion matrices (**a**–**c**).

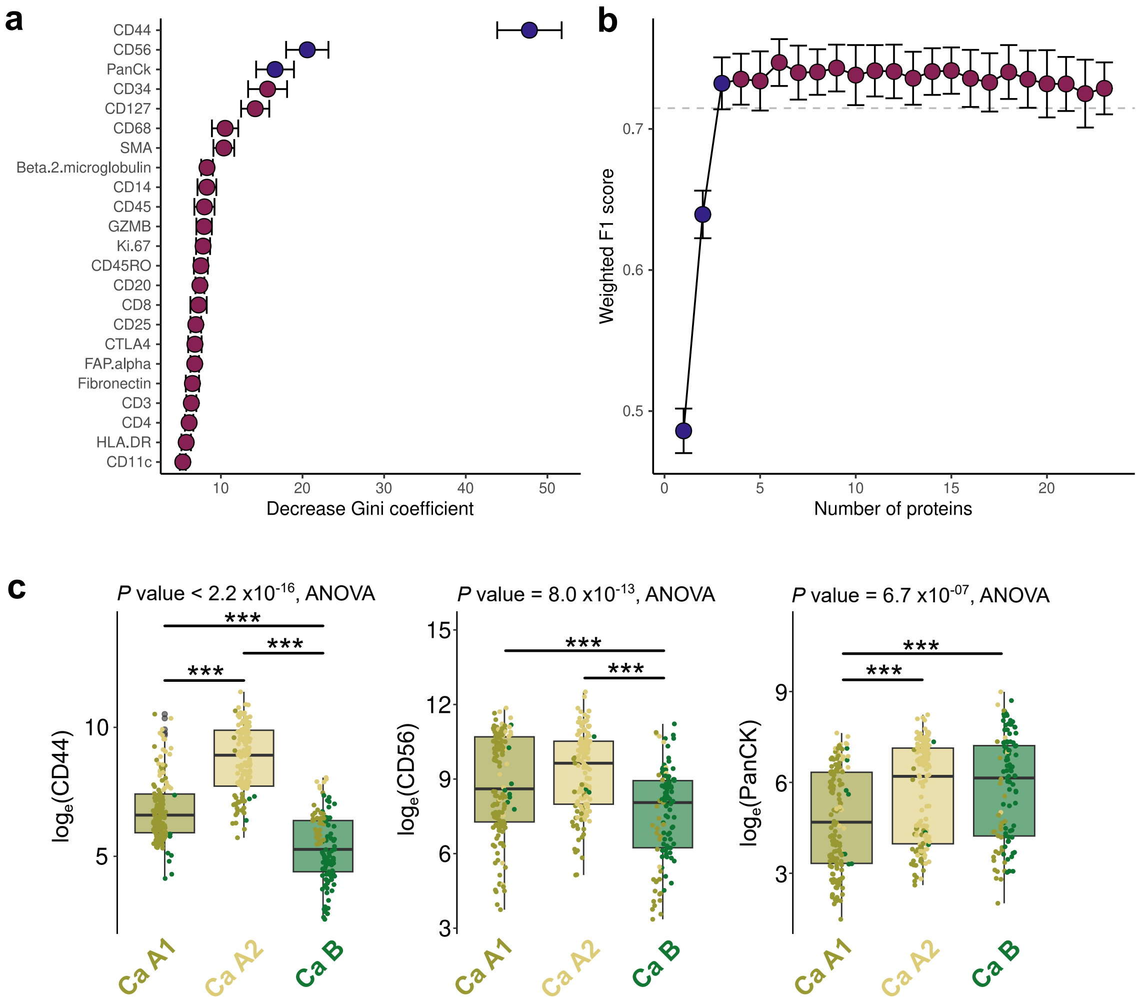

**Supplementary Figure S7. Identification of the most important proteins for predicting molecular group with digital spatial profiling.** **a**, Mean decrease in Gini coefficient per protein included in the RF2 model (y-axis). Error bars correspond to the mean plus or minus standard deviation. **b**, Mean weighted F1 scores of predictions from random forest models trained on the most explanatory proteins. The number of probes included in each model is given on the x-axis. Error bars correspond to the mean plus or minus standard deviation. In **a**, blue points correspond to the 3 most important proteins and in **b**, blue points correspond to RF models including these proteins. **c**, Distribution of measurements of the three most explanatory proteins of the digital spatial profiling experiments as a function of patient molecular group. The points correspond to the values of each area of interest (AOI) and their colour corresponds to their molecular group as predicted by random forest. Pairwise comparisons in mean loge protein expression performed with Mann-Whitney U tests. * 0.01 < *P* value < 0.05; ** 0.001 < *P* value ≤ 0.01; *** *P* value ≤ 0.001.

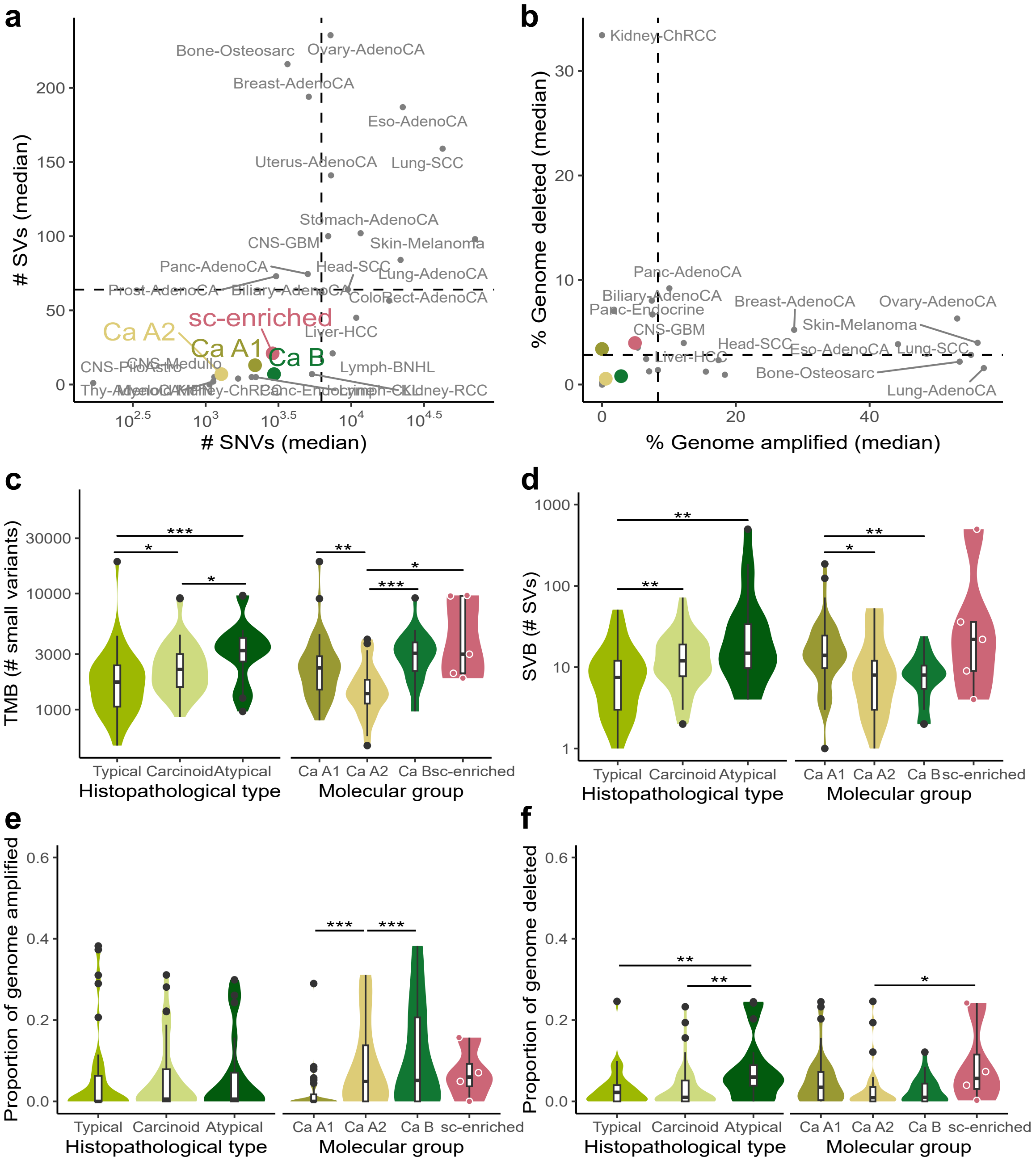

**Supplementary Figure S8. Mutational burden in lung NETs.** **a**, Median number of small variants (SNVs plus indels, x-axis) and structural variants (SVs, y-axis) in LNET cohort molecular groups ($n$ = 102) compared to 25 common cancers from the PCAWG consortium ($n$ = 2,778). **b**, Median percentage of the genome amplified (x-axis) and deleted (y-axis), in lung NET molecular groups compared to common cancers from the PCAWG consortium. Samples and colours of molecular groups are as per (**a**); dashed lines represent the median values across PCAWG cancer types. **c**, Violin plot of the distribution of the tumour mutational burden (TMB, number of small variants) in the lung NET cohort (y-axis) as a function of the histopathological classification or molecular group classification (x-axis). **d**, As per (**c**) for the structural variant burden (SVB, number of structural variants). **e**, As per (**c**) for the amplified copy number burden (proportion of the genome amplified). **f**, As per (**c**) for the deleted copy number burden (proportion of the genome deleted). In (**c**-**f**), significance levels correspond to t-tests. * 0.01 < $P$ value < 0.05; ** 0.001 < $P$ value ≤ 0.01; *** $P$ value ≤ 0.001.

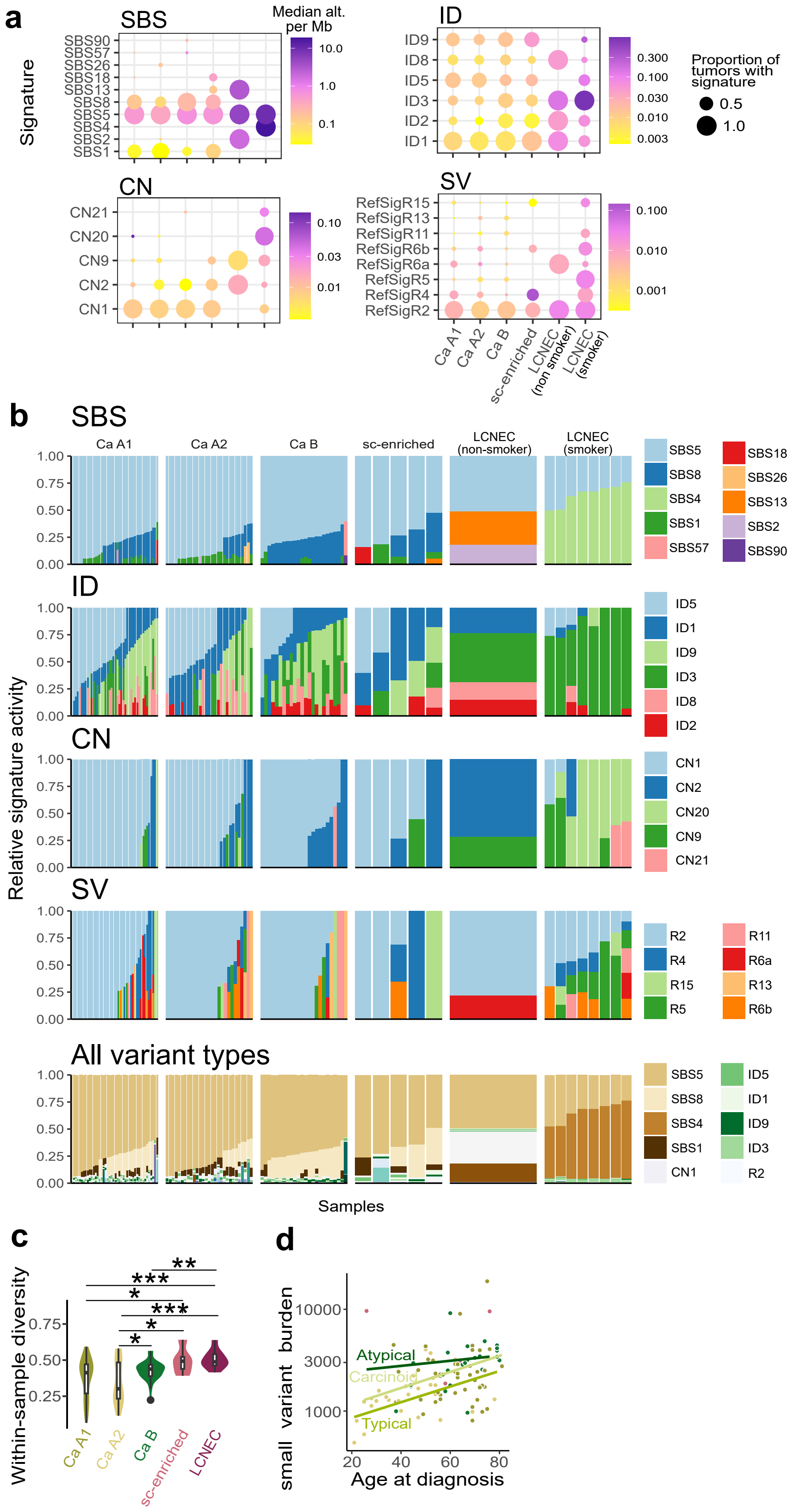

**Supplementary Figure S9. The repertoire of mutational signatures in lung NETs and LCNEC. a**, Dot plot of mutational signature burdens for different types of variants. Dot size represents the proportion of tumours with a given signature, and dot colours represent the median number of alterations per megabase among tumours with the signature present. SBS, single base substitution; ID, Indel; CN, copy number; SV, structural variant. **b**, Barplots of the relative proportion of signatures (y-axis) across samples (x-axis) for different types of variants. **c**, Signature variability analysis showing the within-sample diversity measure (y-axis) which corresponds to the mean Gini-Simpson index of each molecular group (x-axis). Analysis takes into account mutational signature proportions from all variant types. Asterisks represent significance codes for two-sided t-tests. **d**, Relationship between age at diagnosis (x-axis) and number of small variants (y-axis), in log10 scale. Lines represent linear models of age as a function of the number of variants by molecular group. In all plots and analyses, signatures with a relative activity below 5% were removed, and plots represent the lung NEN cohort with WGS data available ($n$ = 111, $n$ = 102 samples of the lung NET cohort plus $n$ = 9 LCNEC). * 0.01 < $P$ value < 0.05; ** 0.001 < $P$ value $\leq$ 0.01; *** $P$ value $\leq$ 0.001.

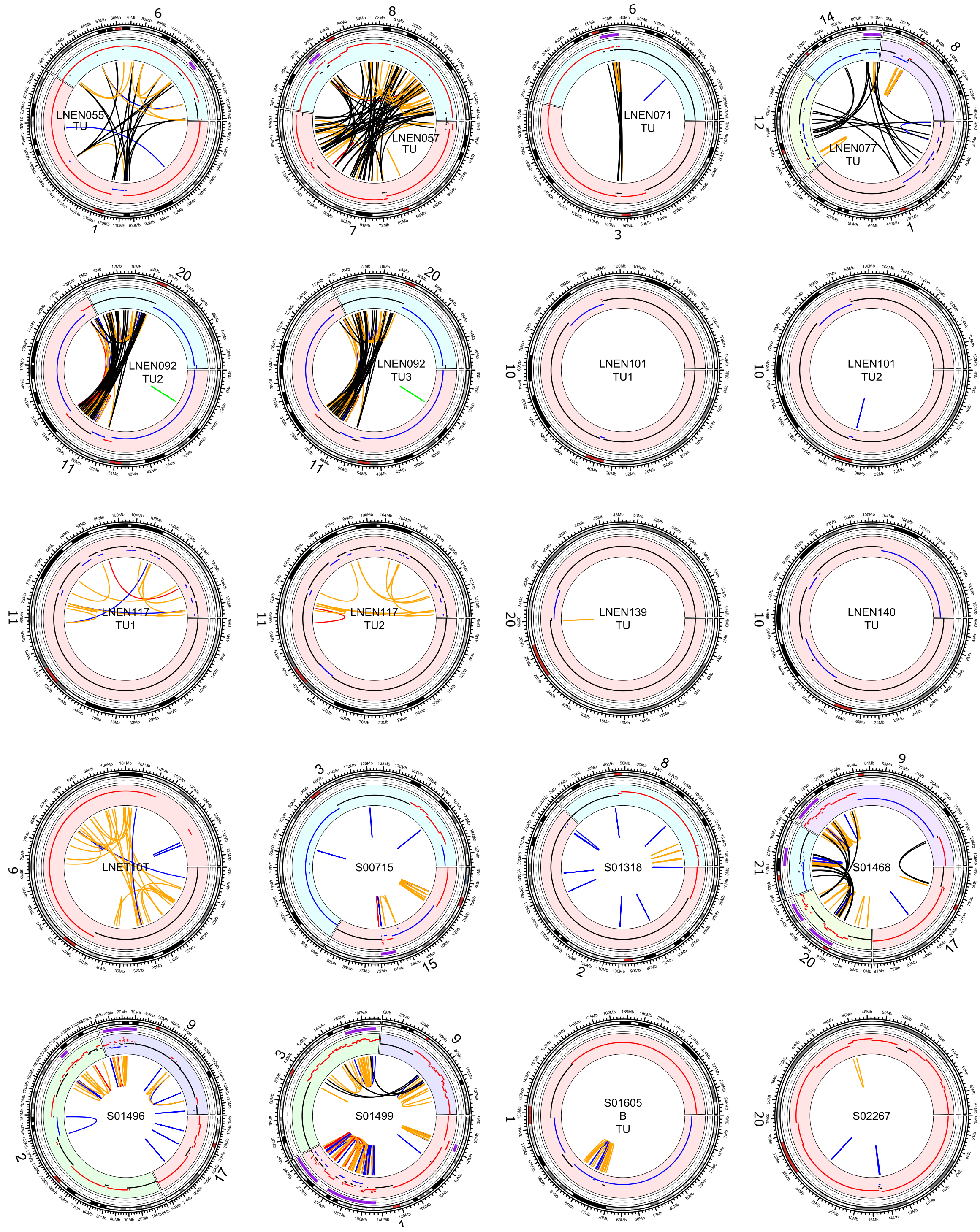

**Supplementary Figure S10. Shattered regions.** Each plot corresponds to a circos plot of a tumour sample (sample id centered) where a shattered region was detected with R package svpluscnv. Genomic coordinates are represented circularly. Inward to outward: structural variants (colored arcs, black for translocations, orange for inversions, blue for deletions, red for duplications), CNVs (red segments for amplifications, blue for deletions, black for normal copy number), detected shattered regions (purple bars), and ideogram with genomic coordinates (colors represent cytogenetic bands).

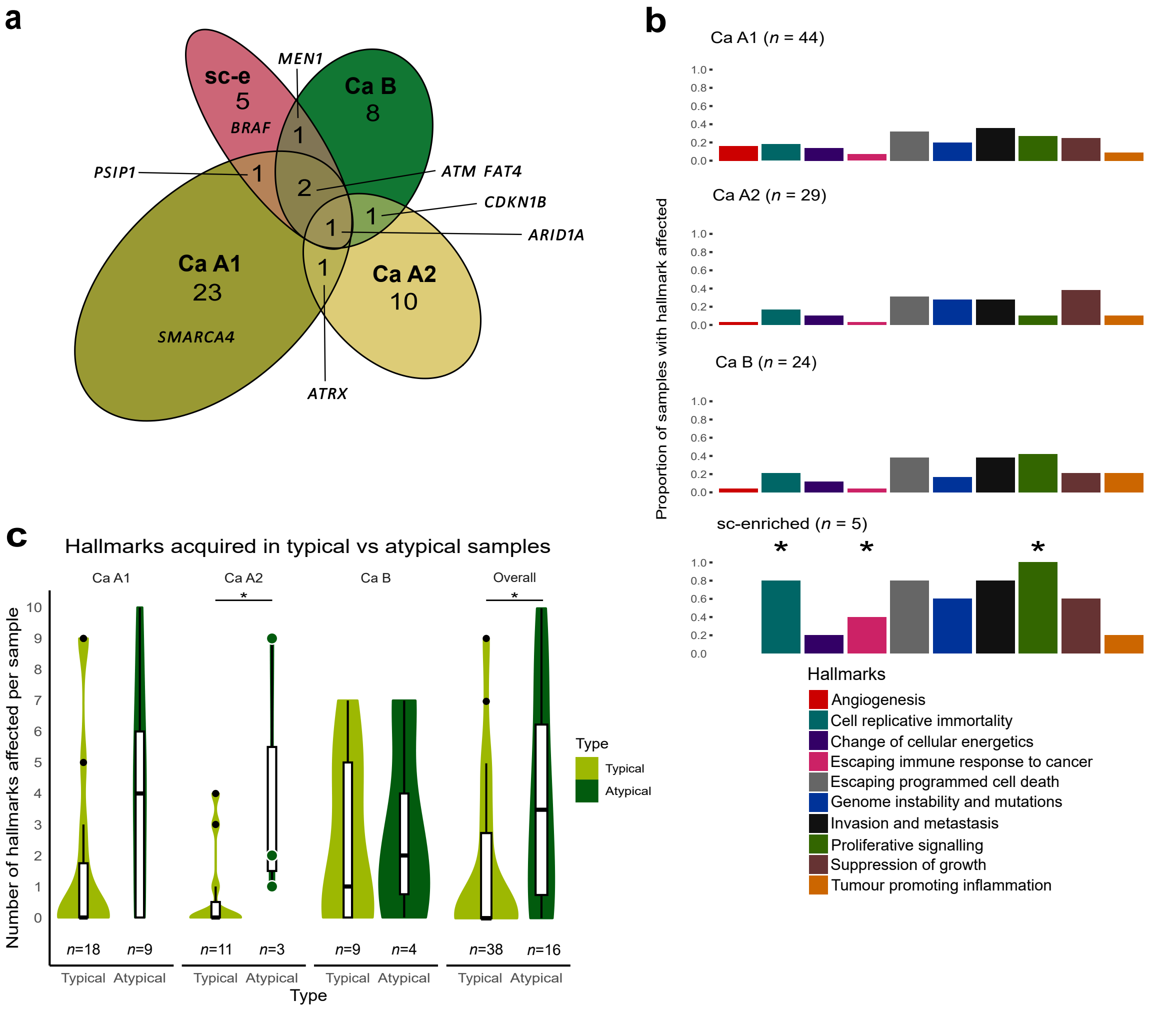

**Supplementary Figure S11. Genomic hallmarks of cancer affected by damaging small variants within lung NET molecular groups. a**, Euler diagram of genes involved in hallmark acquisition. Each gene can only belong to one molecular group on the diagram. The genes shown on the diagram as examples are those attributed to at least two different participants. **b**, Average hallmark profile for each lung NET molecular group. Each bar represents the proportion of samples where that hallmark is affected by damaging small variants. Asterisks indicate significance level (*P* value) obtained from logistic regression models, or in the case of hallmarks Angiogenesis and Proliferative signalling, Fisher's exact tests, performed between archetype Ca A1 and each other archetype individually. **c**, Distribution of the number of hallmarks acquired through damaging small variants per sample in typical versus atypical tumours by molecular group, and overall. The overall distribution does not include sc-enriched samples. Comparisons made with Mann-Whitney U tests. * 0.01 < *P* value < 0.05; ** 0.001 < *P* value ≤ 0.01; *** *P* value ≤ 0.001.

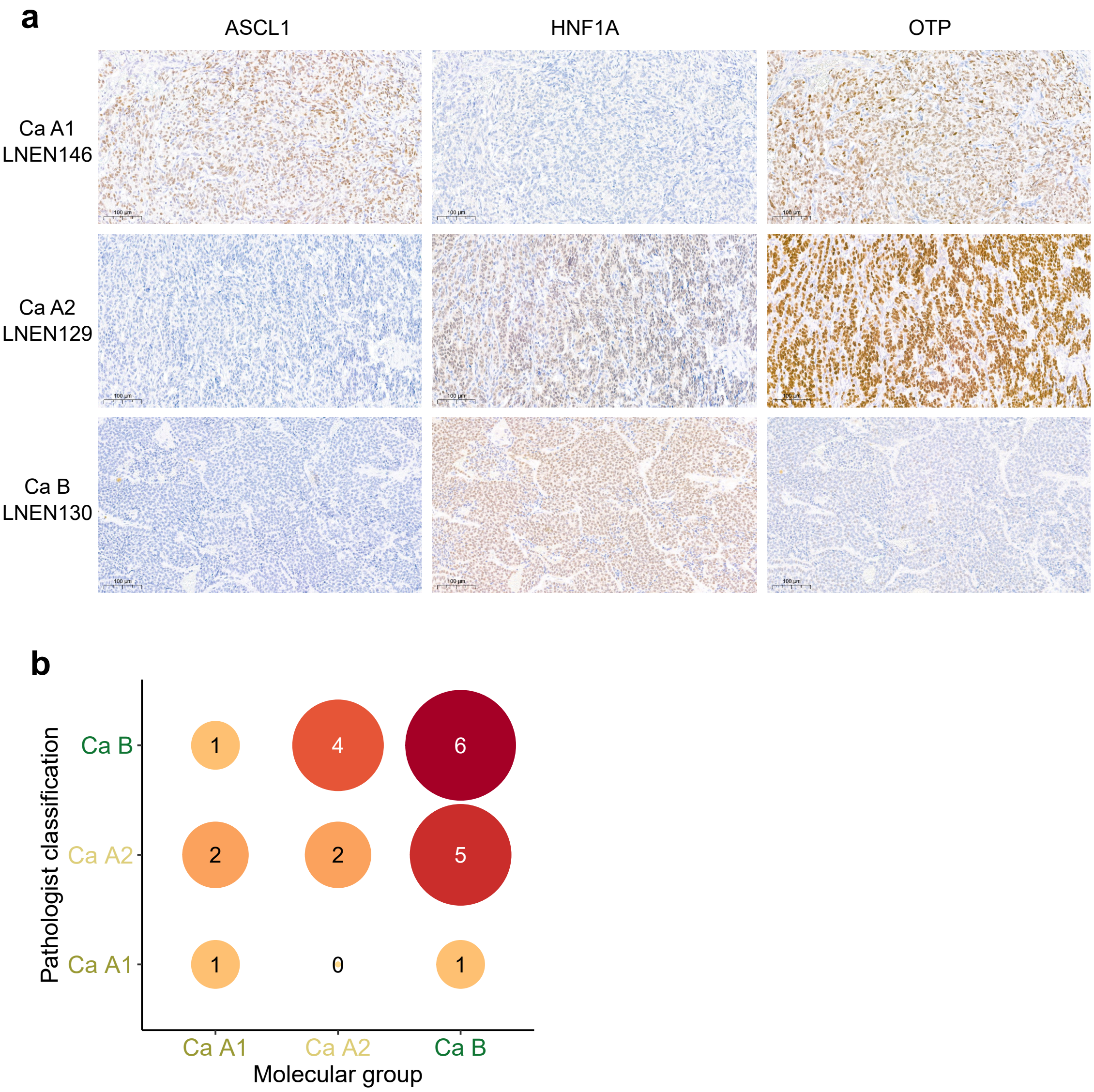

**Supplementary Figure S12. IHC and morphological classifications. a**, Representative images of one patient slide per molecular group (rows) stained for ASCL1, HNF1A and OTP (columns), to identify molecular groups Ca A1, Ca A2, and Ca B. **b**, Confusion matrix of the performance of the morphological classification for molecular groups, *n* = 22.

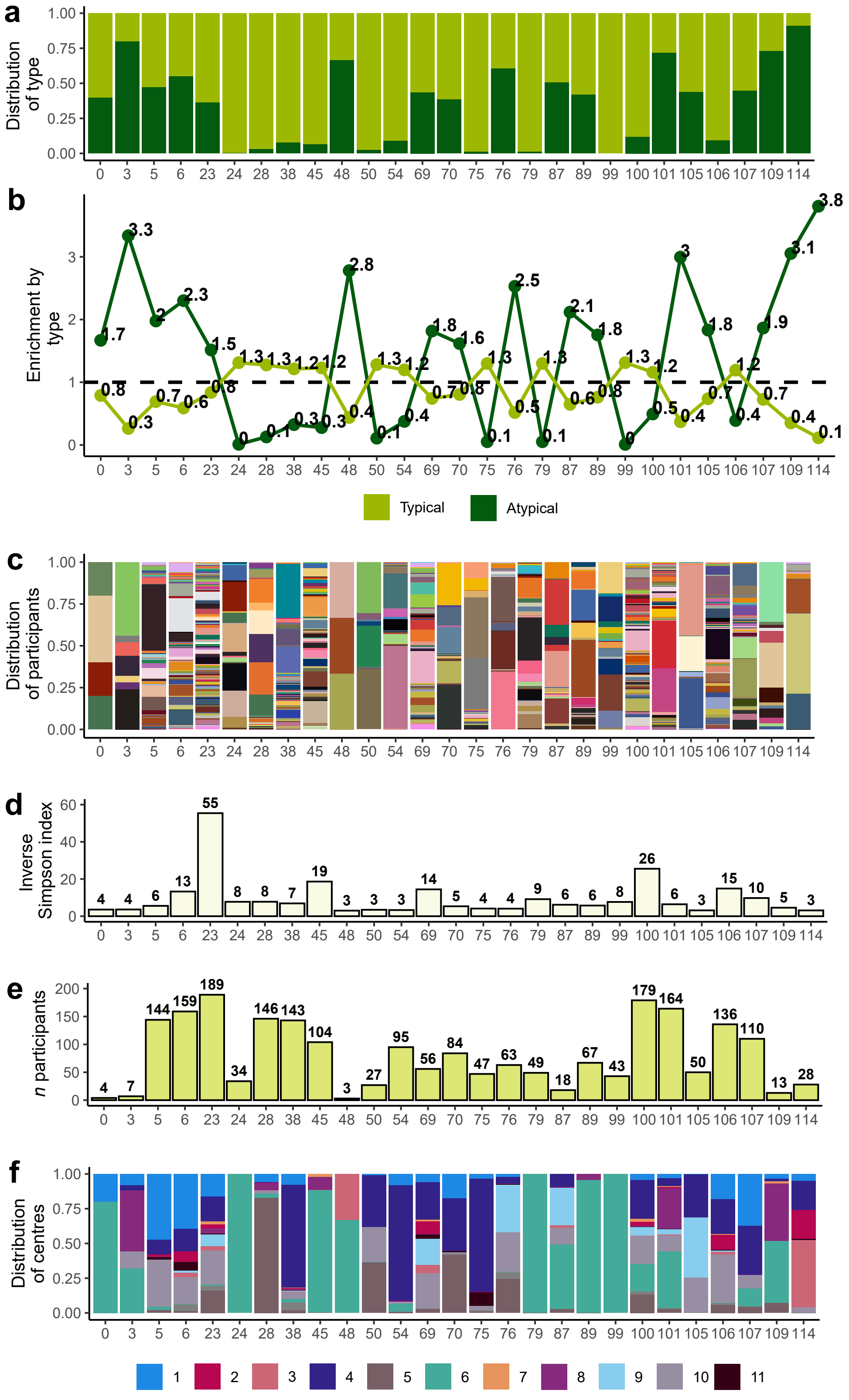

**Supplementary Figure S13. Overview of Leiden partitions (self-supervised branch) selected to predict histological types. a**, Proportion of tiles (y-axis) from whole-slide images (WSIs) from participants with typical and atypical tumours per selected Leiden partition (x-axis). **b**, Enrichment score (y-axis) calculated separately for typical and atypical tumours per partition (x-axis). Dashed horizontal line indicates a score of 1 implying that the proportion of tiles of histological type T within the partition is equal to the proportion of tiles of histological type T within the dataset. **c**, Proportion of tiles (y-axis) per WSI, represented by different colours, per partition (x-axis). **d**, Inverse Simpson index (y-axis) per partition (x-axis). Value indicates the number of WSIs that tiles within the partition are primarily extracted from. **e**, Total number of WSIs (y-axis) contributing tiles to each partition (x-axis). **f**, Proportion of tiles from each contributing centre, represented by colours (y-axis), per partition (x-axis).

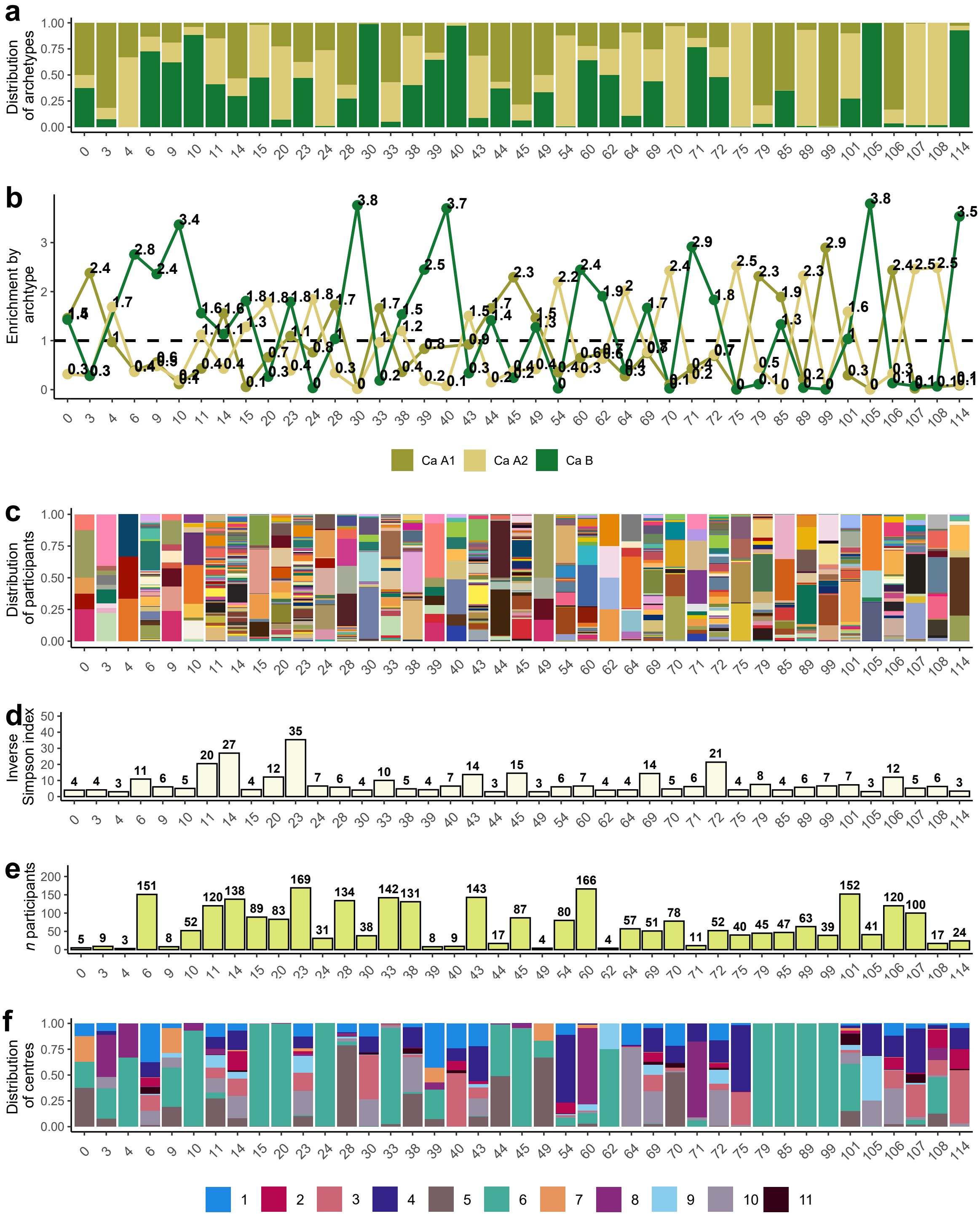

**Supplementary Figure S14. Overview of Leiden partitions (self-supervised branch) selected to predict molecular groups. a**, Proportion of tiles (y-axis) from whole-slide images (WSIs) from participants with Ca A1, Ca A2, and Ca B tumours per selected Leiden partition (x-axis). **b**, Enrichment score (y-axis) calculated separately for three molecular groups per partition (x-axis). Dashed horizontal line indicates a score of 1 implying that the proportion of tiles of molecular group type M within the partition is equal to the proportion of tiles of molecular group M within the dataset. **c**, Proportion of tiles (y-axis) per WSI, represented by different colours, per partition (x-axis). **d**, Inverse Simpson index (y-axis) per partition (x-axis). Value indicates the number of WSIs that tiles within the partition are primarily extracted from. **e**, Total number of WSIs (y-axis) contributing tiles to each partition (x-axis). **f**, Proportion of tiles from each contributing centre, represented by colours (y-axis), per partition (x-axis).

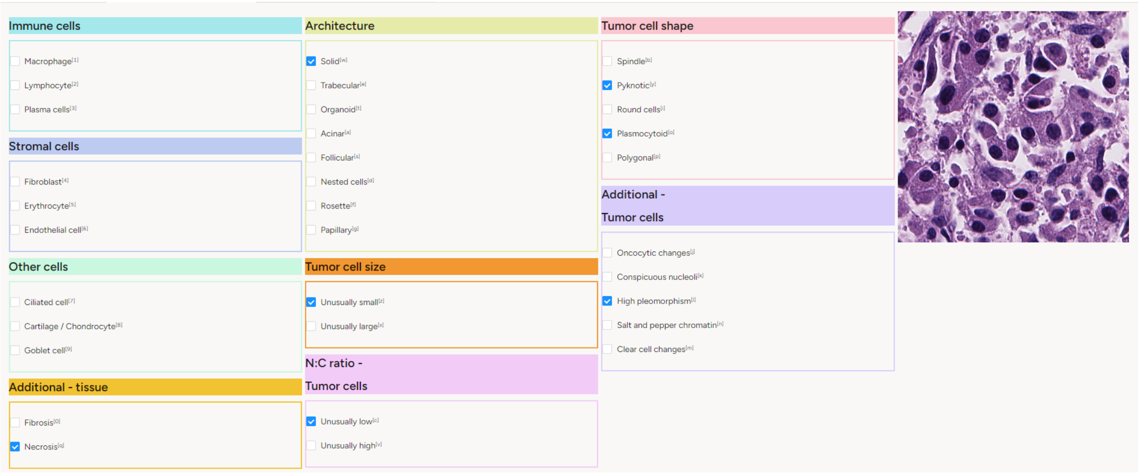

**Supplementary Figure S15. Screen shot of the Label Studio online platform for tile-by-tile annotations.** While visualising each individual tile, to the right, pathologists indicated by checking boxes, which features were present within the tile for nine evaluation categories.

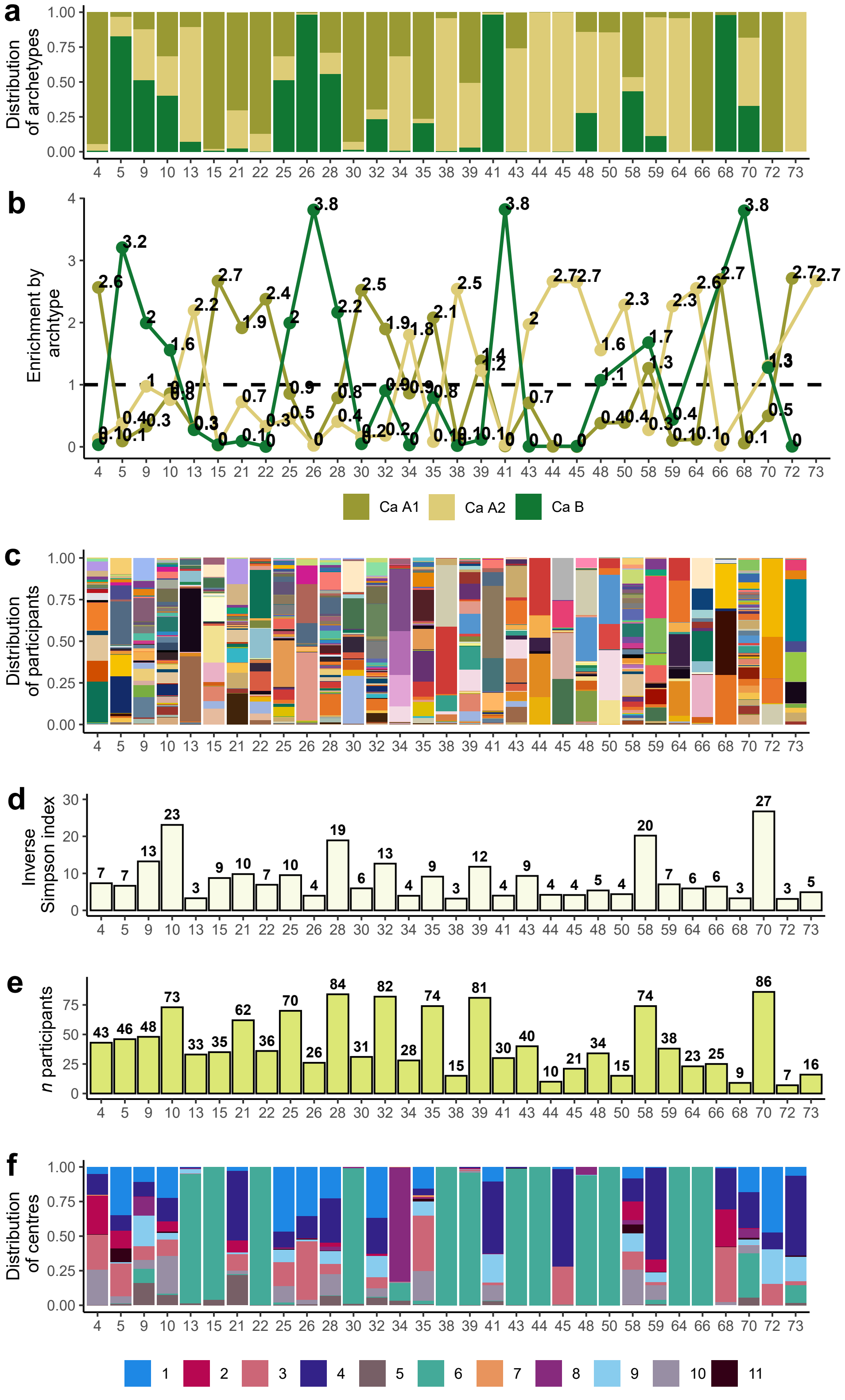

**Supplementary Figure S16. Overview of Leiden partitions (supervised branch) selected for pathological review. a**, Proportion of tiles (y-axis) from whole-slide images (WSIs) from participants with Ca A1, Ca A2, and Ca B tumours per selected Leiden partition (x-axis). **b**, Enrichment score (y-axis) calculated separately for three molecular groups per partition (x-axis). Dashed horizontal line indicates a score of 1 implying that the proportion of tiles of molecular group type M within the partition is equal to the proportion of tiles of molecular group M within the dataset. **c**, Proportion of tiles (y-axis) per WSI, represented by different colours, per partition (x-axis). **d**, Inverse Simpson index (y-axis) per partition (x-axis). Value indicates the number of WSIs that tiles within the partition are primarily extracted from. **e**, Total number of WSIs (y-axis) contributing tiles to each partition (x-axis). **f**, Proportion of tiles from each contributing centre, represented by colours (y-axis), per partition (x-axis).

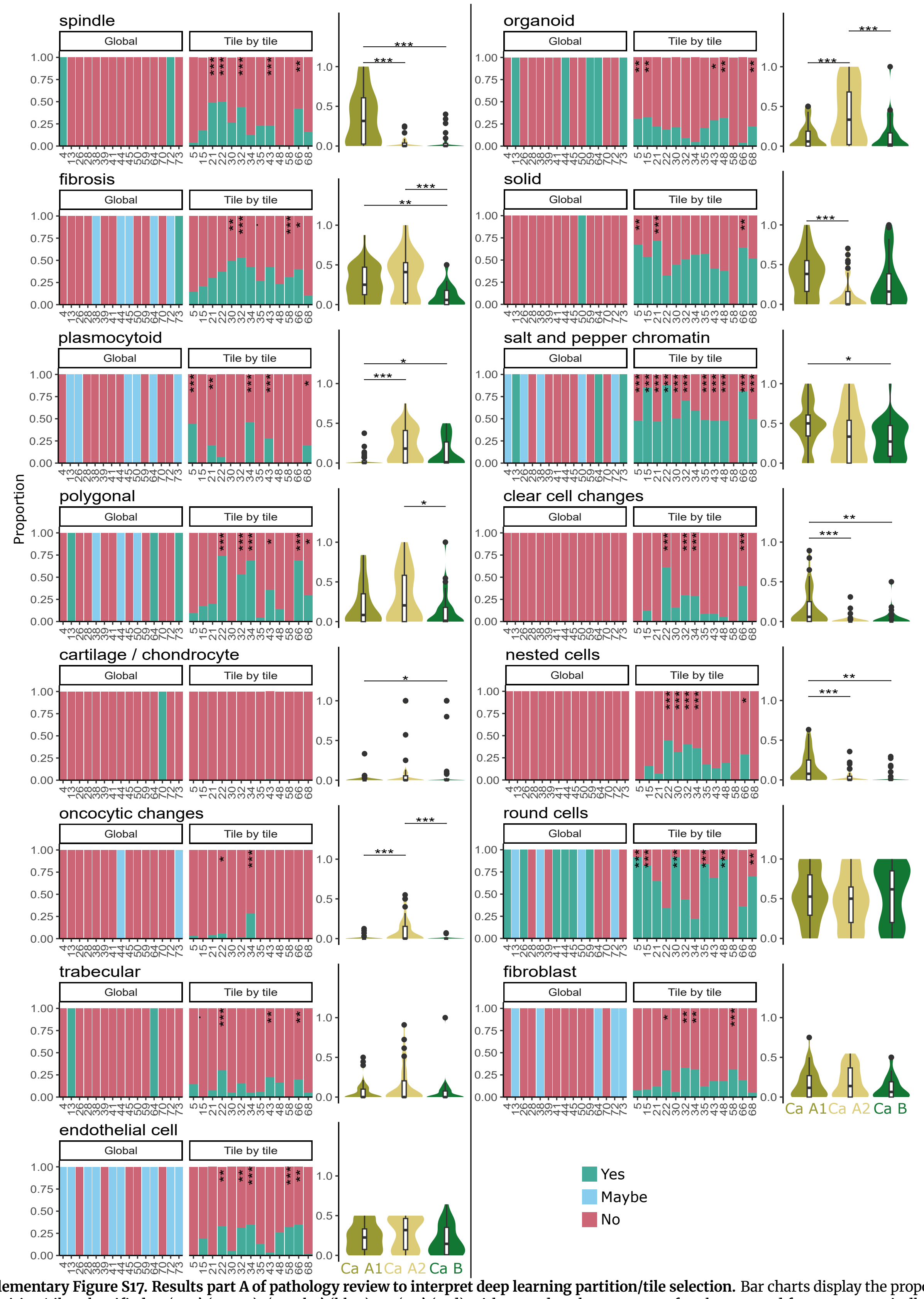

**Supplementary Figure S17. Results part A of pathology review to interpret deep learning partition/tile selection.** Bar charts display the proportion of partition/tiles classified as ‘yes’ (green), ‘maybe’ (blue), or ‘no’ (red) with regard to the presence of each proposed feature, e.g. spindle cells, (y-axis) per partition (x-axis). Partitions evaluated globally are described in column one, those annotated on a tile-by-tile basis are described in column two. Asterisks in column two indicate significance level (*q* value) obtained from Fisher’s exact tests. * 0.01 < *q* value < 0.05; ** 0.001 < *q* value ≤ 0.01; *** *q* value ≤ 0.001. Violin and boxplots in column three indicate the proportion of tiles per molecular group classified as ‘yes’ and ‘maybe’ with regard to the presence of the feature. For partitions annotated tile-by-tile, the proportions of ‘yes’ and ‘maybe’ were summed, with the ‘maybe’ level assigned half the weight of the ‘yes’ level. For globally annotated partitions, ‘yes’ corresponded to 100% of the tiles displaying the feature, ‘maybe’ to 50% of the tiles, and ‘no’ to 0% of the tiles displaying the feature. Asterisks in column three indicate the significance level (*q* value) obtained from permutation tests performed between groups. * 0.01 < *q* value < 0.05; ** 0.001 < *q* value ≤ 0.01; *** *q* value ≤ 0.001.

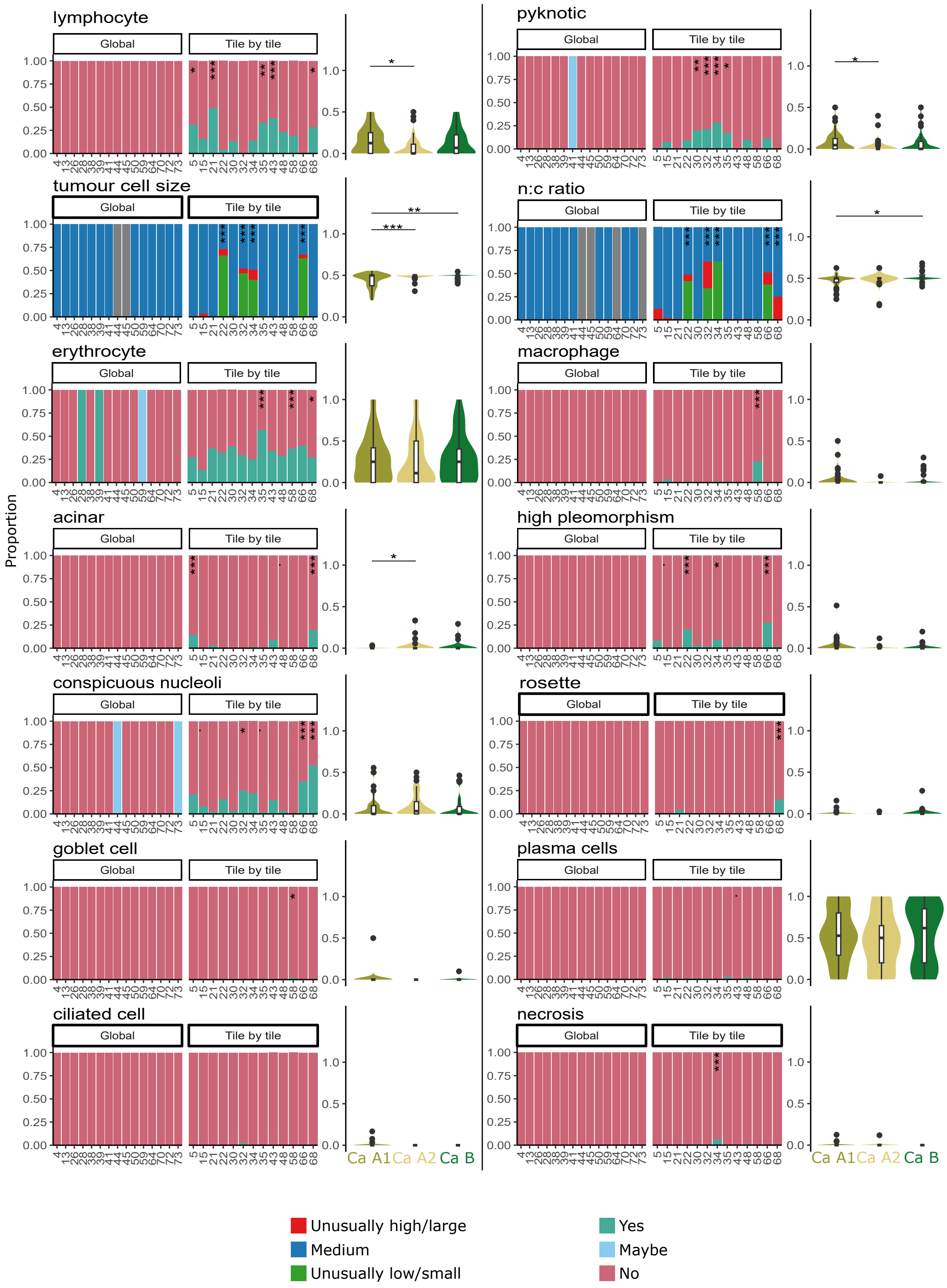

**Supplementary Figure S18. Results part B of pathology review to interpret deep learning partition/tile selection.** As per **Supplementary Figure S17**, for different features. N:C ratio, nucleus to cytoplasm ratio. Classification labels for N:C ratio and tumour cell size differ to all other characteristics evaluated. Labels were only assigned to partitions/tiles if both pathologists agreed, otherwise the partition/partition was marked as NA (grey).

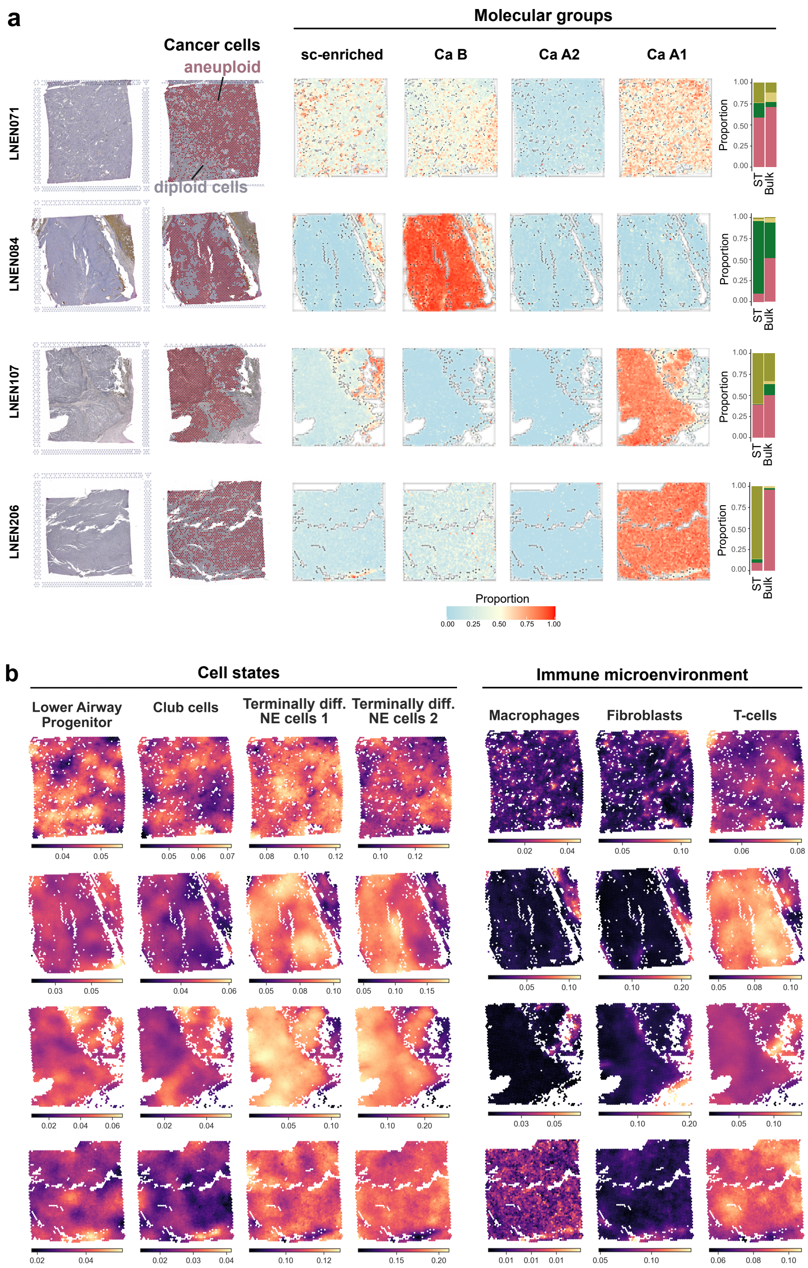

**Supplementary Figure S19. Spatial transcriptomics cell types analysis of four supra-carcinoid samples. a**, Haematoxylin/eosin-stained slide images of four supra-carcinoid tumour samples used in spatial transcriptomic analyses (column 1). Visualisation of cancer cells, defined as aneuploid cells (red), versus non-cancer cells, defined as diploid cells (grey), within each sample slide (column 2). Location of cells with a profile matching that of one of the four lung NET molecular groups, computed using deconvolution of the spots with the average bulk gene expression profile of each group as reference (columns 3-6). Bar charts comparing the proportion of molecular group within the spatial transcriptomic slide (ST; averaged from columns 3-6), and multi-omic sequenced bulk tumour tissue from the same patient, for each of the four samples. **b**, Location of cells with one of four cell profiles, corresponding to the potential cells of origin for the tumours, estimated using spot expression deconvolution (columns 1-4). Location of the main cell types from the tumour immune microenvironment, estimated using spot expression deconvolution (columns 5-7).

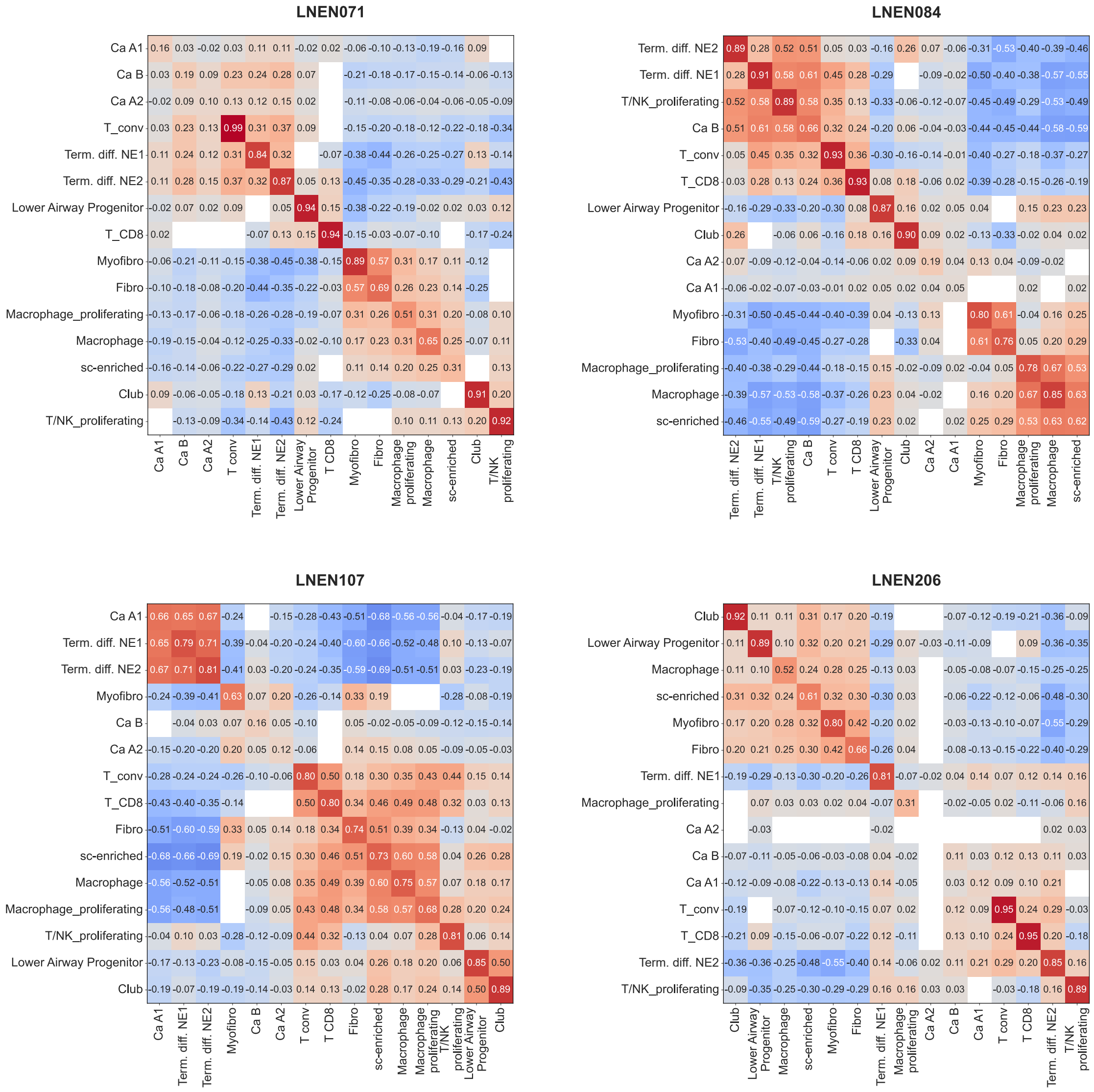

**Supplementary Figure S20. Spatial transcriptomics cell type correlations of four supra-carcinoid samples.** Correlation matrices displaying bivariate spatial cross-correlation (bivariate Moran's R) coefficients for cell type proportions for each supra-carcinoid sample analysed. White areas indicate non-significant results.

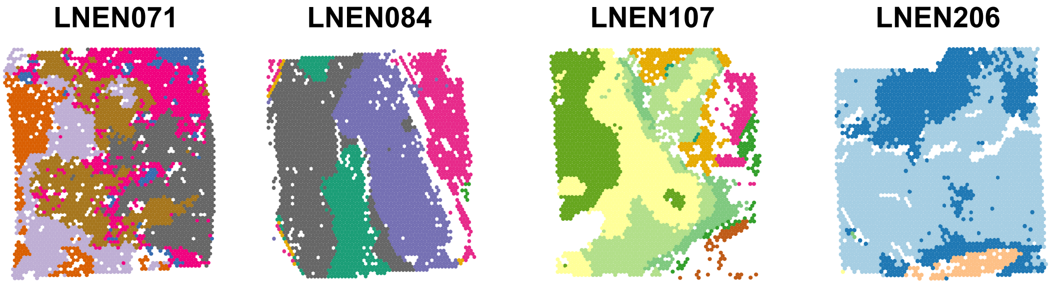

**Supplementary Figure S21. Spatial transcriptomics spatial domains of four supra-carcinoid samples.** Spatial domains (represented by colours) estimated using method IRIS across the four slides.

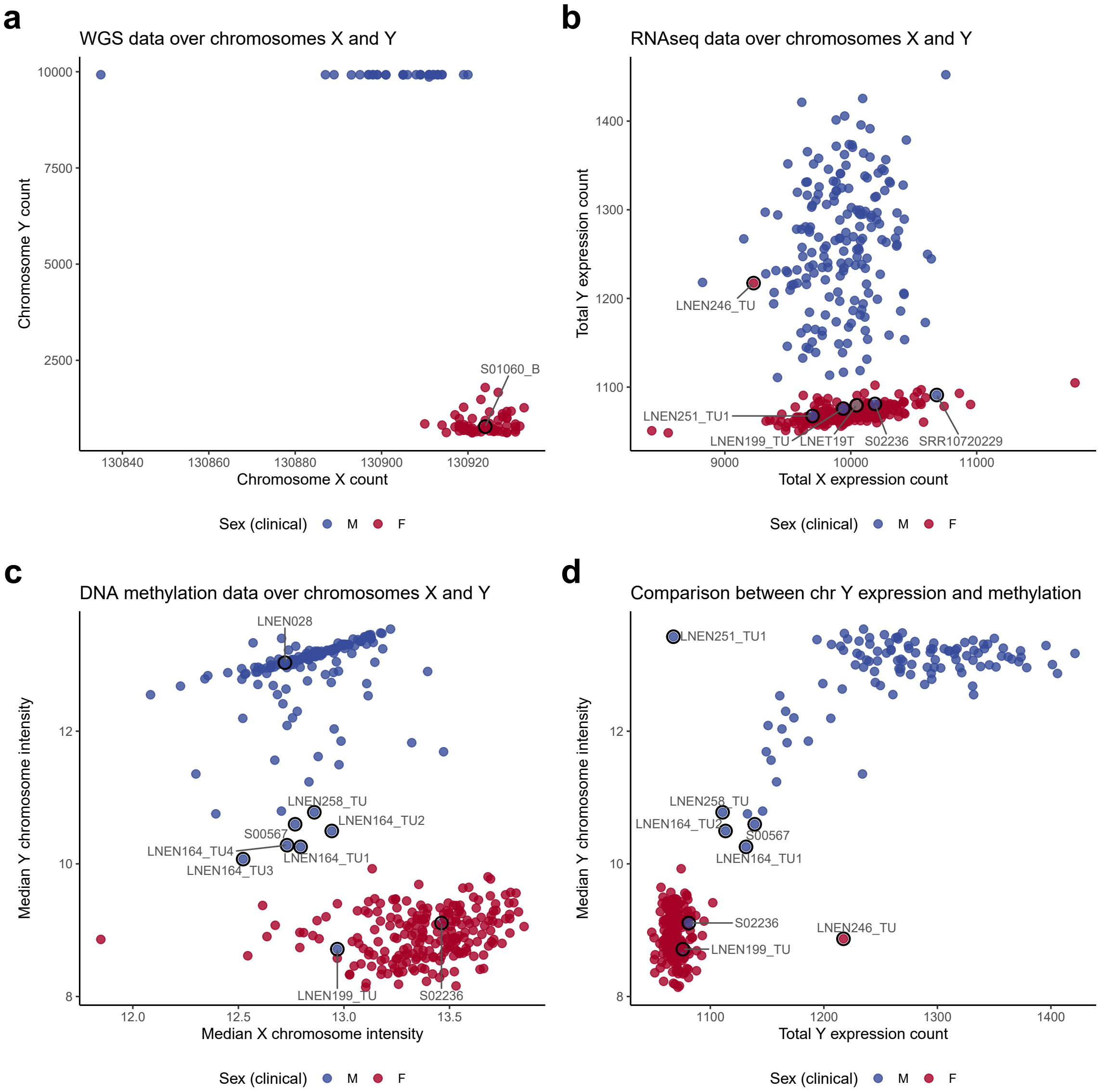

**Supplementary Figure S22. Sex chromosome omics. a**, Total read count on the X (x-axis) and Y (x-axis) chromosomes per sample (*n* = 98). **b**, Total expression level of the X (x-axis) and Y (y-axis) chromosomes (sum of variance-stabilised read counts) per sample (*n* = 408). **c**, Median DNA methylation array total intensity on the X (x-axis) and Y (y-axis) chromosomes per sample (*n* = 356). **d**, Comparison between total Y chromosome expression (x-axis) and median DNA methylation array total Y chromosome intensity (y-axis) for each sample profiled by both RNA sequencing and DNA methylation (*n* = 282). Point colours correspond to clinically reported sex, M, male; F, female; grey is unreported. Samples circled and labelled in black were discordant between clinically reported sex and predicted sex by at least one omic type.

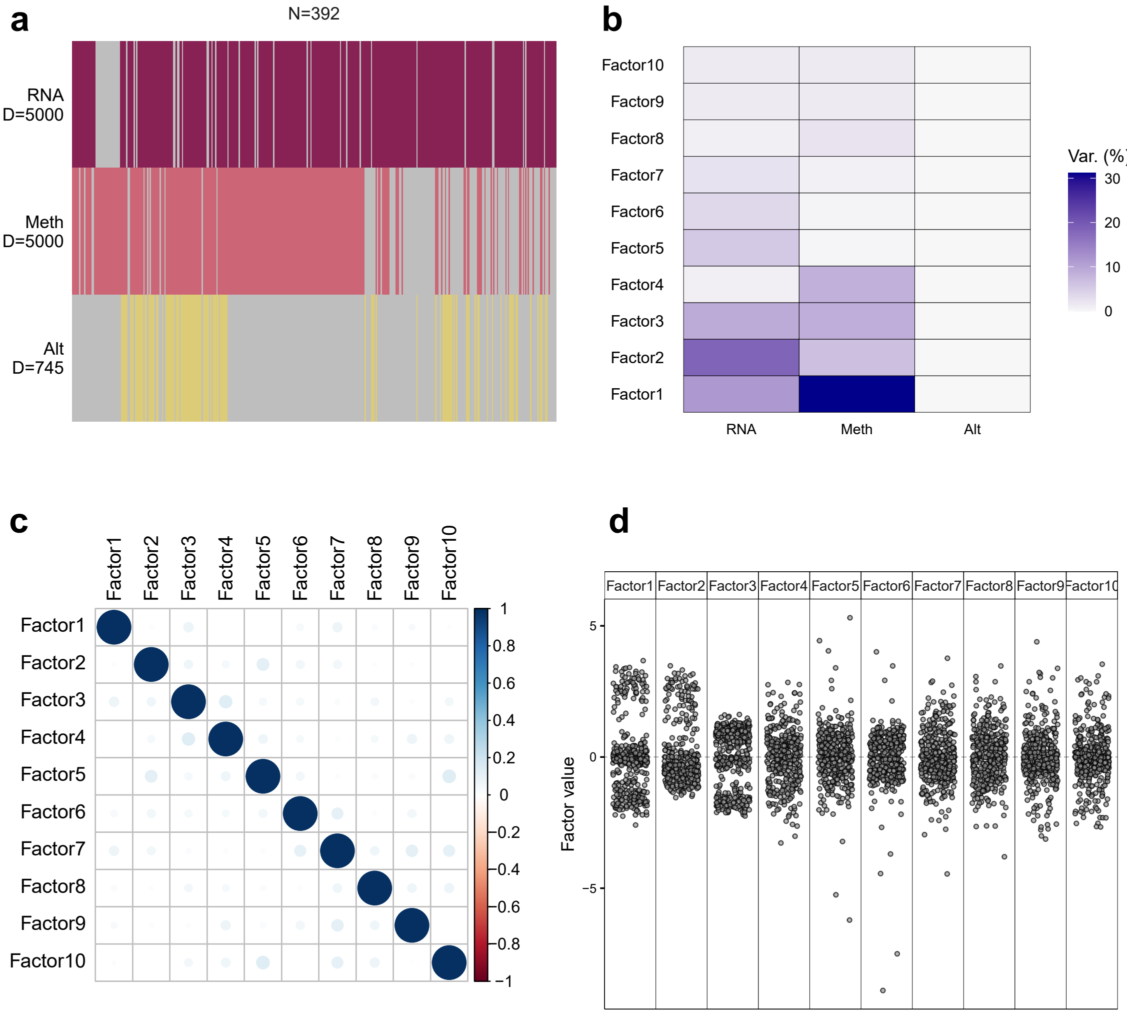

**Supplementary Figure S23. Overview of inputs and outputs of lung NEN cohort MOFA. a**, Input data matrices. D is the number of omic features incorporated per matrix [expression levels for 5000 genes (RNA), DNA methylation levels at 5000 CpG sites (Meth), small and/or structural variant status at 745 genes (Alt)]. Grey indicates missing data. **b**, Variance explained by each latent factor for each data type. **c**, Correlation between sample coordinate along latent factors. **d**, Distribution of samples along each latent factor.

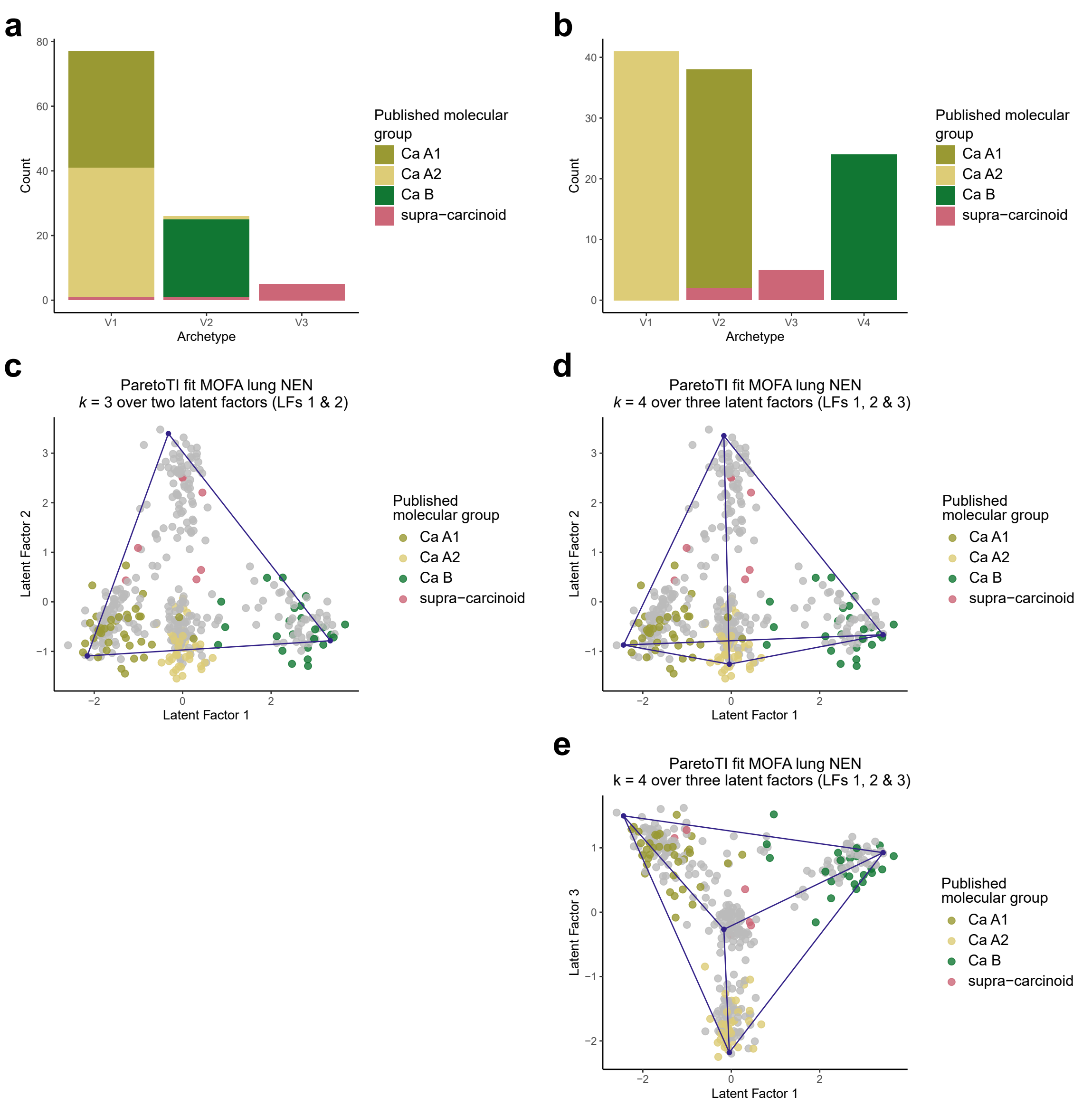

**Supplementary Figure S24. Correspondence between ParetoTI archetypes identified in the lung NEN cohort and previously published molecular groups. a-b**, Count of samples from Alcala *et al.* 2019 ($n$ = 107) and Dayton *et al.* 2023 ($n$ = 1) coloured by published molecular group within archetypes found in the current study when considering $k$ = 3 (**a**) and $k$ = 4 (**b**) archetypes. **c-e**, Scatterplots showing the coordinates of samples from Alcala *et al.* 2019 and Dayton *et al.* 2023 (coloured by previously published molecular group), remaining lung NEN cohort samples ($n$ = 284, grey points), and archetype proportions (blue points) along latent factors for (**c**) $k$ = 3 archetypes derived from latent factors (LFs) 1 and 2, and (**d-e**) $k$ = 4 archetypes derived from LFs 1, 2 and 3.

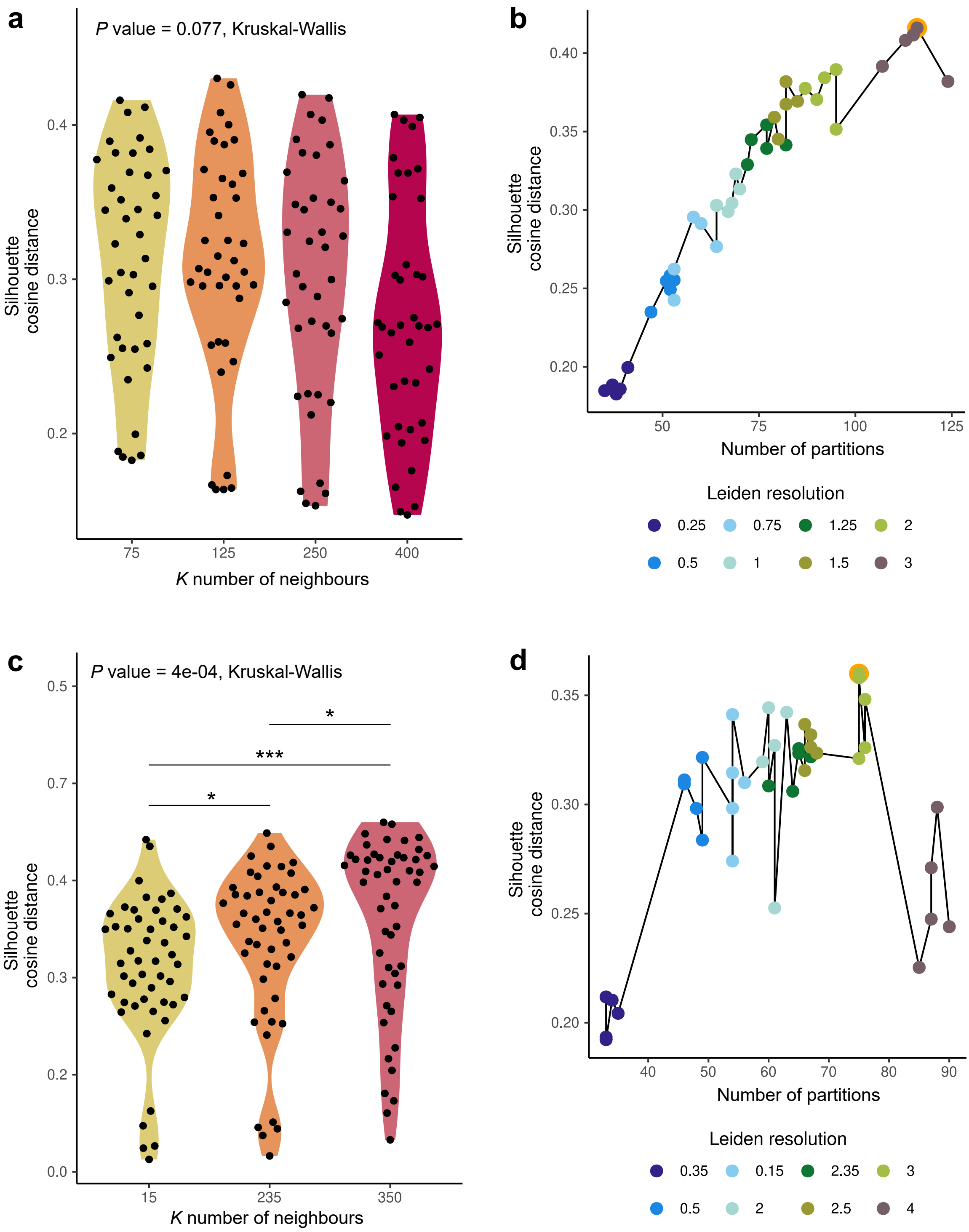

**Supplementary Figure S25. Selection of parameters for Leiden clustering. a-b**, Self-supervised branch. Selection of parameters *K* and $\gamma$ was performed using silhouette scores, calculated according to cosine distances. **a**, Distribution of silhouette coefficients (y-axis) considering different values of *K* number of neighbours between nodes (x-axis). **b**, Silhouette coefficients (y-axis) as a function of the number of the number of partitions (x-axis) for different values of Leiden resolution parameter $\gamma$ (coloured points). The Leiden clustering that maximises the silhouette coefficient for selected value of *K* (*K* = 75) is circled in orange and corresponds to a partition obtained with a resolution of $\gamma$ = 3. **c-d**, Supervised branch. Clustering performed following calculation of attention scores by RoFormer-MIL. Selection of parameters *K* and $\gamma$ was performed using silhouette scores, calculated according to cosine distances. **c**, Distribution of silhouette coefficients (y-axis) considering different values of *K* number of neighbours between nodes (x-axis). Asterisks indicate significance level (*P* value) obtained from Mann-Whitney U tests performed between values of *K*. * 0.01 < *P* value < 0.05; ** 0.001 < *P* value $\leq$ 0.01; *** *P* value $\leq$ 0.001. **d**, Silhouette coefficients (y-axis) as a function of the number of the number of partitions (x-axis) for different values of Leiden resolution parameter $\gamma$ (coloured points). The Leiden clustering that maximises the silhouette coefficient for chosen value of *K* (*K* = 250) is circled in orange and corresponds to a partition obtained with a resolution of $\gamma$ = 2.
