## Supplementary material for "A clinically relevant morpho-molecular classification of lung neuroendocrine tumours": Methods_11072025_medrxiv.pdf

### Methods for article "A clinically relevant morpho-molecular classification of lung neuroendocrine tumours"

### 1. Study cohort

#### 1.1 The lungNENomics project cohort

The majority of samples used in this study are from the lungNENomics project cohort, a multi-national mixed retrospective and prospective cohort of over 400 lung neuroendocrine tumour patients, established by L.F-C and M.F of the Computational Cancer Genomics Team.

Fresh-frozen and formalin-fixed, paraffin-embedded (FFPE) tumour tissues, fresh-frozen adjacent normal lung tissue, and whole blood were collected at diagnosis during surgical resection from 12 contributing centres. Patients provided informed consent for tissue collection and its use in histopathological and molecular analyses (including somatic and germline whole-genome sequencing), as well as for the collection of de-identified clinical data. This study was approved by the International Agency for Research on Cancer Ethics Committee (project number 19-07).

In total, 201 patients from the lungNENomics cohort underwent molecular analysis for this study. WGS was performed for 72 patients, RNA sequencing for 179, and DNA methylation array profiling for 191. All three types of omics data were generated for 60 patients, while an additional 109 were covered by both RNA sequencing and DNA methylation array profiling. For 41 patients, multi-region samples were available (intra-tumoural heterogeneity, ITH, samples). These patients had between two and seven tissue samples from different tumour regions analysed. Four patients underwent spatial transcriptomics (see **section 4**), and 64 underwent spatial proteomics (see **section 10**).

##### *1.1.1 Central pathology review*

Where FFPE material was available, samples underwent blinded central pathology review by six pathologists (L.B., S.L., A.M-L., M.G.P., G.P., and J-M.V.). A detailed description of sample review and the subsequent outcome can be found in Mathian *et al.* 2023[1]. Briefly, i) pathologists were instructed to follow the guidelines in the WHO Classification of Thoracic Tumours 5th edition[2] for the classification as typical or atypical pulmonary carcinoid: mitotic count and presence or absence of necrosis, ii) each pathologist assigned a diagnosis of typical or atypical per patient, and iii) a final diagnosis was made based on the majority vote.

Central pathology review was performed on 187 of the 201 patients, and 10 additional patients from Fernandez-Cuesta *et al.* 2014[3] (see **section 1.3**). This resulted in a different diagnosis from that reported by the contributing centre in 55 cases. As the FFPE slides reviewed by the central pathology team were not necessarily the same slides used for initial diagnosis, it was decided that the review classification could only be used to upgrade (from typical to atypical) but not downgrade tumours. A final 'type' label was therefore assigned to each patient based on their initial and central pathology review diagnosis as follows: initial typical + review typical = typical; any initial diagnosis + review atypical = atypical; any initial diagnosis + review undetermined/insufficient tissue = carcinoid; initial atypical + review typical = carcinoid; any initial diagnosis + review 'AC NETG3 LCNEC' = NET G3. Samples that were not reviewed by central pathology ($n$ = 14) were labelled 'carcinoid'.

##### *1.1.2 DNA and RNA extraction*

Unless otherwise stated, whole-genome sequencing, RNA sequencing, and DNA methylation array data were generated from fresh-frozen tissue. Frozen tissue samples were evaluated by the study pathologist (S.L), those with at least 70% tumour content were selected for downstream DNA and RNA extraction.

DNA extraction was performed using the Gentra Puregene Tissue Kit (158667, Qiagen, samples extracted between 2018 and 2020), or the DNAdvance Tissue Kit (A48705, Beckman Coulter, samples extracted from 2021 onwards), following the manufacturer's instructions. All DNA samples were quantified using the fluorometric method (Qubit dsDNA BR Assay Kit, Life Technologies) and assessed for purity (260/280 and 260/230 ratios) by NanoDrop (Thermo Scientific). RNA extraction was performed using the miRNeasy Mini Kit (217004, Qiagen, samples extracted between 2018-2020), or the RNAdvance Tissue Kit (A32646, Beckman Coulter, samples extracted from 2021 onwards), following the manufacturer's instructions, and then treated with DNase I for 15 min at 30°C. RNA concentration and sample purity (260/280 and 260/230 ratios) were assessed using NanoDrop (Thermo Scientific). DNA and RNA integrity were checked with a TapeStation 4200 system using, respectively, Genomic DNA ScreenTapes and Reagents, and RNA ScreenTapes and Reagents, Agilent Technologies.

#### 1.2 Publicly available datasets

Additional datasets used in this study were obtained from the following publications: Peifer *et al.* 2012[4] (accession number: EGAS00001000925), Fernandez-Cuesta *et al.* 2014[3] (EGAS00001000650), George *et al.* 2015[5] (EGAS00001000925), George *et al.* 2018[6] (EGAS00001000708), Alcala *et al.* 2019[7] (EGAS00001003699), Laddha *et al.* 2019[8] (GSE118131), Miyanaga *et al.* 2020[9] (GSE142186), and Dayton *et al.* 2023[10] (EGAS00001005752). Data type and sample numbers are provided in the relevant data processing sections.

#### 1.3 Lung NET and lung NEN cohorts

To maximise our ability to characterise rare lung neuroendocrine tumours (lung NETs), the 201 samples from the lungNENomics cohort were combined with 115 lung NET samples from previous Computational Cancer Genomics team members publications (Fernandez-Cuesta *et al.* 2014, Alcala *et al.* 2019 and Dayton *et al.* 2023), and two previously published lung NETs studies (Laddha *et al.* 2019 and Miyanaga *et al.* 2020) which clustered with LCNEC (supra-carcinoids, see **Section 6.1**). Unless otherwise stated, all analyses were performed on this combined study cohort, subsequently referred to as the lung NET cohort (**Supplementary Table S1**). To investigate associations with lung neuroendocrine carcinomas, the lung NET cohort was combined with 73 large cell neuroendocrine carcinomas (LCNEC) from George *et al.* 2018 to form the lung NEN cohort. See **Supplementary Fig. S1**, **and Supplementary Tables S1-S3** for a summary of the lung NET and lung NEN cohorts.

##### *1.3.1 Examination of technical and clinical features of lung NET and lung NEN cohorts*

Technical and clinical features of interest were assessed for their statistically significant relationship with one another. Technical features examined were sample source (study), omics group (data type availability), whole-genome sequencing batch, RNA sequencing batch, DNA methylation array batch, Sentrix ID and Sentrix position, and tumour purity estimated by i) study pathologist (S.L.), ii) whole-genome sequencing (see **Section 2.11**), and iii) RNA sequencing (see **Section 3.6**). Clinical features examined were sex (inferred, see **Sections 2.3**, **3.3** and **5.3**), age category (continuous age values were cut into three groups: (15.9 – 40.7], (40.7 – 65.3], (65.3 – 90.1]), type (see **Section 1.1.1**), tumour location, stage, recurrence, smoking status, history of asbestos exposure, history of cancer, history of radiation exposure, and history of neuroendocrine genetic disorder. Unless otherwise stated, the associations between categorical variables were assessed with Fisher's exact tests, and between continuous and categorical variables with linear regression. Variables were grouped by theme (categorical technical, continuous technical, categorical clinical, and continuous clinical) for statistical analysis then adjusted for multiple testing within each group using the Benjamini & Hochberg method[11]. Results can be found in **Supplementary Table S2**.

***1.3.2 Survival analysis of lung NET and lung NEN cohorts***

Clinical features were also tested for their association with patient survival. Cox proportional hazards models were used to estimate the hazard ratio of each feature with regard to overall and event-free survival. For the lung NET cohort, clinical features tested were inferred sex, age category, type, tumour location, stage, recurrence, smoking status, history of asbestos exposure, and history of cancer. Insufficient sample numbers were available to test history of radiation exposure or history of neuroendocrine genetic disorder. For LCNEC samples, clinical features tested were sex, age category, stage and smoking status, no other variables were available.

For overall survival, death from disease or unknown cause was considered an event, whilst death from non-disease related known causes, and survival, were labelled as ‘no event’. For event-free survival, death from disease or unknown cause, or tumour recurrence (at the primary site or elsewhere) were considered events, whilst no recurrence during the study period was labelled ‘no event’, patients who had no reported tumour recurrence but who died of an unrelated cause were censored on the date of unrelated death and labelled ‘no event’. Patients were censored on the date of most recent follow up, or day of unrelated death, time from date of diagnosis to event or censor was calculated in months. Results can be found in **Supplementary Table S3**.

#### 2. Whole-genome sequencing

##### 2.1 Sample preparation and sequencing of the lungNENomics cohort

Whole-genome sequencing (WGS) was performed by the Centre National de Recherche en Génomique Humaine (CNRGH, Institut de Biologie François Jacob, Commissariat à l'énergie atomique et aux énergies alternatives) on 106 fresh-frozen lung NETs and their matched adjacent normal tissue or blood samples (from 72 patients). Following extraction, genomic DNA (1 µg) was used to prepare a library for whole-genome sequencing, using the TruSeq DNA PCR-Free Library Preparation Kit (20015963; Illumina), according to the manufacturer’s instructions. After quality control and normalisation, libraries were sequenced to a target depth of 60x for tumour tissues and 30x for matched normal tissue or blood on a HiSeqX5 platform (Illumina) as paired-end 150 bp reads. Sequence quality parameters were assessed throughout the sequencing run and standard bioinformatics analysis of sequencing data was based on the Illumina pipeline to generate FASTQ files for each sample.

##### 2.2 Data processing

WGS reads were mapped to the reference genome GRCh38 (with ALT and decoy contigs) by the CNRGH platform. In summary, the workflow consists of four steps: read mapping (software BWA; v0.7.15-r1140), duplicate marking and reads sorting (software sambamba; v0.6.8-pre1).

Alignment (CRAM) files from previously published WGS datasets were processed following the same procedure using our in-house version of the workflow (https://github.com/IARCbioinfo/alignment-nf v1.0; $n = 26$ lung NETs from Fernandez-Cuesta *et al.* 2014; and $n = 4$ lung NETs and $n = 2$ LCNECs from Dayton *et al.* 2023).

#### 2.3 Sex inference from whole-genome sequencing data

Sample sex was predicted from WGS using the sex determination step in PURPLE, as described in: https://github.com/hartwigmedical/hmftools/blob/master/purple/README.md#1-sex-determination. Two lung NET samples were found to be discrepant between clinically reported sex and WGS-predicted sex, S01060_B_TU (lungNENomics) and S01539 (Fernandez-Cuesta *et al.* 2014); however inspection of X and Y chromosome coverage, obtained from the PURPLE implementation of COBALT, indicated these samples had low coverage over the Y chromosome (**Supplementary Fig. S22**).

#### 2.4 Small variant calling

We called somatic single nucleotide variants (SNVs) using Mutect2 from GATK (v4.2.0), and Indels and multi-nucleotide polymorphisms (MNPs) using both Mutect2 and Strelka2 (v2.9.10), retaining only Indels and MNPs detected by both methods to avoid false discoveries that are more common in these variants, as previously described in Alcala *et al.* 2024[12]. Germline variants were called with Strelka2 only (v2.9.10). See workflows https://github.com/IARCbioinfo/mutect-nf release v2.3 and https://github.com/IARCbioinfo/strelka2-nf release v1.2a. We checked using mutational signature decomposition that no known artefactual signatures were present (see **Section 2.11**). Note that no MNPs were present in the intersection of Mutect2 and Strelka2 calls.

#### 2.5 Structural variant calling

We called somatic and germline structural variants using our workflow https://github.com/IARCbioinfo/sv_somatic_cns-nf v1.1, which uses an ensemble approach combining three structural variant callers (DELLY, Manta, and SvABA; see Di Genova *et al.* 2022[13]). In addition, we created a panel of normal SVs from the germline SVs detected in the normal samples by the three callers, and filtered out somatic SVs whose breakpoints both fall within a 100bp region of a germline SV in more than 1% of normal samples.

#### 2.6 Copy number variant calling

Copy number variants (CNVs) were called using PURPLE using our Nextflow pipeline iarcbioinfo/purple-nf v1.1, using a list of high-quality somatic small variants to improve the calls. Additionally, PURPLE estimated tumour purity, ploidy (including whole-genome duplication status, WGD, and microsatellite stability status, MSI). Following Mangiante *et al.* 2023[14], we rounded negative copy number estimates greater than -0.50 to 0 (6 out of 12443 segments) and removed those less than or equal to -0.5 (2 out of 12449 segments, less than 0.02%). Copy number values by segment and by gene are provided in **Supplementary Tables S18 and S19**. We computed CNV profiles using aCNViewer and ran GISTIC2 (v2.0.23) through the aCNViewer wrapper with confidence of 0.99 and broad event length of 0.7. MSI statuses were confirmed using MSIsensor-pro[15] (v1.2.0) in tumour-normal pair mode, which detected no sample with more than 0.5% of altered MSI sites. Sample purity, ploidy, WGD and MSI statuses and copy number values for significantly altered broad and focal events are provided in **Supplementary Table S17**.

##### *2.6.1 Timing of amplifications*

Amplifications were timed based on the allelic fractions of small variants and their amounts using R package mutationtimeR.

#### 2.7 Shattered regions detection

Spearman correlation between SV and CNV break counts was 0.57 across all samples, indicating good concordance between SV and CNV calls. We detected shattered chromosomal regions consistent with chromothripsis or chromoplexy using the svpluscnv R package (v0.9.1), which combines somatic CNV segment breakpoints (see **Section 2.6**) and structural variant breakpoints to identify regions with clustered breakpoints. We combined three sets of parameters to detect shattered regions. We used thresholds corresponding to (i) intermediate concordant evidence from the two types of variants (at least three CNV breaks and three SV breaks within a region), (ii) strong evidence from SVs but lower evidence from CNVs (at least one CNV and 20 SV breaks in a region), and (iii) strong evidence from CNVs but lower evidence from SVs (at least 20 CNVs and one SV in a region). We then used a threshold of seven CNV breaks to separate high- and low-confidence regions following recent practices[14]. Results are provided in **Supplementary Table S17**.

#### 2.8 Copy number, and small and structural variant burden computation

We computed mutational burdens for each type of alteration. For small variants and SVs, they correspond to the total number of such variants in each sample. For CNVs, we separated amplified and deleted segments, counting only those with an integer copy number different from 2 in autosomes and sex chromosomes in females, and different from 1 in Y chromosomes, and excluding samples with whole-genome duplication (according to software purple) in order to focus on arm-level and focal copy number changes. We tested the differences in burdens for small variants and SVs using pairwise t-tests on $\log_{10}(\text{burden}+1)$ values because of the spread of the burden distribution across several orders of magnitude. For deletions and amplifications, we compared the proportion of the genome either amplified or deleted, and favoured non-parametric tests (permutation tests; lmp function from R package lmPerm) because of the skew in the distribution of mutational burdens and the number observations tied at zero. In order to test simultaneously the differences between molecular groups and histological types, we computed models where the histological type variable was nested within the molecular group variable, using either linear (for small variants and SVs) or permutation models with function lmp (for deletions and amplifications). Results are presented in **Supplementary Table S20**.

In order to assess whether the burdens of the different types of variants were high or low, we compared them to that of common cancers using data from the Pancancer Analysis of Whole Genomes (PCAWG) consortium[16], following[14]. We downloaded the PCAWG somatic variant data following the instructions at https://docs.icgc-argo.org/docs/data-access/icgc-25k-data#open-release-data---object-bucket-details (release of August 2016 for metadata: sample sheet v1.4 and specimen histology v9), keeping only samples from the white list, and cohorts with at least 30 samples. Burdens were computed as described above for our own cohorts, taking the median value for each cohort to compare with that of lung NET molecular groups (results are provided in **Supplementary Figure. S8**).

#### 2.9 Driver mutation detection

Identification of cancer driver genes was performed with IntOGen[17]. IntOGen combines multiple driver detection methods to distinguish signals of positive selection from neutral mutagenesis across a cohort of tumours. The IntOGen pipeline was run on small variants and Indels identified in the lung NET cohort ($n$ = 102 individual patients with WGS data, see **Section 2.1**). The ten drivers which passed a filter of being expressed at > 1 TPM in 80% of the lung NET cohort were included for further analysis; excluded drivers were *FAM47C*, *FAT4*, and *MUC16*. A list of driver genes and altered samples are provided in **Supplementary Table S23**.

#### 2.10 Evolutionary trajectory inference

Recurrent evolutionary trajectories were identified using the R package revolver. We used as input all samples with WGS data available, including all regions sequenced whenever available, and used driver small variant and CNV alterations along with their inferred clonality. For CNVs, we considered all segments with a copy number between X-0.2 and X+0.2 to be clonal with copy number X, and otherwise to be subclonal. In order to separate the effect of WGD from that of focal amplifications, following what is performed by programs ACNViewer and GISTIC2, we considered amplifications and deletions relative to the ploidy of the sample, and in order to reduce noise in the trajectory calling, we removed from the analyses samples with uncertain WGD status (i.e., samples with ploidy confidence intervals including several ploidy values). Note that samples without any detected driver (small variant or CNV) are not taken into account by the algorithm, thus leading to 49 patients included in the analysis. Drivers with less than two alterations were also removed, leading to 18 driver events for 227 variants. Significant driver-to-driver trajectories were assessed using Fisher's exact test, and *q* values were computed using the Benjamini-Hochberg method.

#### 2.11 Mutational signatures detection

We computed SBS, DBS, INDEL, CNV, and SV signatures using SigProfilerExtractor (v1.1.21) on the lung NEN cohort ($n$ = 111 samples with WGS available). We tested from one to five *de novo* signatures with 250 replicates, and the optimal number of signatures was selected automatically by SigProfilerExtractor as the best compromise between maximising average signature stability and minimising mean sample cosine distance. For SBS, DBS, INDEL, and CNV signatures, COSMIC signatures were available, and each *de novo* signature was decomposed into COSMIC v3.3 signatures using default parameters. *De novo* signatures could be decomposed into 11 COSMIC signatures. For CNVs, we used rounded copy number estimates as done by default for PURPLE by subprogram SigProfilerMatrixGenerator, joining consecutive segments with similar rounded copy numbers to avoid over segmentation. For SVs, we used Signal signatures[18], as no COSMIC signatures are available. All results are reported in **Supplementary Table S21**. To confirm that no tumour presented a homologous recombination deficiency (HRD, absence of signatures SBS3, ID6, and SV3), we ran R package R CHORD[19] (v.0.9.1), which combines single base substitutions, indels, and structural variant information to identify HRD (**Supplementary Table S22**). Statistical differences among molecular groups and among histological types were conducted using fisher's exact test to test for an enrichment in presence/absence of certain signatures, and using linear regression model of the log10 number of alterations with age, sex, type and molecular groups as covariables to test for differences among samples where the signature is present (**Supplementary Table S21**).

We also performed signature variability analyses (Sigvar) as recently described[20]. We computed the mean within-sample signature diversity statistic (Gini-Simpson index; **Supplementary Fig. S21**, **Supplementary Table S21**) and compared the values between groups using t-tests.

To compute the signature most likely to be responsible for each small variant driver (**Extended Data Fig. 2e**), we assigned to each driver a mutation class (among the 96 SBS classes for SNVs and among the 83 ID classes for indels) and then computed the probability that a mutation of that class was generated by each mutational signature given the relative attributions of the signatures in the focal sample, following Morrison *et al.*[20]. The resulting probabilities were summed across samples from each group to produce **Extended Data Fig. 2e**.

#### 2.12 Neoantigen detection

Neoantigens were detected using pVACtools[21] as implemented within our nextflow pipeline hla-neo-nf (https://github.com/IARCbioinfo/hla-neo-nf). The level of immunoediting was estimated by the ratio of nonsynonymous to synonymous mutation rate (dN/dS) within neoantigen-rich regions, obtained using the SOPRANO method[22]. The diversity of HLA regions was computed using the Grantham distance (function HLADiversityScore from R package HLAdivR).

#### 2.13 Identification of pathogenic germline small variants

Small variants were called from germline WGS using Strelka2 (v2.9.10). VCF files were then annotated for their pathogenicity using the software InterVar[23] (v2.2.1). We selected variants labelled 'likely pathogenic' and 'pathogenic' from column 'InterVar: InterVar and Evidence', which resulted in 805 likely pathogenic/pathogenic germline variants for 102 patients (**Supplementary Table S26**). Variants were further filtered for relevance to cancer using an in-house gene list compiled from i) the National Center for Tumor Diseases/German Cancer Consortium (NCT/DKTK) Molecularly Aided Stratification for Tumor Eradication Research (MASTER) trial[24], and ii) peer-reviewed journal articles covering exome/genome sequencing of lung neuroendocrine neoplasms[3–9,25–28]. The MASTER trial aimed to investigate the clinical value of exome/genome sequencing in cancer care and included the evaluation of germline variants associated with genetic cancer predisposition syndromes. As such, 142 genes associated or potentially associated with cancer predisposition were selected based on expert opinion, in-house lists, and peer-reviewed literature, for germline analysis. To this we added 89 genes reported in the literature as being recurrently altered (two or more samples altered) in lung neuroendocrine neoplasms (**Supplementary Table S26**).

#### 2.14 Identification of damaging small and structural variants

##### *2.14.1 Filtering for damaging small variants*

Small variant calls for all samples were combined into a single dataset, annotated with ANNOVAR (v2020-06-08), and filtered to retain only likely damaging alterations as follows: (i) variants on chromosome M were removed, (ii) variants not labelled "exonic", "exonic;splicing", "splicing", and "ncRNA_exonic;splicing" (annotation column *Func.ensGene*) were removed, (iii) variants labelled "synonymous SNV" (annotation column *ExonicFunc.ensGene*), and (iv) variants labelled "silent" (function coding_change.pl), were removed, and finally (v) variants in non-protein coding or lncRNA genes were removed.

Variants were subsequently categorised with maftools (v2.10.05) into Frameshift Indel, In-frame Indel, lncRNA, Missense, Nonsense, Nonstop, Splice Site and Translation Start Site using labels provided in maftools column *Variant_Classification*. Variants labelled as 'Unknown' were manually examined for changes to amino acid sequence and re-categorised ($n$ = 4) or discarded ($n$ = 1) as appropriate. All damaging small variants are provided in **Supplementary Table S15**.

##### *2.14.2 Filtering for damaging structural variants*

We followed the approach from Mangiante *et al.* 2023 to classify SVs as damaging based on their type, and position of the breakpoints (in exon, introns, or intergenic regions). See script at https://github.com/IARCbioinfo/sv_somatic_cns-nf/blob/929dc35e14f6c9813747a7c7d223aa2fb2f32fe8/aux_scripts/SV_annotation.R. All damaging structural variants are provided in **Supplementary Table S16**.

#### 2.15 Enrichment testing of damaging small variants

Genes affected by at least one damaging small variant within the lung NET cohort were tested for enrichment for epigenetic regulatory genes using a gene list compiled by Halaburkova *et al.*[29] (**Supplementary Table S24**). The list of genes which were never affected by a damaging small variant in the lung NET cohort was used as background, and enrichment was tested using Fisher's exact test. Genes that were affected by both damaging and non-damaging small variants were only retained in the test list and were excluded from background.

#### 2.16 Genomic hallmarks of cancer analysis

##### *2.16.1 Datasets required*

The dataset of the 10 hallmarks of cancer given in Hanahan *et al.* 2022[30] used for this study and the corresponding genes that cause their acquisition through mutation were downloaded from the Catalogue of Somatic Mutations in Cancer (COSMIC, v99 GRCh38). This dataset was filtered to only contain cell types associated with human malignant cancers.

The dataset used for the genomic hallmarks of cancer analysis (**Supplementary Table S25**) was created by combining three data frames: Damaging small variants, ParetoTI analysis, and hallmarks of cancer from COSMIC. The resulting combination of these data frames gave us the hallmarks of cancer affected by each damaging mutation for a given sample, as well as the corresponding molecular group.

##### *2.16.2 Distribution of hallmarks affected per patient*

Using the previously mentioned dataset, a data frame in wide format was created, attributing to each sample the presence or absence of an effect from a damaging small variant for a given hallmark (**Supplementary Table S25**). Distributions of hallmarks affected per patient were obtained by computing row sums using the rowSums R function from the base package (v4.4.1) and plotting the results, grouped by molecular group, in a violin plot (ggplot2 package v3.5.1). The resulting four distributions were statistically compared two by two using Mann-Whitney U tests (*wilcox.test* function from the stats package v4.4.1), and significantly different distributions were annotated on the violin plot.

The same analysis was performed to compare histological types (typical versus atypical) within molecular groups and overall. As there were no typical tumours with WGS data within the sc-enriched group, this molecular group was excluded so as to not drive an increase in the number of hallmarks acquired in atypical samples overall.

##### *2.16.3 Genomic hallmark profiles*

An average genomic hallmark profile was created for each molecular group using the same data frame in wide format as in the previous section. Such a profile represents the proportion of patients for which each hallmark is affected by damaging SNVs. These proportions (one proportion per hallmark, ten per profile) were obtained by dividing the column sums (colSums function from the base package v4.4.1) by the number of samples for a given molecular group. Each profile was plotted as a bar plot (ggplot2 package v3.5.1).

To statistically compare each proportion between molecular groups, logistic regression models were used (one per hallmark; glm function from the stats package v4.4.1) to evaluate the association between each molecular group and a given hallmark. Ca A1 was used as the reference for each model. Corresponding forest plots were obtained for each model using the forest_model function from the forestmodel package (v0.6.2). However, hallmarks ‘angiogenesis’ and ‘proliferative signalling’ were not evaluated using such a model, as their proportions in the sc-enriched group were respectively 0 and 1 (which both introduce a null variance in a logistic regression model). Consequently, these two hallmarks were statistically evaluated between molecular groups using a Fisher’s exact test on each molecular group pair (fisher.test function from the stats package v4.4.1). Hallmarks with significant differences compared with Ca A1 were annotated on each bar plot. All statistical results are provided in **Supplementary Table S25**).

###### *2.16.4 Euler diagram of genes involved in hallmark acquisition*
A Euler diagram was created showing how many genes are involved in hallmark acquisition per molecular group, as well as how many are shared between molecular groups. The Euler function from package eulerr (v7.0.2) was used to create and plot the diagram. Genes that affect hallmarks in at least two patients were annotated on the diagram.

#### 3. Bulk RNA sequencing

##### 3.1 Sample preparation and sequencing of the lungNENomics cohort
RNA sequencing (RNA-seq) was performed at the Cologne Center for Genomics on 246 lung neuroendocrine tumours (from 180 patients). Following extraction, 1 µg total RNA was used for library preparation with the TruSeq mRNA stranded sample preparation kit (20020595; Illumina). After poly-A selection (using poly-T oligo-attached magnetic beads), mRNA was purified and fragmented using divalent cations under elevated temperature. RNA fragments underwent reverse transcription using random primers, followed by second strand complementary DNA (cDNA) synthesis with DNA Polymerase I and RNAse H. After end repair and A-tailing, indexing adapters were ligated. Products were then purified and amplified (14 PCR cycles) to create final cDNA libraries. After library validation and quantification (TapeStation, Agilent Biotechnologies), equimolar amounts of the library were pooled. The pool was quantified using a KAPA Library Quantification Kit (KK4835; Peqlab) and the 7900HT Sequence Detection System (Applied Biosystems). The pool was sequenced using an Illumina NovaSeq 6000 and a paired-end 100 nt protocol.

##### 3.2 Data processing
Reads were trimmed for the adapter sequence using Trim Galore (v0.6.5 for expression quantification, and v0.4.2 for alternative splicing analyses), then mapped to reference genome GRCh38 (using annotation gencode v33) with STAR software (v2.7.3a). Reads were realigned locally using ABRA2 (workflow https://github.com/IARCbioinfo/abra-nf release v3.0), and base quality scores were recalibrated using GATK (workflow https://github.com/IARCbioinfo/BQSR-nf release v1.1). Expression was quantified for each sample, generating a raw read count table with gene-level quantification for each gene of the comprehensive gencode gene annotation file (v33), as well as a table with Transcripts per Million (TPM), and Fragments per Kilobase per Million (FPKM), using StringTie software (v2.1.2) (Nextflow pipeline accessible at https://github.com/IARCbioinfo/RNAseq-transcript-nf release v2.2). Quality control was performed at each step. FastQC software (v0.11.9; https://www.bioinformatics.babraham.ac.uk/projects/fastqc/) was used to check raw reads quality and RSeQC software (v3.0.1) was used to check alignment quality.

FASTQ files from previously published RNA-seq datasets were processed following the same procedure ($n$ = 66 lung NETs from Fernandez-Cuesta *et al.* 2014; $n$ = 51 small cell lung carcinoma (SCLC) from Peifer *et al.* 2012 and George *et al.* 2015; $n$ = 69 LCNECs from George *et al.* 2018; $n$ = 20 lung NETs from Alcala *et al.* 2019; $n$ = 30 lung NETs from Laddha *et al.* 2019; *n* = 6 lung NETs from Miyanaga *et al.* 2020; and *n* = 7 lung NETs and *n* = 2 LCNECs from Dayton *et al.* 2023).

Subsequently, the raw gene count matrices and TPM matrices for all lungNENomics and publicly available datasets were combined into single gene count and TPM matrices. The gene count matrix was then variance-stabilised (R package DESeq2, v1.34.0) for use in statistical analyses. Two technical replicates were then removed from the matrices before further analyses (LNEN154_TU1_R2 and LNEN171_TU1_R2, both lungNENomics series).

##### 3.3 Sex inference from RNA sequencing data

Expression levels obtained from RNA-seq data on sex chromosomes were examined to identify any samples which did not cluster with others of the same clinical sex. A comparison of the sum of variance stabilised read counts on the X and Y chromosomes per patient identified five outlier samples (**Supplementary Fig. S22**). Samples S02236 (George *et al.* 2018), and LNEN199_TU (lungNENomics) were clinically reported as male but were predicted to be female by both RNA-seq and DNA methylation array data (**Section 5.3**), therefore all clinical data entries for these samples were replaced with NA in case the information had been entered erroneously. Sample SRR10720229 (Miyanaga *et al.* 2020) is also reported to be male but clustered with female samples on RNA-seq data. No WGS or DNA methylation array data was available for this sample to confirm tumour-specific loss of chromosome Y, therefore it was retained as male. Lastly, samples LNEN246_TU and LNEN251_TU1 (lungNENomics) were reported as female and male, respectively but did not cluster with samples of the same sex over X/Y chromosome expression. However, as their sex predicted by DNA methylation array matched their clinical sex (**Section 5.3**), RNA-seq data was discarded for subsequent analyses in case the RNA sample did not correspond to the correct patient ID. Finally, sample LNET19T (Dayton *et al.* 2023) had no clinically reported sex but was inferred to be female based on RNA-seq data.

##### 3.4 Fusion genes identification

Fusion genes were computed using STAR-fusion[31] and Arriba[32] (nextflow workflows https://github.com/IARCbioinfo/RNAseq-fusion-nf v1.1 and https://github.com/IARCbioinfo/gene-fusions-nf v1.1, respectively). Our STAR-fusion workflow follows the STAR-fusion best practices and relies on fusion inspector for validation. Our Arriba workflow was run both with and without providing structural variant calls for samples with WGS (option -d). In both cases, Arriba did not find any high-confidence fusion and only found 7 medium-confidence fusions, and the intersection with STAR-fusion detected fusions was null. This is in line with the very few structural variants detected in coding regions, and with the fact that most driver alterations affected tumor suppressor genes, which are not expected to be visible at the RNA level, rather than oncogenes, which would have led to highly expressed, high-confidence fusions.

##### 3.5 UMAP of neuroendocrine neoplasms

Data from sex and mitochondrial chromosomes were removed from raw gene count matrices obtained from bulk RNA-seq data (*n* = 59,607 genes) from 634 samples including 273 lung NETs (109 Ca A1, 89 Ca A2, 62 Ca B, 13 sc-enriched), 69 LCNEC, 51 SCLC, 135 pancreatic NETs, 88 small intestine NETs and 18 rectal NETs. Read counts were then variance stabilised using DEseq2 R package (v1.40.2). The 50 most variable genes were then selected to get VST50 on which Uniform Manifold Approximation and Projection (UMAP) was performed, using umap R package (v0.2.10.0) with number of neighbours equal to 15 and all the other parameters set to the default value.

#### 3.6 Immune contexture deconvolution

The proportion of cells that belong to each of ten immune cell types (B cells, macrophages M1, macrophages M2, monocytes, neutrophils, NK cells, CD4+ T cells, CD8+ T cells, CD4+ regulatory T cells, and dendritic cells) were estimated from the RNA-seq data using softwares quanTIseq[33] (downloaded 14 September 2020) using our workflow for parallel processing of samples (https://github.com/IARCbioinfo/quantiseq-nf release v1.1), and R package immunedeconv, that runs multiple deconvolution methods (Sturm *et al.* 2019[34]). In line with the immunedeconv package recommendations, we chose ABIS, EPIC, ESTIMATE, MCPCounter, and MUSIC, because they allow inter-patient comparisons. Methods may not be directly comparable given they use different reference cell types, and not all allow for unknown cell types to be present. Thus, in order to compare the values while taking into account the variability due to the method we computed z-scores across samples for each method and cell type, and compared the values of the z-scores by fitting a linear mixed model using R package lmer, with the method as a random effect. See results of the test in **Supplementary Table S10.**

RNA-seq estimated purity was also obtained from quanTIseq as 1 - the sum of the ten immune cell type proportions. Estimated cell type proportions are provided in **Supplementary Table S9**.

#### 3.7 Immune archetype inference

We inferred immune archetypes using the 3-feature classification method from Combes *et al.* 2022[35]. To ensure all archetypes were represented we used cohort lung NEN plus SCLC, and LUAD and LUSC TCGA datasets. We reprocessed the TCGA datasets as described in **Section 3.2** to limit batch effects. Following Combes *et al.* for each of the three genes (features), we converted expression into expression scores corresponding to the percentile ranks across samples. We then performed unsupervised clustering as in the original paper (Louvain clustering, using 100 *k* nearest neighbours and a resolution of 0.5) of the samples based on their 3-feature percentile scores. Once clustered, following the original method, cluster labels from the 3-feature classification of Combes *et al.* were attributed based on the prevalence and distributions of the features within the cluster (visualised as violin plots of all clusters in each feature, **Supplementary Fig. S4**). See **Supplementary Fig. S4**, that was compared to Fig. 2A-C from Combes *et al*. Immune archetypes are provided in **Supplementary Table S11**.

#### 3.8 Estimation of T-cell inflamed/pembrolizumab-responder phenotype

Mean expression (vst) of a panel of 18 genes, obtained from Ayers *et al.*[36], was calculated per sample. Expression levels were compared between molecular groups by t-tests. Results are presented in **Supplementary Table S34**.

#### 3.9 Calculation of SCLC-I score

To generate the SCLC Immune (SCLC-I) score and thus identify samples with an SCLC-I-like profile, we computed the mean expression values for 13 genes highly expressed by SCLC-I tumours in Gay *et al.*[37] (*CD274*, *PDCD1*, *CD80*, *CD86*, *CTLA4*, *CD38*, *IDO1*, *TIGIT*, *VSIR*, *ICOS*, *LAG3*, *CCL5*, and *CXCL10*). Mean variance stabilised expression was used to perform statistical analysis displayed in **Extended Data Fig. 3g**, and mean TPM expression was used for illustration (**Extended Data Figs. 3f,g**).

#### 3.10 Deconvolution of pulmonary neuroendocrine cell states

To characterise the pulmonary epithelial cell type composition of bulk RNA-seq samples we applied MuSiC (Multi-subject Single-cell Deconvolution), a method designed to infer cell type compositions from bulk RNA-seq data with cell-type specific gene expression from scRNA-seq reference data[38]. As reference profiles we used epithelial cells from foetal lung tissue airway organoids[39], enriched for lower airway progenitor and pulmonary neuroendocrine cells[40], as well as tumour microenvironment cells (stromal and immune) from lung neuroendocrine tumours[41]. The NE cells in this dataset (NE early 1, NE early 2, NE terminally differentiated 1 and 2) are represented by four cell states, corresponding to different levels of differentiation. Deconvolution was performed using raw gene count matrices from bulk RNA-seq data ($n$ = 273 lung NETs, $n$ = 69 LCNEC, $n$ = 51 SCLC, and one patient-derived tumour organoid (PDTO) from patient LNET10 from Dayton *et al.*[10]), and the scRNA-seq reference matrix. To reduce technical noise and increase robustness, genes expressed in fewer than three single cells were excluded from the analysis, as such low expression is typically too sparse to provide informative signal. Cell proportions are provided in **Supplementary Table S9**.

#### 3.11 Identification and analysis of molecular group core genes

##### *3.11.1 Identification of molecular group core genes*

Core upregulated and downregulated genes were identified for each molecular group of lung NETs using linear regression as follows. Variance stabilised expression values for autosomal protein-coding or lncRNA genes expressed at TPM ≥ 1 in at least two samples ($n$ = 25,804 genes, $n$ = 273 samples), were used to perform linear regression. For each gene and for each molecular group a linear model was built to calculate the relationship between the expression $g$ of the focal gene and the proportion $x$ of the focal archetype, such as $g(x) = \beta_0 + \beta_1 x + \epsilon$. The intercept $\beta_0 = g(0)$ corresponds to the estimated expression of the focal gene in samples with no proportion of the focal archetype, and the slope $\beta_1 = g(1) - \beta_0$ corresponds to the difference in the expression of the focal gene in samples purely belonging to the focal archetype compared to samples with no proportion of this archetype. $P$ values were adjusted for multiple testing using the Benjamini & Hochberg method, and $\log_2$ fold changes were calculated as follows: $\log2(\frac{\beta_0+\beta_1}{\beta_0})$.

For each molecular group, genes were then assigned a label of ‘positive’ ($q$ value < 0.05 and $\log_2$ fold change > 1), ‘negative’ ($q$ value < 0.05 and $\log_2$ fold change < -1), or ‘false’ ($q$ value ≥ 0.05 and/or -1 < $\log_2$ fold change < 1). Finally, each gene was assigned as a core upregulated or downregulated for a molecular group if it fulfilled the following criteria: a core upregulated gene in molecular group $x$ must be labelled ‘positive’ for group $x$ and labelled either ‘negative’ or ‘false’ for the three other groups, and *vice versa* for a core downregulated gene. As such, no gene can be a core upregulated gene for more than one molecular group, nor a core downregulated gene for more than one molecular group. Core upregulated and downregulated genes are shown in **Supplementary Table S27**.

##### *3.11.2 Identification of key Gene Ontology pathways*

The Gene Ontology biological processes for human genes database was downloaded to identify significantly enriched pathways among the core up-regulated genes specific to each molecular group (Fisher’s exact test). To summarise the numerous core pathways identified, we constructed directed weighted graphs, where edges were drawn from the pathway containing the most genes to the one containing fewer, with edge weights corresponding to the Jaccard index between the two gene lists. We then applied the Walktrap community detection algorithm to define clusters of pathways involved in similar processes. Finally, each cluster of pathways was named based on the most central pathway according to weighted out-degree values, forming what we term a ‘super-pathway’ (**Fig. 2c, Supplementary Table S28**).

*3.11.3 Identification of master transcription factors*

We used the CaCTS algorithm[42] (v1.0; https://github.com/lawrenson-lab/CaCTS) to determine potential master transcription factors (TFs) per molecular group. As suggested by Reddy *et al.* we used the 1671 master TFs identified by Saint-André *et al.*[43] and Lambert *et al.*[44], and considered as master TF genes those that are both in the top 5% of the most expressed genes and in the top 5% of genes with the highest Jensen-Shannon divergence scores.

*3.11.4 Testing for enrichment of hallmarks of cancer*

The integrated human gene set collection associated with the ten hallmarks of cancer proposed by Menyhart *et al.*[45] (downloaded from https://cancerhallmarks.com/download) was used to identify significantly enriched hallmarks among the core upregulated genes of each molecular group (Fisher's exact test). In **Extended Data Fig. 3e**, we report the number of sc-enriched core upregulated genes and the three genes with the highest fold change for each significant hallmark.

*3.11.5 Testing for enrichment of neuroendocrine cell genes*

Genes highly expressed in lung neuroendocrine (NE) cells were obtained from Travaglini *et al.*[46]. Enrichment testing was performed using Fisher's exact tests between target lists of core upregulated genes per molecular group against background of all other genes profiled with RNA sequencing (**Supplementary Tables S29 and S30**).

*3.11.6 Creation of the RNA sequencing heatmap (Fig. 2c)*

To illustrate the findings from this section, we selected a small subset of core genes or master TFs for each molecular group. For each molecular group, a list of genes was created which were core upregulated, within a super-pathway, and were expressed at median TPM > 1 within the molecular group. From these lists, genes were then selected manually based on previous relevant publications[7,8,28] or out of interest from literature[47–53]. Additionally, putative master TFs for each molecular group were plotted.

#### 3.12 Proliferation index

A measure of proliferation rate per sample was obtained from variance-stabilised read counts (R package DESeq2) as follows. A proliferation index score was calculated for each sample as the median expression of the top 1% of genes (131 genes) significantly positively correlated with the expression of *PCNA* (proliferating cell nuclear antigen) in 27 different healthy tissue types[54,55], **Supplementary Tables S1 and S35**). A higher proliferation index indicates greater proliferation. Associations between proliferation index and lung NET molecular group ($k = 4$) and type (typical versus atypical) were assessed with ANOVA and t-tests, respectively. Correlation tests between molecular group proportion and proliferation index were assessed with Pearson correlation. Results are shown in **Supplementary Table S33**.

#### 3.13 Epithelial-mesenchymal transition measurement

A score of epithelial-mesenchymal transition (EMT) per sample was calculated from variance-stabilized read counts as the mean expression of 52 mesenchymal-associated genes minus the mean expression of 25 epithelial-associated genes, as previously described[56], **Supplementary Tables S1 and S35**). A higher EMT score indicates a more mesenchymal-like gene expression profile than epithelial-like. Associations between EMT score and lung NET molecular group ($k = 4$) and type (typical versus atypical) were assessed with ANOVA and t-tests, respectively. Correlation tests between molecular group proportion and EMT score were assessed with Pearson correlation. Results are shown in **Supplementary Table S33**.

#### 3.14 Determination of Proneural, HNF+, or Luminal regulatory subtype

Gene sets characteristic of Proneural, HNF+ and Luminal regulatory subtypes were obtained from Davis *et al.*[57]. We selected *ASCL1*, *SOX4*, and *TCF4* for Proneural classification, *HNF1A*, *HNF4A*, *FOXA3*, *FGFR3*, and *FGFR4* for HNF+ classification, and *OTP* for Luminal classification. The bimodality of gene expression (variance stabilised expression values) was first assessed for each gene individually using Hartigan's Dip Test (R package diptest, v0.76-0) all were significantly bimodal except *SOX4* and *TCF4* ($P$ values ≥ 0.05), then correlation between genes within each set was tested by Pearson correlation (**Supplementary Table S31**). All genes within Proneural and HNF+ gene sets were significantly correlated with one another, and therefore a single gene was selected for classification of each subtype. For Proneural, we selected *ASCL1* as it was the only bimodal gene, and for HNF+ we selected *HNF1A* as it was specifically highlighted by Davis *et al.*

To determine sample regulatory subtype we first classified samples as being *ASCL1* high or low, *HNF1A* high or low, and *OTP* high or low. For this we followed the procedure described in Moonen *et al.*[58], first fitting two Gaussian mixture model distributions to the distribution of variance stabilised gene expression (R package mclust, v5.4.10), then selecting a cut-off as the lowest density point of the two Gaussian distributions (**Supplementary Table S31**). Cut-off values were as follows: ≥ 11.16856 for $ASCL1^{high}$, ≥ 6.247015 for $HNF1A^{high}$, and ≥ 11.8089 for $OTP^{high}$. Finally, a regulatory subtype was assigned using the following criteria: Proneural = $ASCL1^{high}/HNF1A^{low}/OTP^{high}$, HNF+ = $ASCL1^{low}$ $/HNF1A^{high}/OTP^{high}$, and Luminal = $ASCL1^{low}/HNF1A^{high}/OTP^{low}$. All samples which didn't fit these criteria were given the classification of Other (**Supplementary Table S31**).

#### 3.15 *TERT* expression analysis

A category of TERT high or low had previously been assigned to a subset of the lung NET cohort ($n$ = 76) with RNA-seq data as described in Werr *et al.*[59]. A $\log_2$ (sFPKM) cut-off of 8.17 was defined by Werr *et al.* to distinguish high from low TERT samples. To assign a TERT expression category to the entire lung NET cohort, we first examined the association between *TERT* $\log_2$(FPKM) obtained from internal processing pipelines (see **Section 3.2**) and values from Werr *et al.* Finding a significant correlation between the two measurements ($P$ value = 1.64 $\times 10^{-24}$, r = 0.95, Pearson correlation, note all samples with -Inf (as FPKM = 0) were excluded from correlation test). A linear model was then fit between the two *TERT* expression measurements, again excluding samples with –Inf, and the y-intercept for a $\log_2$(sFPKM) value of 8.17 was calculated using the model coefficients ($m$ = 0.9244959, $c$ = -12.5544998). The y-intercept was used as the internal $\log_2$(FPKM) *TERT* expression cut off to distinguish TERT expression categories, high expression was defined as $\log_2$(FPKM) ≥ -5.001368. Results are provided in **Supplementary Table S36**.

#### 3.16 Statistical analysis of individual gene expression levels

Expression of genes of interest (variance stabilised expression values) were directly compared between lung NET molecular groups ($k$ = 4) and tumour type (typical versus atypical) using ANOVA and t-tests, respectively. Genes were grouped by category (somatostatin receptors, hormone receptors, viral receptors) and adjusted for multiple testing using the Benjamini & Hochberg method[11]. Where comparisons were significant ($q$ value < 0.05) for molecular group, comparisons between pairs of groups were performed using t-tests, and where significant for both molecular group and type, comparisons between pairs of groups were further assessed within typical only and atypical only groups. Plotting of significant results was performed with $\log_{10}$(TPM + 1) expression values.

#### 4. Spatial transcriptomics

##### 4.1 Sample preparation and sequencing

Spatial transcriptomic sequencing of four FFPE samples was performed at Centre Léon Bérard using the 10x Genomics Visium v1 platform. Each sample was placed on a 10x Genomics Visium slide followed by deparaffinisation, H&E staining and decrosslinking steps, according to 10x Genomics guidelines. Human probes targeting approximately 18,000 genes were hybridised overnight on the slides and captured on each spot after ligation between the LHS and RHS probes. Libraries were produced for each sample following 10x Genomics protocols. Libraries were prepared and sequenced on an Illumina NovaSeq 6000 machine with a target sequencing depth of 50,000 reads per spot.

##### 4.2 Data processing

Samples were processed using SpaceRanger (v1.3.0). Data were demultiplexed, and reads mapped to reference genome GRCh38, and tissue and fiducial detection was performed before barcode/unique molecular identifier (UMI) counting, generating feature-barcode matrices. Quality controls of raw (percentage of valid barcodes and UMIs, quality scores) and processed data were performed (percentage of reads mapped, median counts per spot).

###### *4.2.1 Domain identification and spot deconvolution*

Data from the spatial spots was clustered across all samples simultaneously with cell type location estimation using the IRIS algorithm[60]. This allowed for checking whether domains were patient-specific or shared across samples. The reference single-cell profiles used as input to IRIS were the same as those used for the deconvolution of NE cell states within bulk RNA-seq data with MuSiC (**section 3.10**) in order to allow for their comparison, and to include potential neuroendocrine cells of origin in the computation.

###### *4.2.2 Computation of molecular group scores*

In order to estimate which spots had expression profiles resembling that of the lung NET molecular groups, we performed deconvolution of the expression of the spots (method CARD[61]) using the average expression profile of each molecular group as references, focusing on the list of core differentially expressed genes (see **section 3.11.1**).

###### *4.2.3 Spatial correlation analysis*

To assess the co-localisation of different cell types and molecular group profiles, as well as between expression of signalling genes and cell types/molecular group profiles, identified on FFPE slides, we computed bivariate spatial cross-correlation coefficients[62] and their *P* values using a permutation test without replacement (**Supplementary Figs. S20 and S21, Supplementary Table S47**).

#### 5. DNA methylation arrays

##### 5.1 Sample preparation and DNA methylation detection of the lungNENomics cohort

DNA methylation arrays were performed at the International Agency for Research on Cancer for 281 lung NETs (from 191 patients). Following DNA extraction, 600 ng of purified DNA was bisulphite-converted using the EZ-96 DNA Methylation kit (D5004; Zymo) following the manufacturer's recommendations for Infinium assays. Then, 250 ng of bisulphite-converted DNA was used for amplification, fragmentation, and hybridisation on Infinium

MethylationEPIC v1.0 BeadChips (WG-317-1003, Illumina), following the manufacturer's protocol. Chips were scanned using Illumina iScan to produce two-colour raw data files (IDAT format). Samples were assigned to chips using stratified randomisation to mitigate the batch effects. Samples were assigned to evenly distribute, in order of priority, histopathological type, provider, sex, smoking status, and age. ITH samples were placed on the same chip as their corresponding tumour sample. The position of samples on each chip was then randomised.

##### 5.2 Data processing

IDAT files from the lungNENomics cohort and an additional 76 ($n$ = 56 lung NETs, $n$ = 20 LCNECs) from Alcala *et al.* 2019 and George *et al.* 2018 were imported into the R statistical programming environment and processed using R packages minfi (v1.40) and ENmix (v1.30.03), following our standard workflow (https://github.com/IARCbioinfo/Methylation_analysis_scripts). Two-colour intensity data from internal control probes were manually inspected to check the quality of successive sample preparation steps (bisulphite conversion, hybridisation, extension, and staining; ENmix). All samples passed the QC steps of per sample $\log_2$ methylated and unmethylated chip-wise median signal intensity comparison (minfi), and overall p-detection value measurement (all $P$ values < 0.01, minfi).

Four groups of probes were removed: (i) poor performing probes with a p-detection value > 0.01 in at least one sample (41,279 probes discarded), p-detection value was computed by comparing the total signal (methylated and unmethylated) of each probe with the background signal level from non-negative control probes (minfi) (ii) cross-reactive probes (41,777 probes discarded), cross-reactive probes co-hybridise to multiple locations within the genome and therefore cannot be reliably investigated (iii) probes on the sex chromosomes (16,440 probes discarded), and (iv) probes with SNPs within the single base extension site, or target CpG site, at a minor allele frequency of > 5% (database dbSNP build 137, 7,510 probes discarded). This resulted in a normalised, filtered dataset of 758,853 probes for 357 samples. Beta and M-values were extracted (functions getBeta and getM, minfi), and probes recording M-values of $-\infty$ for at least one sample were replaced with the next lowest M-value in the dataset.

##### 5.3 Sex inference from DNA methylation array data

Sample sex was predicted from DNA methylation array data using a predictor based on the median total intensity on the X and Y chromosomes (function getSex, R package minfi, **Supplementary Fig. S22**). S00567 (George *et al.* 2018), LNEN164_TU, and LNEN258_TU (both lungNENomics) were clinically male but predicted female according to DNA methylation data. However, their RNA-seq profile was consistent with male sex and these samples were therefore retained as male. Samples S02236 (George *et al.* 2018) and LNEN199_TU (lungNENomics) were predicted to be female by both RNA-seq and DNA methylation array data (see **Section 3.3**), and clinically reported as male, therefore all clinical data entries for these samples were replaced with NA in case the information had been entered erroneously. Finally, sample LNEN028 (Alcala *et al.* 2019) had no clinically reported sex but was inferred to be male based on DNA methylation array data.

#### 6. Single-omic consensus clustering analyses

##### 6.1 Single-omic consensus clustering for identification of supra-carcinoids

Consensus clustering was performed for RNA-seq and DNA methylation array datasets separately to identify new instances of supra-carcinoids, defined as lung NETs clustering with LCNECs. Principal components analysis (PCA) was subsequently performed on each dataset

(R package ade4, v1.7-20, number of factors set to ten, data centred and unscaled) to visualise consensus clusters.

###### *6.1.1 RNA sequencing data*

The sample set (*n* = 284) comprised *n* = 179 lung NETs from lungNENomics, *n* = 30 lung NETs from Laddha *et al.* 2019, *n* = 6 lung NETs from Miyanaga *et al.* 2020, and *n* = 69 LCNEC from George *et al.* 2018. To generate the expression data for consensus clustering the variance stabilised read count matrix was subset to exclude genes on chromosomes X, Y and M, filtered to retain only genes with a minimum difference of ≥ 1 TPM across the sample set, then reduced to the top 5,000 genes by variance and median centred. Clustering with a k-means clustering algorithm based on Euclidean distances was repeated 100 times with random 80% subsampling to generate consensus clusters for *k* = 2-8 (R package ConsensusClusterPlus, v1.58). PCA was performed using the top 5,000 genes by variance (filtered as above) for visualisation. Results are presented in **Supplementary Table S4**.

###### *6.1.2 DNA methylation array data*

The sample set (*n* = 211) comprised *n* = 191 lung NETs from lungNENomics, and *n* = 20 LCNEC from previously published data (Alcala *et al.* 2019, George *et al.* 2018). To generate the M-value matrix for consensus clustering the matrix was filtered to retain only probes with a minimum difference of ≥ 0.1 beta value across the sample set, then reduced to the top 5,000 probes by variance and median centred. Clustering with a k-means clustering algorithm based on Euclidean distances was repeated 100 times with random 80% subsampling to generate consensus clusters for *k* = 2-8 (R package ConsensusClusterPlus, v1.58). PCA was performed using the top 5,000 probes by variance (filtered as above) for visualisation. Results are presented in **Supplementary Table S4**.

##### 6.2 Single-omic consensus clustering to examine the relationship between lung NETs and SCLC

Additional consensus clustering was performed by combining RNA-seq data from the lung NET and lung NEN cohorts with SCLC data from two previous publications (Peifer *et al.* 2012 and George *et al.* 2015). Three matrices were generated for three analyses: i) lung NEN cohort (*n* = 342 with RNA-seq data), ii) lung NET cohort + SCLC (*n* = 324, comprising lung NET cohort, 15 SCLC from Peifer *et al.* 2012 and 36 SCLC from George *et al.* 2015), and iii) lung NEN cohort + SCLC (*n* = 393).

To generate the expression data matrices for the three consensus clustering analyses, the following procedure was used per sample set: variance stabilised read count matrix was subset to exclude genes on chromosomes X, Y and M, filtered to retain only genes with a minimum difference of ≥ 1 TPM across the sample set, then reduced to the top 5,000 genes by variance and median centred. Clustering with a k-means clustering algorithm upon Euclidean distances was repeated 100 times with random 80% subsampling to generate consensus clusters for *K* =2-8 (R package ConsensusClusterPlus, v1.58). PCA was performed using the top 5,000 genes by variance (filtered as above) for visualisation. Results are presented in **Supplementary Table S4**.

The area of the convex hull formed by the most extreme points (samples) over principal component axes 1 and 2 was calculated for all samples in the analysis, and for lung NET-only samples, using R functions chull() and Polygon() (package sp v. 1.5-0).

### 7. Multi-omics factor analysis
Multi-Omics Factor Analysis (MOFA) was performed using software MOFA2[65] (v1.4.0) for two cohorts (i) lung NETs only (*n* = 319) and (ii) lung NETs plus LCNEC (*n* = 392). Cohort lung NET (i) comprised 201 newly sequenced lungNENomics samples, 116 samples from the previous Computational Cancer Genomics team publications Alcala *et al.* 2019 and Dayton *et al.* 2023, plus two previously published lung NETs which clustered with LCNEC (supra-carcinoids) (see **Section 6.1**) in order to maximise the number of rare subtypes available for molecular characterisation. Cohort lung NEN (ii) was composed of cohort lung NET (i) and an additional 73 LCNEC (from George *et al.* 2018).

#### 7.1 Pre-processing of RNA-seq data
For each cohort, variance stabilised raw gene counts (DESeq2, v1.34.0) for 273 and 342 samples (cohort lung NET and cohort lung NEN, respectively) were subset to exclude genes on chromosomes X, Y and M, filtered to retain genes with a minimum variance of TPM ≥ 1 across the cohort, then reduced to the top 5,000 genes by variance. Samples with no RNA-seq data were assigned NA values for each gene.

#### 7.2 Pre-processing of DNA methylation array data
For each cohort, M-values for 247 and 267 samples (cohort lung NET and cohort lung NEN, respectively) were filtered to retain probes with a minimum variance of beta value ≥ 0.1 across the cohort, then reduced to the top 5,000 probes by variance. Samples with no DNA methylation array data were assigned NA values for each probe.

#### 7.3 Pre-processing of small and structural variant data
Damaging small and structural variants for each cohort (see **Section 2.14**) were combined and filtered to exclude variants in genes that i) were lowly expressed (where maximum TPM < 0.01 per gene across the sample set), or no TPM values were available, and ii) were not altered in ≥ 2 samples. MOFA input matrices were then created by assigning a value of 0 (wild-type) or 1 (altered) to each sample per gene. Samples for which WGS data was available but had no damaging small or structural variants were included with a value of 0 for each gene, those with no WGS data were assigned values of NA for each gene.

#### 7.4 Pre-processing of copy number variant data
Copy number values for the eight significant broad and focal events as detected in the lung NET cohort by GISTIC2 (see **Section 2.6**) were included as a single input matrix. Samples with no WGS data were assigned values of NA for each event.

#### 7.5 Generating MOFA latent factors
Two MOFA runs were performed, one for each cohort (lung NET and lung NEN). Input datasets for each run comprised those described in **Sections 7.1-7.4** above, and the number of latent factors was set to ten. See https://github.com/IARCbioinfo/MS_lungNENomics for detailed method and input datasets. A summary of input data and MOFA runs, including QC, is shown in **Supplementary Figs. S2 and S23, Supplementary Table S5**. Sample coordinates along latent factors are provided in **Supplementary Table S4**.

##### *7.5.1 Measuring convex hull area over MOFA latent factors*
The area of the convex hull formed by the most extreme points (samples) over MOFA lung NEN latent factors 1 and 2 was calculated for all samples in the analysis, and for lung NET only samples, using R functions chull() and Polygon() (package sp v. 1.5-0).

##### 7.6 Intra-tumoural heterogeneity lung NET MOFAs

An additional 91 MOFA analyses of lung NETs were performed to measure how similar each ITH tumour piece was to its original sample. For each ITH sample, the values of the original sample within the four input matrices were replaced with the corresponding values from the ITH sample, keeping the rest of the matrices unchanged. Where an ITH piece had different data availability to the original sample, the values for the missing omic dataset were replaced by NA. Each ITH MOFA run was subsequently performed as described in **Section 7.5** to generate latent factor positions. In addition, ParetoTI analysis was performed (see **Section 8.1**) for each of the 91 ITH MOFA runs to determine the molecular group of each ITH region.

A measure of how similar within the latent factor space each ITH piece was to its original sample was obtained by calculating the Euclidean distance between the pairs of samples over latent factors 1, 2 and 5 (those used to determined sample molecular group, **Supplementary Table S48**). In the event that an ITH latent factor was the inverse of the original, the values for that latent factor were multiplied by -1 before the distance calculation was performed. Where an ITH latent factor order had changed, for instance original latent factor 5 was correlate d with ITH factor 4, the distance calculation was performed using the factors which significantly correlated with the original LFs 1, 2 and 5. Pairs where the Euclidean distance was greater than the mean distance between samples within the original sample molecular group were visualised over factors 1, 2 and 5. Euclidean distances were also used to calculate the silhouette statistic. Scores range from –1 to 1, where negative values indicate a sample is closer to samples from another molecular group than its own molecular group label. Group label was assigned to each ITH piece from a patient as the molecular group of the tumour region used in the lung NET cohort.

##### 7.7 Variable associations with MOFA latent factors

Technical and clinical features of interest were assessed for their statistically significant relationship with sample latent factor positions. Unless otherwise stated, the association between latent factor positions and continuous variables was assessed with Pearson correlation tests, and with categorical variables using linear regression. Variables were grouped by theme (categorical technical, continuous technical, and categorical clinical) for statistical analysis then adjusted for multiple testing within each group using the Benjamini & Hochberg method[11]. Overall and event-free survival associations with latent factor positions were assessed using the Cox proportional hazards model. Definitions of overall and event-free survival can be found in **Section 1.3.2**. Results can be found in **Supplementary Table S5**.

#### 8. Pareto task inference analysis

Molecular groups of samples were identified through the application of multi-task evolutionary theory by Pareto task inference (ParetoTI)[66,67] to the latent factors identified in each MOFA analysis (R package ParetoTI, v0.1.13). The ParetoTI algorithm fits a low-dimensional polytope over the samples as they are positioned within the multi-dimensional latent factor space, with the vertices of the polytope representing archetypes (or subgroups) of the data.

##### 8.1 ParetoTI analysis of MOFA lung NET

For ParetoTI analysis latent factors which were exclusively associated with technical features were excluded (LFs 4, 8 and 9, see **Supplementary Table S5**) then ordered by the proportion of variance in RNA-seq data explained (highest to lowest: 1, 2, 5, 6, 3, 10, 7, see **Supplementary Table S5**). The ParetoTI model was fitted over six combinations of successive latent factors (LFs 1, 2; LFs 1, 2, 5; LFs 1, 2, 5, 6; LFs 1, 2, 5, 6, 3; LFs 1, 2, 5, 6, 3, 10; and LFs 1, 2, 5, 6, 3, 10, 7), using 200 bootstraps with 75% subsampling, to generate polytopes with between two and six vertices (archetypes). See https://github.com/IARCbioinfo/lungNENomics_Archetype for detailed method. Metrics of how well each model performed, and the resulting archetype positions, are reported in **Supplementary Table S6**.

Performance metrics examined were (i) the t-ratio, i.e. the ratio of the volume of the best-fitting polytope to the volume of the convex hull of the data, (ii) the variance explained by each polytope, and (iii) the total variance in position of archetypes during bootstrapping. Each metric was also assessed for statistical significance, and a model was considered a significant fit if the $P$ value for all three metrics was < 0.05. We subsequently examined two significant fits: $k = 3$ generated over LFs 1 and 2, and $k = 4$ generated over LFs 1, 2 and 5.

###### *8.1.1 Archetype attribution and naming*

The proportion of each archetype was calculated per sample per fit, and the samples were assigned to the archetype for which they had the greatest proportion (**Supplementary Table S1**). Archetypes were then examined for enrichment of samples previously classified into the molecular groups of Ca A1, Ca A2, Ca B, and supra-carcinoid (Alcala *et al.* 2019, Dayton *et al.* 2023, **Supplementary Fig. S3**). Finally, archetypes were renamed as the molecular group to which they best corresponded as follows ($k = 4$): V1 = supra-carcinoid enriched, V2 = Ca A1, V3 = Ca B, V4 = Ca A2.

##### 8.2 ParetoTI analysis of MOFA lung NEN

For ParetoTI analysis, latent factors that were exclusively associated with technical features were excluded (LFs 5 and 10, see **Supplementary Table S5**) then ordered by the proportion of variance in RNA-seq data explained (highest to lowest: 2, 1, 3, 6, 7, 9, 4, 8, see **Supplementary Table S5**). The ParetoTI model was fitted over seven combinations of successive latent factors (LFs 2, 1; LFs 2, 1, 3; LFs 2, 1, 3, 6; LFs 2, 1, 3, 6, 7; LFs 2, 1, 3, 6, 7, 9; LFs 2, 1, 3, 6, 7, 9, 4; and LFs 2, 1, 3, 6, 7, 9, 4, 8), using 200 bootstraps with 75% subsampling, to generate polytopes with between two and six vertices (archetypes). See https://github.com/IARCbioinfo/lungNENomics_Archetype for detailed method. Metrics of how well each model performed, and the resulting archetype positions, are reported in **Supplementary Table S6**.

We subsequently examined two significant fits: $k = 3$ generated over LFs 1 and 2, and $k = 4$ generated over LFs 1, 2 and 3.

###### *8.2.1 Archetype attribution and naming*

The proportion of each archetype was calculated per sample per fit, and the samples were assigned to the archetype for which they had the greatest proportion (**Supplementary Table S1**). Archetypes were then examined for enrichment of samples previously classified into the molecular groups of Ca A1, Ca A2, Ca B, and supra-carcinoid (Alcala *et al.* 2019, Dayton *et al.* 2023, **Supplementary Fig. S24**). Finally, archetypes were renamed to match the molecular group to which they best corresponded as follows ($k = 4$): V1 = Ca A2, V2 = Ca A1, V3 = LCNEC, V4 = Ca B.

##### 8.3 Measuring molecular heterogeneity within molecular groups

Two methods were used to assess heterogeneity between molecular groups, Euclidean distances over MOFA LFs and variance across MOFA input datasets. The mean Euclidean distances between samples within the same molecular group over MOFA lung NET LFs 1, 2 and 5, and MOFA lung NEN LFs 1, 2 and 3, were calculated, providing a measure of the average spatial distance on the molecular map between samples of the same molecular group (**Supplementary Table S48**).

The variance of MOFA input datasets was obtained by first subsetting each input matrix to contain only samples of a single molecular group, resulting in four matrices per original input, calculating the variance by row (i.e. gene, probe, genome segment) within each sub-matrix, then averaging the variance of each sub-matrix (**Supplementary Table S48**).

##### 8.4 Variable associations with molecular groups

Technical and clinical features of interest were assessed for their statistically significant association with molecular groups. Unless otherwise stated, the association between molecular groups and categorical variables was assessed with Fisher's exact tests, and molecular groups with continuous variables was assessed using ANOVA, in the R statistical programming environment. Variables were grouped by theme (categorical technical, continuous technical, categorical clinical, categorical morphological) for statistical analysis then adjusted for multiple testing within each group using the Benjamini & Hochberg method[11]. Significant associations between a variable and molecular groups overall were then further examined between pairs of molecular groups using either Fisher's exact tests (for categorical variables) or two-tailed t-tests (for continuous variables). Additionally, Binomial tests (one-proportion z-tests) were used to test for enrichment within each molecular group for particular feature levels, for example, to determine whether there were more typical samples in Ca A1 than expected. Variables with greater than one third of the cohort missing data (history of cancer, history of radiation, recurrence, neuroendocrine genetic disorder, location, asbestos exposure, and smoking pack years) were assessed for differences in proportion of missing data between molecular groups. Results not presented in main figures can be found in **Supplementary Table S7**.

Kaplan-Meier survival estimates for molecular groups were calculated for both overall survival and event-free survival. Definitions of overall and event-free survival can be found in **Section 1.3.2**. Results not presented in main figures can be found in **Supplementary Table S8**.

#### 9. Deep learning histopathological analyses

##### 9.1 Image pre-processing

Haematoxylin/eosin (HE) or haematoxylin/eosin/saffron (HES) whole-slide images (WSIs) for 212 patients from the lungNENomics cohort (193 with molecular group data) were available for use in deep learning histopathological analysis. WSIs were cut into 384 x 384 pixel tiles, and those with more than 80% background pixels were excluded, resulting in a dataset of 3.5 million tiles. Tile colours were normalised using the colour deconvolution method proposed by Vahadane *et al.* 2016[68].

##### 9.2 Barlow Twins self-supervised deep learning model

To identify correlations between morphological features of lung NETs and their molecular profiles, we first trained a self-supervised network to generate similar representations for tiles with common morphological features. We reused the Barlow Twins models originally proposed by Zbontar *et al.*[69] and first applied to histological images by Quiros *et al.*[70]. The original Pytorch implementation of the model developed by Facebook Research was reused for this study (https://github.com/facebookresearch/barlowtwins). Two Wide ResNet-50 networks pre-trained on ImageNet were used as the Barlow Twins backbone. The model was trained on a subset of 300,000 randomly selected HE/HES tiles. A large batch size of 896 images was chosen as proposed by Zbontar *et al.* so we developed a parallel implementation of Barlow Twins running simultaneously on 8 NVIDIA V100 GPUs with 16 GB of RAM each. After training for 240 epochs, Barlow Twins was used to create a reduced representation of the 3.5 million tiles, generating 128-dimensional vectors for each input image. Similar vectors are assumed to have similar morphological features, and *vice versa* for dissimilar vectors.

#### 9.3 Computation of Leiden morphological partitions on Barlow Twins encoded vectors

A random set of 200,000 encoded vectors generated by Barlow Twins was used to create morphological partitions based on the Leiden clustering algorithm[71]. The unweighted graphs required for the computation of the Leiden communities were constructed based on the inverse of the distances between the $k$ nearest neighbours of the Barlow Twins encoded vectors from the selected tiles. To ensure the relevance of the Leiden clustering, the algorithm was run for 100,000 iterations. The computation was accelerated on a single NVIDIA A100 GPU with 80GB of RAM thanks to NVIDIA RAPIDS Suite of AI libraries (v24.4.0). Our implementation is available on GitHub at: https://github.com/IARCbioinfo/LeidenForTilesCommunity_accGPU . The resulting high-performance implementation allowed us to explore the following hyperparameters: The number of neighbours used to construct the graphs, denoted $K \in [75, 125, 250, 400]$, and the Leiden resolution parameter denoted $\gamma \in [0.25, 0.75, 1, 1.25, 1.5, 2, 3]$.

We explored 32 combinations between these parameters, repeating the calculation of the Leiden algorithm five times per combination, to explore the reproducibility of this clustering technique. To select the best clustering, the silhouette scores were calculated for each run according to the cosine distances. The silhouette scores were first compared based on the parameter $K$, and then according to $\gamma$. No value of $K$ was significantly associated with higher silhouette scores ($P$ value = 0.07, Kruskal-Wallis). $K$ = 75 was chosen because it allows a faster computation of Leiden partitions. For the Leiden resolution parameter $\gamma = 3$, was associated with the highest silhouette scores. Finally, for $K$ = 75 and $\gamma = 3$ the second run was associated with the best value and then used to approximate Leiden partitions (**Supplementary Fig. S25**).

Since the proportions of tiles in each of the resulting 116 Leiden partitions by WSI provide the information used to predict the diagnosis or molecular group of patients, each of the 3.5 million tiles had to be assigned to a Leiden partition. Therefore, for each tile, the partition of its nearest neighbour in the set used to calculate the Leiden groups was used as a proxy for its Leiden partition.

#### 9.4 Classification strategy based on Barlow Twins encoded vectors

Before predicting the patient diagnoses or molecular group, the approximate Leiden partitions were filtered according to the following rules. First, since the partitions must include tiles from a sufficient number of WSIs to represent a meaningful morphological partition, the inverse of the Simpson index, which measures the diversity of a composition vector with non-negative entries that sum to one that correspond to the proportions of individuals from different categories[72], was calculated for each partition. Partitions with a score $\leq 2$, indicating low diversity (e.g., a partition that was only found in 2 samples and in the same number of tiles in each sample), were discarded to ensure sample diversity within partitions. Secondly, since the partitions must be significantly enriched for a group to be informative for a classifier, the enrichment per group and per partition was calculated. Only partitions with one or more groups associated with an enrichment score below 0.5 or over 1.5 were retained.

These rules were applied to the 116 Leiden partitions to retain 27 partitions to predict the histological type of 212 patients (**Supplementary Fig. S13**), and 41 partitions to predict the molecular group of 193 patients (**Supplementary Fig. S14**). The proportions of tiles per partition and per patient were used to obtain a single vector per WSI[70]. These vectors of proportions were used to train random forest models to predict either the patient's diagnosis, i.e. typical or atypical, or their molecular group. A 'leave one out' strategy was used to predict once each patient once, based on a training set that included all the other patients. Weighted F1 scores were computed to evaluate and compare model performance. Results are presented in **Supplementary Tables S38 and S39**.

#### 9.5 RoFormer-MIL supervised deep learning model

The Multiple Instance Learning (MIL) RoFormer model[73] was used as a supervised deep learning approach to directly predict the molecular group of each WSI and to identify morphological partitions associated with these groups, particularly in the absence of established histological hypotheses. RoFormer-MIL offers the following advantages over the random forest model used in **section 9.4**: i) it captures interactions between tiles within the WSI using its Transformer module; ii) it accounts for the relative spatial positioning of the tiles within the WSI, and iii) it assigns importance to the individual tiles during classification through the "Attention-based Deep Multiple Instance Learning" (ABMIL) attention scores[74].

We replaced the original naïve tile encoding method, which used a pre-trained ResNet 50 on ImageNet, with the encoded vectors obtained from Barlow Twins. RoFormer-MIL was trained using a multi-fold train/test strategy, to predict each WSI once. Thus, the 193 WSIs were split 32-fold, with each fold containing five to nine samples in the test set and nine samples in the validation set.

We modified the architecture of the model to adapt RoFormer-MIL to 128-dimensional encoding vectors (**Supplementary Tables S40 and S41**). To mitigate overfitting, the following adjustments were made: i) encoded vectors were standardised by WSI, ii) ReLU activation functions were substituted for leaky ReLU activations functions[75], iii) Networks weights were initialised by with Xavier method[76], and iv) early stop mechanisms and checkpoint saving were configured on the minimal validation loss rather than the maximum binary accuracy as originally proposed.

#### 9.6 Leiden morphological partitions based on RoFormer-MIL highest attention scores

The top 5% of ABMIL attention scores by channel and WSI were used to select the encoding vectors to compute Leiden clusters. Only correctly predicted WSIs were used, as the aim of these clusters was to discover the significant morphological features that led to correct classification. In total, 286,097 vectors were selected, the same parameters as in the self-supervised branch were explored, except for the number of neighbours $K = 400$, which was excluded due to memory constraints. To select the best clustering, silhouette coefficients based on cosine distances were calculated for each run. The clusters obtained with $K = 250$ had a significantly higher silhouette coefficient. The $\gamma$ value that maximised the silhouette coefficient values was 2.0. Finally, the third run with $K = 250$ and $\gamma = 2$ was associated with the highest silhouette coefficient and was therefore chosen as the best cluster for interpreting the Leiden scores (**Supplementary Fig. S25**).

The resulting 75 Leiden partitions were then filtered according to the criteria defined in **Section 9.4**. The first criterion eliminated 34 partitions, and the second excluded 10 more, leaving 31 partitions after filtering (**Supplementary Fig. S16, Supplementary Table S42**).

#### 9.7 Pathological review to interpret deep learning-based results

##### *9.7.1 Partition level interpretability and selection of morphological features*

For each of the 31 remaining partitions, five of the six pathologists assessed their interpretability based on a random selection of 28 tiles per partition. The evaluation included three criteria: i) whether the partition needed to be excluded because it represented recurrent artefacts, such as blurred images, or damaged tissue; ii) whether the partition was homogenous enough to be described globally without requiring a tile-by-tile annotation; and iii) whether 50 tiles was estimated as sufficient to capture the morphological heterogeneity, or if additional tiles were required for a comprehensive description.

The results of this first step are summarised in **Supplementary Table S43**, where the majority rule was applied to aggregate the responses of all pathologists (**Supplementary Table S43**). Three partitions (9, 10, and 25) were excluded due to the presence of significant artefacts. Four partitions (28, 39, 58, and 70) were initially discarded because they contained few tumour cells, however, were reported to have other potential features of interest such as stroma and parenchyma and were therefore retained. Partitions 28, 39 and 70 were sufficiently described based on comments during this first evaluation step and were therefore considered globally annotated, whilst partition 58 underwent tile-by-tile annotation.

Of the 25 remaining partitions, 12 were deemed sufficiently homogeneous to be described globally, while 13 (including partition 58) required a tile-by-tile description due to significant morphological heterogeneity. To reduce the workload, partitions were distributed between two pathologists per group, with separate assignments for global and tile-by-tile annotation. A preliminary list of morphological features relevant to lung carcinoids was compiled for annotation. Pathologists reviewed this list, providing input on its relevance and suggesting additional features. All proposed modifications were incorporated into the final list for the second step of annotation. The selected features were grouped into the following categories:

- Cell composition:
    - Presence of immune cells (macrophages, lymphocytes, etc)
    - Stroma description: fibroblasts, endothelial cells, etc.
    - Additional non-tumoural cells (ciliated cells, chondrocytes, etc)
- Tissue description:
    - Presence of necrosis or fibrosis
    - Tissue architecture
- Description of tumour cells:
    - Tumour cell size
    - Nucleus-to-cytoplasm ratio
    - Tumour cell shape
    - Additional features (conspicuous nucleoli, salt-and-pepper chromatin, etc)

##### *9.7.2 Global annotations*

Partitions that were sufficiently homogeneous for global description were presented to one pair of pathologists using the 28 tiles from the first step. In this second step, each pathologist assessed whether the selected morphological features were present. **Supplementary Table S44** summarises these annotations. Each partition was independently reviewed by two pathologists.

##### *9.7.3 Tile-by-tile annotations*

Partitions exhibiting significant morphological heterogeneity were annotated on a tile-by-tile basis. A random selection of 50 tiles per partition was uploaded to the Label Studio web application (https://labelstud.io/) where pathologists reviewed individual tiles and assigned annotations by selecting our predefined features (**Supplementary Fig. S15**). Each partition was independently annotated by two pathologists, and all annotations are provided in **Supplementary Table S45**. For partition 58, an extended set of 75 tiles was annotated, while all other partitions followed the standard 50-tile annotation protocol.

##### *9.7.4 Interpretation of annotations*

To consolidate individual annotations, features were classified into three categories:

- 'Yes': indicating that the feature was identified by both pathologists, either at the tile level (for tile-by-tile annotations) or at the partition level (for globally annotated partitions).
- 'No': indicating that the feature was not identified by either pathologist.
- 'Maybe': indicating the feature was identified by only one of the two pathologists.

This rule was applied to all features except those associated with higher uncertainty, for which additional considerations were implemented:

- 'Tumour cell size' and 'N:C ratio' (nucleus-to-cytoplasm ratio): in the absence of a defined threshold or measuring tool, the annotation "unusually large" or "unusually small" was recorded only if both pathologists agreed. Otherwise, the feature was left unspecified (NA), with "medium" as the default classification.
- 'Tissue architecture': due to the inherent difficulty of assessing architecture in small tiles, only structures identified by both pathologists were considered. No architectural combinations were accepted, except for "organoid and trabecular", as trabecular is a subcategory of organoid architecture[77]. Additionally, the nested cell architecture was classified under organoid.

For tile-by-tile annotated partitions, a Fisher's exact test was performed to assess whether specific features were significantly enriched in a given partition compared to others, and *P* values were corrected for multiple testing using the Benjamini-Hochberg method (**Supplementary Table S45**, **Supplementary Figs. S17 and 18**).

Finally, feature prevalence was evaluated across molecular groups. A total of 29 features were evaluated. For partitions annotated tile-by-tile, the proportions of 'yes' and 'maybe' were summed, with the 'maybe' level assigned half the weight of the 'yes' level. For globally annotated partitions, 'yes' corresponded to 100% of the tiles displaying the feature, 'maybe' to 50% of the tiles, and 'no' to 0% of the tiles displaying the feature. Permutation tests (10,000 replicates) were performed to compare the means of the groups: a global test to compare the three groups, and pairwise tests for each pair of groups (**Supplementary Figs. S17 and S18**). *P* values were corrected for multiple testing using the Benjamini-Hochberg method, leading to 13 out of 29 features significant at the 5% threshold (**Supplementary Table S45**).

#### 9.8 Generalisation of the presence of key morphological features

To estimate the presence of identified morphological features based on pathological review, we used the CONCH[78] visual-language foundation model. All tiles were first embedded using the CONCH image encoder. To assess the relevance of specific text prompts corresponding to key features identified by pathologists, cosine similarity scores were computed for manually annotated tiles. For validation, the model was tested on selected annotated tiles. For example, using 100 tiles annotated as containing spindle-shaped cells and 451 tiles without this feature (as determined by two pathologists), the prompt "spindle shape" achieved a ROC-AUC score of 0.89. Similarly, the prompt "fibrotic tissue" yielded a ROC-AUC score of 0.74, based on 112 tiles containing this feature and 343 tiles without. Following this prompt evaluation step, cosine similarity scores were computed between each text prompt and all tiles. Finally, the median cosine similarity score was calculated per WSI, providing a single value per slide. These scores were then used to compare the distribution of morphological features across different molecular groups (**Fig. 3d**). In order to test the robustness of the results to the text prompt used, we repeated the analysis using 21 alternative prompts for spindle cells (e.g., "narrow spindle shape" or "narrow, elongated cells") and fibrosis (e.g., "fibrous tissue with collagen" or "fibrous connective tissue with collagen bundles"; **Extended Data Fig. 4d**).

##### 9.9 Morphological review for classification

H&E slides for 22 unclassified/misclassified cases from the immunohistochemistry panel classification (see **Section 11**) were provided to pathologist M.V. for review and classification. M.V was instructed to firstly indicate the presence or absence of four features (spindle cell shape, organoid architecture, solid architecture, fibrosis) by selecting one of four levels from a dropdown menu (absent, low presence, moderate presence, or high presence). Then he was asked to use this information to assign a molecular group of Ca A1, Ca A2, or Ca B based on the following guidelines. Ca A1: frequent spindle cells, solid architecture and fibrosis, rarely organoid architecture; Ca A2: frequent organoid architecture and fibrosis, few spindle cells; Ca B: frequent solid architecture, few spindle cells. Results are presented in **Supplementary Figure S12 and Supplementary Table S37**.

#### 10. Digital spatial profiling

##### 10.1 Experimental set up

To assess the heterogeneity of lung NETs and to characterise the tumour microenvironment, we used NanoString GeoMx's Digital Spatial Profiling (DSP) technology[79]. The DSP data combine immune cell and tumour cell areas from 64 patients in the lung NET cohort, including 25 Ca A1, 18 Ca A2, and 21 Ca B patients (**Supplementary Fig. S5**). A total of 513 areas of interest (AOIs) were selected blindly, with regard to the molecular group, and processed at the Centre Léon Berard (**Supplementary Fig. S5**). Immune AOIs were selected based on CD45 expression assessed by UV illumination, and tumour cell AOIs by fluorescence of PanCk. The expression of 39 proteins from three panels (immune cells, immune activation state, and immune cell typing) was spatially quantified using the NanoString nCounter® platform.

##### 10.2 DSP data quality control and normalisation

We performed quality control on the selected AOIs and protein probes according to the instructions in the GeoMx Data Analysis and nCounter user manuals, as well as the protein normalisation strategy. A total of 44 AOIs (8.6%) were excluded (**Supplementary Fig. S5**): four due to mixed tumour/immune region design, 16 due to a low number of nuclei (i.e. less than 20), six due to a low surface area (i.e. less than 1600 μm2), 15 due to lack of collected material, and three with insufficient expression of the housekeeper probes S6, Histone H3 and GAPDH (i.e. the logarithm of their geometric mean was considered an outlier with respect to the rest of the distribution, being less than five) or too low expression of the negative control probes Rb IgG, Ms IgG2 (i.e. the logarithm of their geometric mean was less than three with respect to the rest). This resulted in available data for 469 AOIs.

To check the quality of the protein measurements, the signal to background ratios of each probe were calculated, where the background signal was defined as the geometric mean of the negative controls. We excluded all proteins for which the mean $log_2$ signal to background ratio was less than zero. Nine proteins were excluded according to the quality control performed within tumour cell AOIs, i.e. AOIs fluorescing at PanCk: CD66b, CD163, CD80, PD-L1, CD27, ICOS, PD-L2, CD40, and PD-1. For immune cells AOIs (fluorescently labelled with CD45), five proteins were excluded: CD80, PD-L2, CD66b, PD-L1, and FOXP3. As tumour and immune cell AOIs were analysed together, a common set of non-control proteins that passed this quality control step was determined, resulting in the inclusion of 23 immune-related proteins (**Supplementary Fig. S5**).

##### 10.3 Molecular group prediction for AOIs

To evaluate the ability of the selected immune proteins to distinguish the three main molecular groups, namely Ca A1, Ca A2, and Ca B, we employed random forest (RF) models to predict individual AOIs based on the molecular group defined at the patient level through multi-omics analysis (see **Section 8.1**). The original RF model (RF1) used normalised counts of 23 proteins as the input features and were trained using a five-fold cross-validation approach. This training process was repeated 100 times, resulting in each AOI being predicted 100 times. The probabilities for each class were averaged across the 100 predictions. AOIs were deemed unclassifiable if the ratio of the highest to second-highest average probability was less than 1.5; otherwise, the AOIs were assigned to the molecular group corresponding to the most probable class. The model's performance at the AOI level was evaluated using contingency matrices and F1 scores (**Supplementary Fig. S6, Supplementary Table S13**). AOIs were deemed correctly classified if the predicted molecular group corresponded with the molecular group assigned at the patient level. We hypothesised that misclassification of AOIs might result from intra-tumour heterogeneity in the immune microenvironment across AOIs from the same patient rather than inaccuracies in the RF1 model. To test this hypothesis, a new set of RF models (RF2) was trained only on AOIs that had been correctly classified, because these AOIs presumably represent AOIs with immune microenvironments matching the patient-level group label, using the same methodology as in RF1. We then performed a prediction for AOIs that had been misclassified by the RF1 model using RF2. The high reproducibility of predictions between the original RF1 and the RF2 models (**Supplementary Fig. S6, Supplementary Table S13**) validated the reliability of the decision rules. The final molecular group prediction for each AOI, used in subsequent analyses, was inferred from RF2 models, and AOIs for which the predicted molecular group differed from the patient-level assignment were interpreted as indicative of intra-tumoural heterogeneity (**Supplementary Fig. S6, Supplementary Table S13**). The intra-tumoural heterogeneity of immune proteins was illustrated by the prediction of each AOI per patient (**Supplementary Fig. S8**). Furthermore, this intra-tumoural heterogeneity within each molecular group was summarised by the prediction frequencies of each molecular group (**Supplementary Fig. S8**). Finally, for each molecular group a Binomial test was performed to determine whether one of the two groups that contradicted the patient's true group was more likely (**Supplementary Table S13**).

##### 10.4 The most discriminating immune proteins of molecular groups

To rank the proteins based on their ability to discriminate the three molecular groups, RF models were trained to classify the molecular groups of the AOIs, and the mean decrease in Gini index for each protein was recorded. In this third set of RF models, only AOIs that were correctly classified by the second set of models, meaning their predictions matched the molecular group defined at the patient level, were included. The models were trained using a five-fold cross-validation strategy, repeated 100 times. The average decrease in Gini index for each protein across all models was then calculated to rank the proteins according to their discriminatory power for molecular group determination (**Supplementary Fig. S7, Supplementary Table S14**). To determine the minimum number of proteins needed to replicate the classification performance of the second set of RF models, new RF models were trained using progressively larger sets of the top *n* most predictive proteins from the previous ranking. The same training strategy was applied. For each set of these new RF models, the weighted average F1 score of the classifications was calculated and reported (**Supplementary Fig. S7**). Subsequently, the elbow method was employed to identify the smallest set of proteins that achieved the performance of the second set of RF models. For each protein in this optimal set, normalised counts were reported by patient molecular group and according to the molecular groups predicted at the AOI level (**Supplementary Fig. S7**).

#### 11. Immunohistochemistry classification panel

Ninety samples from the lung NET cohort with FFPE tissue available that had been classified as Ca A1, Ca A2 or Ca B as described in **Section 8.1** were randomly selected for assessment by immunohistochemistry (IHC). IHC staining for ASCL1, HNF1A, and OTP was performed at Maastricht University Medical Centre following the protocol described in Leunissen *et al.*[80]. H-score cutoffs for classification as Ca A1, Ca A2, and Ca B are as follows: OTP $\geq$ 40 & ASCL1 $\geq$ 10 (Ca A1); OTP $\geq$ 150 & HNF1A $\geq$ 30 (Ca A2); OTP $\leq$ 20 & HNF1A $\geq$ 80 (Ca B). H-scores, predicted molecular group, and true molecular group are shown in **Supplementary Table S37**, example images are shown in **Supplementary Fig. S12**.
